# Development and evaluation of an open-source, standards-based approach to explainable artificial intelligence for managing co-morbidity and clinical guidelines using argumentation techniques and the Transition-based Medical Recommendation model

**DOI:** 10.1101/2022.12.12.22283312

**Authors:** Jesús Domínguez, Denys Prociuk, Branko Marović, Kristijonas Čyras, Oana Cocarascu, Francis Ruiz, Ella Mi, Emma Mi, Christian Ramtale, Antonio Rago, Ara Darzi, Francesca Toni, Vasa Curcin, Brendan Delaney

**Author notes:** Joint first authors (Jesús Domínguez, Denys Prociuk). Joint last authors (Francesca Toni, Vasa Curcin, Brendan Delaney).

## Abstract

I.

**Objective:** Clinical Decision Support (CDS) systems (CDSSs) that integrate clinical guidelines need to reflect real-world co-morbidity. In patient-specific clinical contexts, transparent recommendations that allow for contraindications and other conflicts arising from co-morbidity are a requirement. We aimed to develop and evaluate a non-proprietary, standards-based approach to the deployment of computable guidelines with explainable argumentation, integrated with a commercial Electronic Health Record (EHR) system in a middle-income country.

**Materials and Methods:** We used an ontological framework, the Transition-based Medical Recommendation (TMR) model, to represent, and reason about, guideline concepts, and chose the 2017 International Global Initiative for Chronic Obstructive Lung Disease (GOLD) guideline and a Serbian hospital as the deployment and evaluation site, respectively. To mitigate potential guideline conflicts, we used a TMR-based implementation of the Assumptions-Based Argumentation framework extended with preferences and Goals (ABA+G). Remote EHR integration of computable guidelines was via a microservice architecture based on HL7 FHIR and CDS Hooks. A prototype integration was developed to manage COPD with comorbid cardiovascular or chronic kidney diseases, and a mixed-methods evaluation was conducted with 20 simulated cases and five pulmonologists.

**Results:** Pulmonologists agreed 97% of the time with the GOLD-based COPD symptom severity assessment assigned to each patient by the CDSS, and 98% of the time with one of the proposed COPD care plans. Comments were favourable on the principles of explainable argumentation; inclusion of additional co-morbidities were suggested in the future along with customisation of the level of explanation with expertise.

**Conclusion:** An ontological model provided a flexible means of providing argumentation and explainable artificial intelligence for a long-term condition. Extension to other guidelines and multiple co-morbidities is needed to test the approach further.

**Funding:** The project was funded by the British government through the Engineering and Physical Sciences Research Council (EPSRC) – Global Challenges Research Fund.^1^

## II. Research in context panel

### A. Evidence before this study

We searched PubMed and Google Scholar without time or language restriction for the terms ‘Clinical Decision Support [MESH and text all fields) AND ‘Argumentation’ [Text, all fields]. 25 articles were found, all of which were discussion or theoretical papers. Although there is an extensive literature on Clinical Decision Support over more than 40 years with many information models, software integration and evaluations, there have been no studies that have applied computational argumentation approaches in the clinical domain. Clinical Decision Support (CDS) has been shown to provide benefit in clinical settings in terms of adherence to clinical guidelines, screening and preventive activities, diagnosis, and prescribing. However, increasing co-morbidity and the wide range of clinical guidelines that may apply to an individual patient have led to difficulty in providing clear and explicit guidance. In addition, proprietary and electronic health record (EHR)-based solutions for maintaining knowledge for CDS increase costs and constrain the sharing of computable knowledge resources in Low- and Middle-Income Countries (LMIC) settings.

### B. Added value of this study

The Transition-based Medical Recommendation (TMR) model is an ontology-based framework for representing, and reasoning about, statements from clinical guidelines. The TMR model does not, however, incorporate the reasoning necessary to determine possible resolutions to clinical guideline conflicts, nor personalise recommendations with explanations. In this study, and using the 2017 International Global Initiative for Chronic Obstructive Lung Disease guideline in a Serbian hospital as an exemplar, we showed that the TMR model can be enhanced for conflict resolution by using the state-of-the-art assumptions-based argumentation extended with preferences and goals so that explainable patient-centred treatment plan options can be provided to clinicians and have the potential to impact care.

### C. Implications of all the available evidence

Given the increasing levels of co-morbidity in chronic conditions, CDS without the management of conflicts among clinical guidelines in EHR systems will become increasingly detached from the real world of patient care. In addition, rates of non-communicable disease are rising in LMIC and solutions to this problem must be open and affordable. Combining ontology-based representations of clinical guideline statements such as those represented by the TMR model, with approaches to argumentation based on artificial intelligence, opens the potential to create standards-based, shareable, and interacting computable guidelines that can be enacted across heterogeneous FHIR-compliant EHR systems.

## III. Background

Increasingly, developers of clinical guidelines, such as the National Institute for Health and Care Excellence (NICE)^2^ in the UK, have sought to implement aspects of guidance via encouraging the creation of computational algorithms, often proprietary, within Electronic Health Records (EHR). However, multimorbidity, defined as the coexistence of two or more chronic medical conditions in an individual patient,^3^ is common in the real world and presents a multitude of competing priorities, potential contraindications, and guideline exceptions to the clinician. Although studies have shown clinical decision support (CDS) improves clinicians’ adherence to clinical and operational guidelines for medication, prevention, and treatment,^4–7^ problems with ‘alert fatigue’, confusion and contradiction between different CDS alerts can represent a threat to patient safety.^8^ Computable Guidelines (CGs), machine-interpretable versions of guidelines, have the potential to alleviate some of this burden on the clinician by using ‘argumentation’, i.e., defining the ‘best’ option from a series of logical statements.^9^ However, in Low- and Middle-Income Countries (LMIC), the resources required for proprietary EHR-integrated CDS systems (CDSSs), localization and maintenance are often not available.^10^ In contrast, open source code can be written to extract data and trigger a rule externally (for example using both the FHIR API (a global standard to promote data-level interoperability among disparate EHRs) and CDS Hooks^11^ (a specification for standardising the seamless integration of external services in EHRs). Growing functionality in CDS can be represented as multiple interacting models and ontologies as written rules are replaced by computable ones,^12^ allowing, for example, a model of the patient’s current clinical state to present data to a model of appropriate guideline statements, applied to derive a care recommendation.

To meet the challenge of co-morbidity, a model of clinical reasoning between conflicting statements is required. Argumentation models amount to automated systems that emulate human reasoning,^13^ positioning arguments and counterarguments for a given issue to find the ‘winning’ arguments. Argumentation is a good fit for modelling patient-centric reasoning with multiple guidelines where interacting recommendations and alerts give rise to conflicts, and the context of a patient brings in various conditions, goals, and preferences.^14^

The ROAD2H project aimed to develop and evaluate a representative CDSS for embedding and enacting the internationally accepted Global Initiative for Chronic Obstructive Lung Disease^15^ (GOLD) guideline,^1^ illustrating the role of argumentation-based techniques in presenting conflict-safe care plan proposals to clinicians, and integrating the system with a modern standards-based commercial EHRs in a middle income country (Heliant,^16^ the largest healthcare information systems provider in Serbia) using a combination of knowledge representation and interoperability standards.

## IV. Methods

### A. Clinical case study

We chose the management of Chronic Obstructive Pulmonary Disease (COPD) with cardiovascular (CVD) or chronic kidney disease (CKD) co-morbidities as an exemplar. The GOLD guideline classifies the COPD symptom severity as a series of stages or groups, each with recommended treatments in a ‘step up’ fashion. In addition, some of the therapies are contraindicated in co-morbidity, for example, beta agonist medications with angina.^15^ COPD is increasingly common in LMIC, so we aimed at designing a CDSS that integrates both COPD symptom severity assessment and treatment planning with the workflow of pulmonologists in the EHR. Requirements were to identify potential treatment conflicts and suggest alternative care plans.

### B. The Transition-based Medical Recommendation (TMR) model

To reason about potential interactions among, or within, guidelines, we adopted the TMR model,^4,17^ a formalism to represent guidelines and detect conflicts using semantic web technologies and logic rules. TMR-represented recommendations (see Table 1) comprise a care action and its causation belief, that is, the expected effect on the measured property the care action affects. Care actions promote transitions which comprise the initial (clinical) state and the expected state of the affected measured property. Care action effects are either positive or negative, potentially increasing or decreasing the value of the measured property. There are implementations of TMR for prototyping guideline representation (using the RDF model for guideline representation and SPARQL^18^ as its query language) and interactions theory (using the logic-based programming language SWI-Prolog); however, neither implementation offers enactment nor automated merging of CG statements. Like other multimorbidity-oriented formalisms,^5,6^ TMR currently lacks the reasoning capabilities to resolve the identified interactions and does not consider patient-specific conditions, goals, or treatment and lifestyle preferences.^7^ The computational argumentation formalism described next aims to integrate all these elements to provide automated reasoning with interacting TMR-based CGs, considering the patient’s context and preferences.

**Table 1.** Recommendations based on GOLD guideline, represented as TMR knowledge. SAMA stand for Short-Acting Muscarinic Antagonist, SABA stands for Short-Acting Beta-Agonist, ALS stands for Airflow Limitation Severity, CB stands for Causation Belief, and Rec for Recommendation.

| Rec ID | English representation | Care action | CB ID | Effect | Measured property | Initial state | Expected state after care action application | Expected transition on initial state of measured property |
| --- | --- | --- | --- | --- | --- | --- | --- | --- |
| R<sub>sama</sub> | Recommend administering SAMA bronchodilator to COPD patients with mild ALS | Administer SAMA bronchodilator | CB1 | + | ALS | Mild ALS | Mild ALS | maintain |
| R<sub>saba</sub> | Recommend administering SAMA bronchodilator to COPD patients with mild ALS | Administer SABA bronchodilator | CB2 | - | ALS | Mild ALS | Mild ALS | Maintain |
| R<sub>beta</sub> | COPD patients with co-morbid cardiovascular disease should be aware that beta-agonists bronchodilators could increase the risk of cardiac rhythm disturbances | Administer beta-agonist bronchodilator | CB3 | + | At risk of cardiac rhythm disturbances | Low risk | High risk | Increase |
| R<sub>jab</sub> | Recommend administering pneumococcal vaccine to patients over 64 years of age | Administer pneumococcal vaccine | CB4 | - | At risk of pneumonia | High risk | Low risk | decrease |

### C. Explainable argumentation

To reason over the guideline conflicts, we made use of the *Assumption-Based Argumentation with Preferences and Goals*^*14*^ (ABA+G) technique which represents knowledge using a formal (logical) language, rules, and defeasible assumptions. Rules and assumptions allow for a transparent and interpretable representation of the TMR concepts, particularly recommendations and their components, as illustrated in Figure 1, where interactions among recommendations can likewise be captured via rules. For example, and following Figure 1, the contradictory interaction between recommendations *R*_*saba*_ and *R*_*beta*_ is expressed via two rules that assume *R*_*saba*_ leads to an objection against *R*_*beta*_, and vice versa. Moreover, since *R*_*sama*_ is identified as an alternative recommendation to *R*_*saba*_, then acceptance of R_*beta*_ leads to the acceptance of R_*sama*_ by application of a rule which assumes both that R_*beta*_ and R_*Saba*_ object each other and that R_*beta*_ and R_*sama*_ do not object each other.

**Figure 1.**
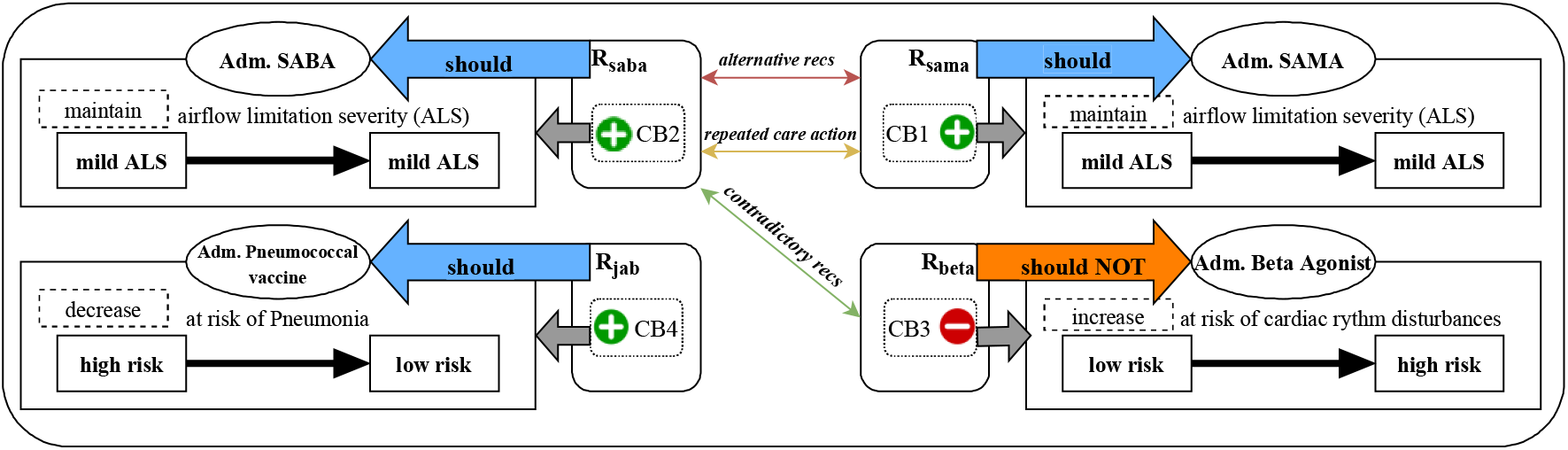
Graphical representation of the recommendations in Table 1, and their identified potential interactions when administered together. Adm. stands for Administer, recs stand for Recommendations, CB for Causation Belief, SABA stands for Short-Acting Beta-Agonist bronchodilator, and SAMA for Short-Acting Muscarinic Antagonist bronchodilator.

Explainability, as in not only providing explanations accompanying reasoning outcomes, but also the overall transparency in knowledge representation and reasoning mechanisms, is a fundamental requirement of CDSS.^19^ Argumentation is itself an explainable reasoning paradigm^20^, and it has also given rise to explanations in various settings, including AI.^21–23^ ABA+G natively affords means to explain its reasoning outcomes. Specifically, ABA+G yields explanations of the reasoning trace, its actions and expected effects. Explanations summarise the considered interactions and preferences, and accompany each proposed set of recommendations (e.g., Table 2). A TMR-based implementation of ABA+G (hereinafter the conflict resolution service) was utilized in this project.^24^

**Table 2.** Example of the conflict resolution service response to the TMR knowledge and interactions in Figure 1. The structured clinical knowledge from Table 1 is also part of the response but omitted here. Underlined words on column Generated explanation denote the fixed parts of the explanation template. Rec(s) stands for Recommendation(s). Symbol + (-) stands for preferable (less preferable). Column Extensions denotes to which extension(s) belongs the recommendation.

| Rec ID reference | Generated explanation | Alternative recs | Repeated care action | Contradictory recs | Extensions |
| --- | --- | --- | --- | --- | --- |
| R<sub>sama</sub> | administration of SAMA <u>has</u> positive contribution <u>on</u> airflow limitation severity <u>to</u> maintain <u>from</u> mild airflow limitation severity <u>to</u> mild airflow limitation severity | R<sub>saba</sub> (+) | R<sub>saba</sub> (+) | .. | ext1 |
| R<sub>saba</sub> | administration of SABA <u>has</u> positive contribution <u>on</u> airflow limitation severity <u>to</u> maintain <u>from</u> mild airflow limitation severity <u>to</u> mild airflow limitation severity | R<sub>sama</sub> (-) | R<sub>sama</sub> (-) | R<sub>beta</sub> | ext2 |
| R<sub>beta</sub> | administration of beta agonist *has* negative contribution *on* at risk of cardiac rhythm disturbances *to* increase *from* low risk of cardiac rhythm disturbances *to* high risk of cardiac rhythm disturbances | .. | .. | R<sub>sama</sub> | ext1 |
| R<sub>jab</sub> | administration of pneumococcal vaccine *has* positive contribution *on* at risk of pneumonia *to* decrease *from* high risk of pneumonia *to* low risk of pneumonia | .. | .. | .. | ext1; ext2 |

### D. Integration with heterogeneous EHR systems

We designed a general-purpose ontology-based CDSS architecture based on open-source and industry standards that consists of an interoperability service, and a CGs enactment architecture which includes ABA+G and extends on an existing CGs authoring microservice architecture.^25^ The interoperability service is common to any implementation and uses SNOMED CT and HL7 FHIR standards to exchange healthcare data with EHRs. Additionally, CDSS-subscribed EHRs invoke event-specific CDS according to CDS hooks specification standards where notifications for CDS and responses are delivered in the form of hooks and cards, respectively.^11^ Cards convey information determined by implementations of the CDSS architecture for specific CG formalisms and CDS events (e.g., TMR and COPD treatment planning, respectively). The CDSS architecture is illustrated in Figure 2. A TMR-based implementation is discussed next.

**Figure 2.**
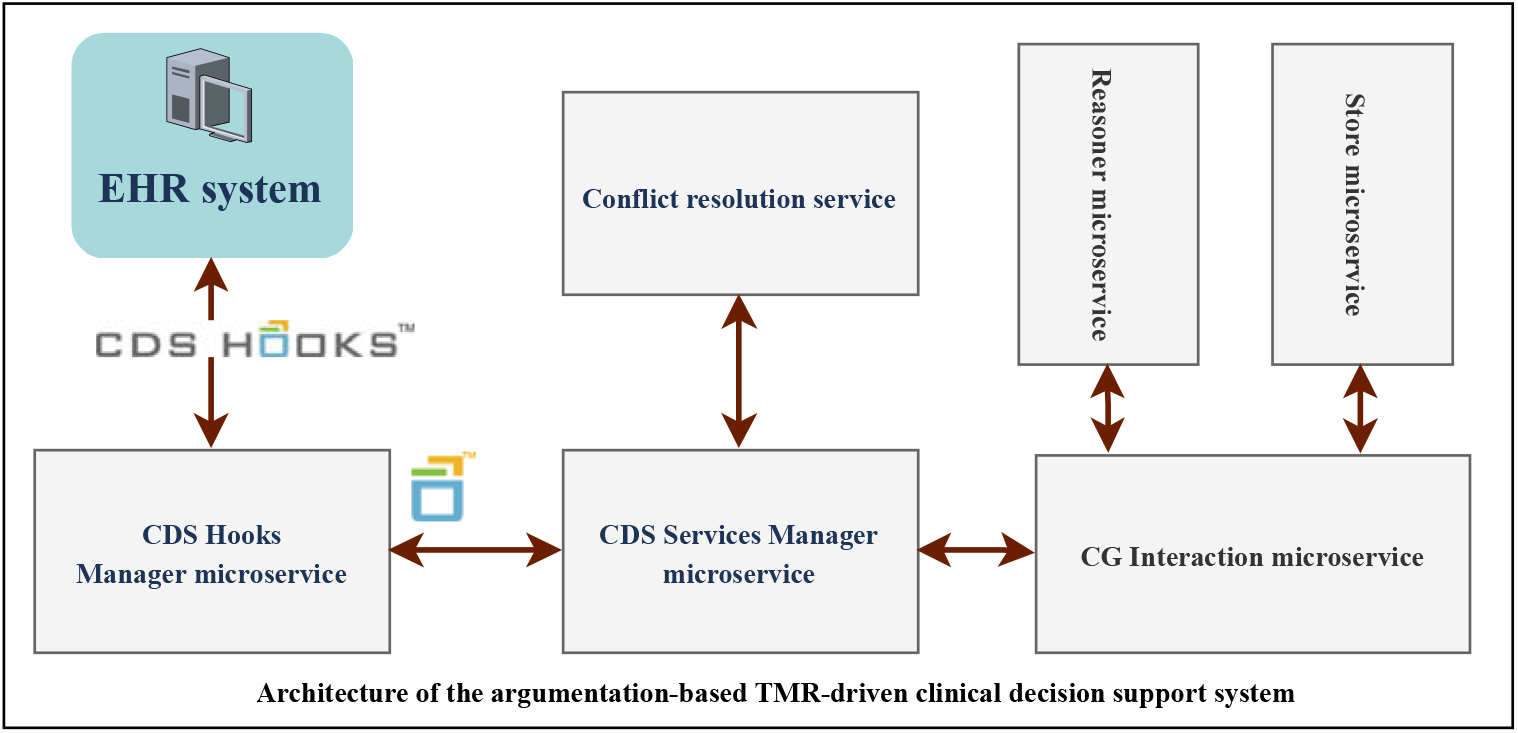
The ontology-based open-source CDS architecture. It is comprised of a fully developed interoperability service, labelled as CDS Hooks Manager microservice in the image, and the CGs enactment architecture (the remaining suite of services). A description can be found in Appendix A.3. The yellow, green, and blue square symbol represents CDS services returning any number of cards in response to CDS hooks notifications.

### E. Evaluation framework

To enable argumentation over TMR, we developed a suite of microservices (hereinafter the [TMR-based] CDS framework) to enact, and personalise, TMR-based CGs that includes the conflict resolution service and leverages a TMR-based implementation of the CGs authoring microservice architecture.^25^ This implementation encapsulated the previously mentioned existing TMR technology to store, query, and reason about TMR-based knowledge.^25^

Patient-relevant parts of implemented TMR-based CGs are triggered in the CDS framework by event-specific data included in the context of the CDS call. Each triggered recommendation is identified by a pair of unique RDF URIs referencing the recommendation and its CG, and combined into a volatile dataset using SPARQL. TMR interaction detection rules are then applied to this dataset. The dataset and detected potential interactions are encoded in JSON alongside user-defined preferences included in the hook’s context, then forwarded to the conflict resolution service. The response of this service consists of a collection of ‘extensions’ originating from the dataset, where each extension comprises TMR recommendations aggregated by considering potential interactions within the extension and preferences. The *explainability* component then refactors the encoded knowledge from each recommendation into information for CDS in both computer-interpretable and textual form. Finally, the response is embedded into a card by mapping extensions to FHIR carePlan types, resulting in personalized conflict-free care plan proposals comprising triggered recommendations potentially from multiple CGs and which include mitigation information on potential interactions among triggered recommendations from the source dataset (e.g., alternative recommendations distributed into distinct proposals). Figure 3 provides a modelled workflow of the system.

**Figure 3.**
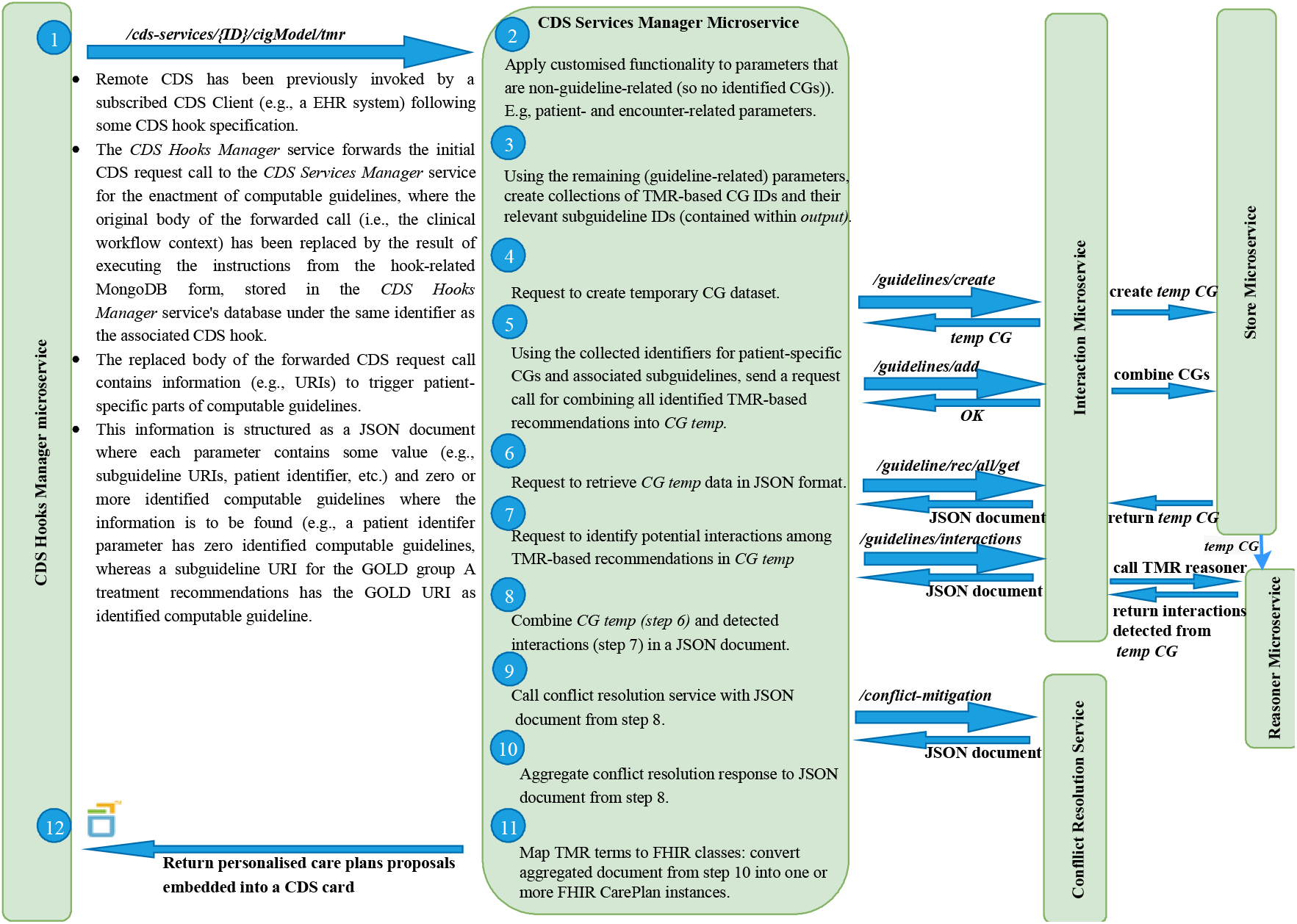
Workflow of the TMR-based COPD framework. Modelled workflow of how CDS calls are handled by the CDS Services Manager microservice in the argumentation-based, TMR-driven CDS system.

To evaluate our proposed CDS approach, we aimed to formalise key aspects of GOLD for stable COPD, including pharmacological, vaccination, physiotherapy, and smoking cessation therapies, plus drug-disease warnings for CVD and CKD co-morbidities. Appendix A.1 discusses the steps towards GOLD guideline formalisation using TMR. Subsequently, we defined hook specifications ‘copd-assess’ (Table 3) and ‘copd-careplan-review’ for COPD management CDS (Table 4) and worked with Heliant to integrate the remote CDSS with their EHR via a graphical user interface (GUI) add-on embedded in the EHR’s COPD tab.

**Table 3.**
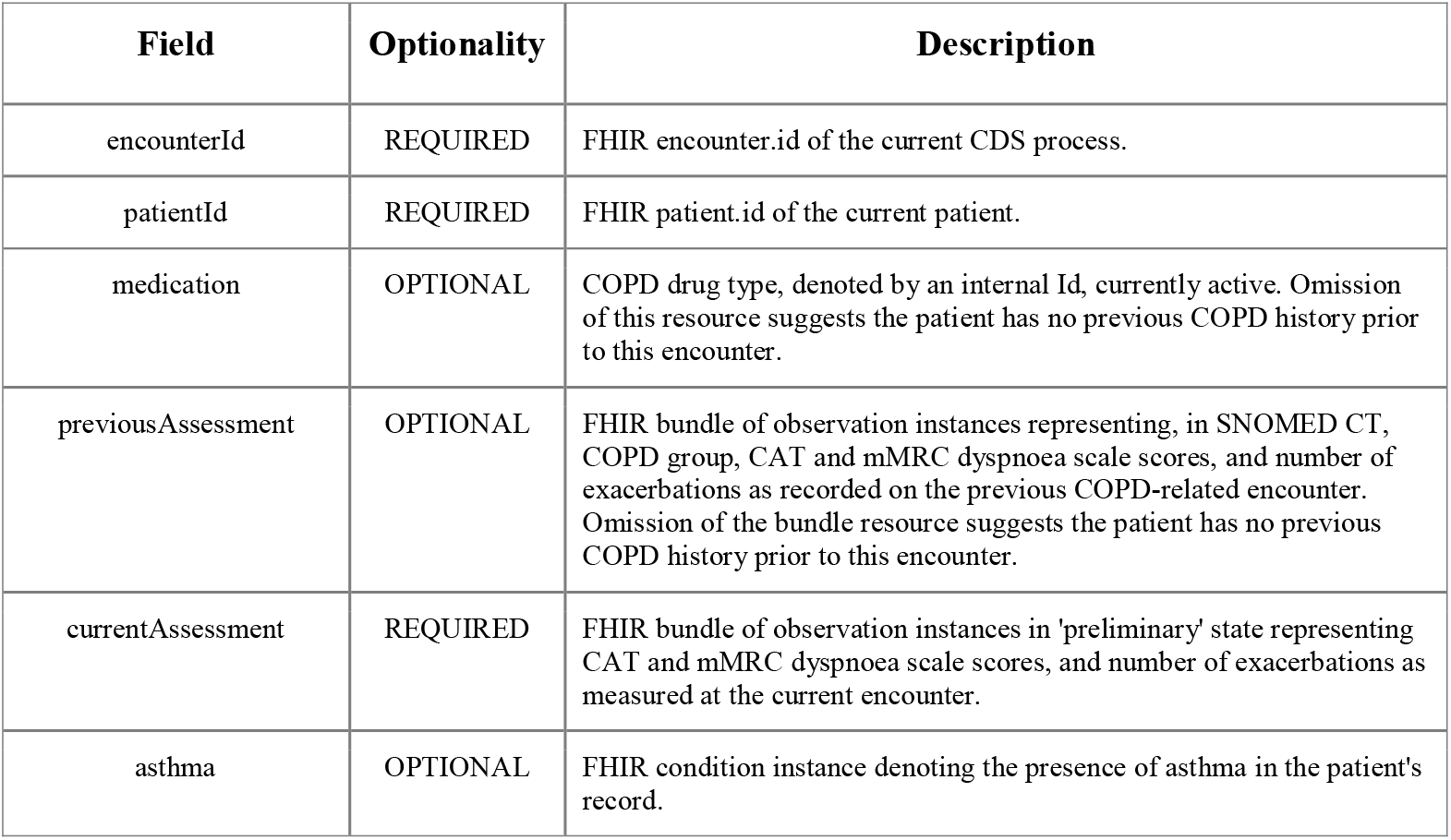
Contextual information for hook ‘copd-assess’. This CDS service collects patient data and COPD-related measurements to assess the patient’s COPD symptom severity, following GOLD’s ABCD assessment algorithm, and to provide an ordered collection of COPD treatments, from most to least suitable to the patient, for each of the GOLD groups.

**Table 4.** Contextual information for hook ‘copd-careplan’. This CDS service collects patient data from the EHR to propose one or more personalized COPD treatment management care plans.

| Field | Optionality | Description |
| --- | --- | --- |
| encounterId | REQUIRED | FHIR encounter.id of the current CDS process |
| patientId | REQUIRED | FHIR patient.id of the current patient |
| birthDate | REQUIRED | Date of birth of the current patient. Supports decision on suggesting administering pneumococcal immunization to patients over 64 years of age |
| smokingStatus | REQUIRED | FHIR observation instance representing, via a SNOMED CT term, the |
|  |  | patient as either a regular smoker or not. |
| co-morbidities | OPTIONAL | FHIR bundle of condition instances representing, via SNOMED CT terms, active diagnoses in the record of the current patient. |
| immunizationStatus | REQUIRED | FHIR bundle of immunization instances denoting, via SNOMED CT terms, whether the patient has completed the annual influenza vaccine, or the pneumococcal vaccine. |
| copdAssessment | REQUIRED | FHIR bundle of observation instance and Medication bundle. The former represents, in SNOMED CT, the user-selected COPD group. The latter represents, using internal Ids, the user-selected COPD drug types requested for CDS. |
| cdsSuggestedTreatments | OPTIONAL | FHIR bundle of medication instances suggested by the COPD-CDS system as a response to hook 'copd-assess' for the current patient. The bundle is aggregated to the conflict resolution input document to provide alternatives to user-selected COPD drug types when resolving potential conflicts among recommendations. |

We used simulated case vignettes to create dummy EHRs as access to real patients was prohibited by COVID-19 restrictions. Two clinical authors (Ella Mi, Emma Mi) created 20 cases. Each case contained data as illustrated in Table 5. All GUI textual outputs were rendered in Serbian and validated by a Serbian clinician.

**Table 5.** Clinical case vignette of patient introduced in Figure 4. Patient demographics, previous COPD clinical history and recorded number of exacerbations at current COPD assessment were populated into the EHR prior the evaluation. The remaining fields are entered by each clinician at evaluation time. SNOMED CT highlights that the condition or observation is recorded in the EHR using this clinical classification.

| Field | Value |
| --- | --- |
| Age | 65 |
| Sex | Male |
| Smoking status (SNOMED CT) | Ex-smoker |
| GOLD | 1 |
| COPD drug type administered at last COPD assessment | None |
| Number of exacerbations in past 12 months at last COPD assessment | 0 |
| CAT score at last COPD assessment (SNOMED CT) | 0 |
| mMRC dyspnoea scale score at last COPD assessment (SNOMED CT) | 0 |
| COPD group assessed at last COPD assessment (SNOMED CT) | None |
| Number of exacerbations in past 12 months at current COPD assessment | 0 |
| CAT score at last COPD assessment (SNOMED CT) | 6 |
| mMRC dyspnoea scale score at last COPD assessment (SNOMED CT) | 1 |
| Asthma (SNOMED CT) | False |
| Influenza vaccine (SNOMED CT) | Completed |
| Pneumococcal vaccine (SNOMED CT) | Not-done |
| Co-morbidities (SNOMED CT) | Cardiovascular disease, hypertension |

This was a mixed-methods evaluation to analyse quantitatively the validity of the recommendations and qualitatively the clinicians’ impressions of the approach. We recruited a purposive sample of five pulmonologists from Clinical Hospital Center Zvezdara in Serbia to use the extended Heliant EHR with each of the 20 cases, providing 100 cases in total. Firstly, we aimed to determine their agreement with the results of both CDS services for each clinical case. Secondly, after operating the system we interviewed each clinician using a structured questionnaire.

## V. Results

### A. COPD management specialisation of the CDS framework

To evaluate our CDS approach for the management of patients with stable COPD, TMR representations of selected GOLD recommendations were defined and loaded into a dedicated database. Similarly for the pair of hook context processing instructions described in Tables 6 & 7, based on the specifications in Tables 3 & 4. This resulted in the specialisation of the TMR-based CDSS for COPD symptom severity assessment and treatment planning decision-making, hereinafter the COPD-CDSS.

**Table 6.** Collection of JSON-based documents applied to contextual data of CDS hook ‘copd-assess’. The collection is uploaded to the NoSQL database of the interoperability service when the CDS service is invoked by any subscribed EHR. Each document provides instructions to query, and manipulate, specific parts of the clinical workflow context data towards delivering CDS. Below, Column Document label identifies each instructions-filled document in the collection. Column Application states the semantics of each document.

| Document label | Application |
| --- | --- |
| copdSeverityAssessment | Returns a SNOMED CT-based GOLD COPD group identifier for the active patient obtained by analysing their current COPD assessment results. |
| goldGroupA_treatmentPriorities | Assuming the active patient has a GOLD COPD group A symptoms severity, it returns an ordered list of suitable treatments by analysing their asthmatic status along with both previous and current COPD assessment results. |
| goldGroupB_treatmentPriorities | Assuming the active patient has a GOLD COPD group B symptoms severity, it returns an ordered list of suitable treatments by analysing their asthmatic status along with both previous and current COPD assessment results. |
| goldGroupC_treatmentPriorities | Assuming the active patient has a GOLD COPD group C symptoms severity, it returns an ordered list of suitable treatments by analysing their asthmatic status along with both previous and current COPD assessment results. |
| goldGroupD_treatmentPriorities | Assuming the active patient has a GOLD COPD group D symptoms severity, it returns an ordered list of suitable treatments by analysing their asthmatic status along with both previous and current COPD assessment results. |
| encounterID | Returns ID of this encounter. |
| patientID | Returns ID of this patient. |
| patientID | Returns ID of this patient. |

**Table 7.** Collection of JSON-based documents applied to contextual data of CDS hook ‘copd-careplan-review’. The collection is uploaded to the NoSQL database of the interoperability service when the CDS service is invoked by any subscribed EHR. Each document provides instructions to query, and manipulate, specific parts of the clinical workflow context data towards delivering CDS. Below, Column Document label identifies each instructions-filled document in the collection. Column Application states the semantics of each document.

| Document label | Application |
| --- | --- |
| co-morbidities | Returns the TMR-based CG ID for chronic kidney or cardiovascular diseases whenever a SNOMED CT-based term found in the hook context is subsumed, or equals to, the SNOMED CT codes for chronic kidney or cardiovascular diseases or history of the diseases. |
| selected_copd_group | Returns the subguideline ID for the GOLD COPD treatment pathways associated with the selected COPD Group. |
| additional_selected_meds | Returns the COPD-based subguideline ID(s) of additional COPD drug types that although are not officially part of the initial selection of GOLD COPD group drug types, are suitable to the active patient. |
| smoking_status | Returns the COPD-based subguideline ID of a smoking cessation recommendation if the patient is identified, via SNOMED CT, as a smoker. |
| influenza_immunisation | Returns the COPD-based subguideline ID of the influenza immunization recommendation if the patient has not completed the seasonal influenza vaccination. |
| pneumococcal_immunization | Returns the COPD-based subguideline ID of the pneumococcal immunization recommendation if the patient's age is over 64 and no pneumococcal jab administration is recorded as completed. |
| selectedTreatmentPathways | Returns the list of COPD drug treatments selected by the user for requesting CDS. |
| alternativeTreatmentPathways | Returns the list of COPD drug treatments suggested by the COPD-CDS system as a response to hook ‘copd-assess’. |
| encounterID | Returns ID of this encounter. |
| patientID | Returns ID of this patient. |

The COPD-specialised GUI interacts with the COPD-CDSS by collecting COPD-related measurements and other relevant clinical details stated in both hooks specifications, to aid with both clinical events. Each event invokes a CDS service, triggered their respective buttons in the GUI (see Figure 4). Using a GUI form to collect or modify input was a design choice made by Heliant that benefitted the pulmonologists when evaluating the COPD-CDSS and was preferred by Heliant over the direct collection of relevant EHR data, which was perceived as less transparent and insufficiently interactive.

**Figure 4.**
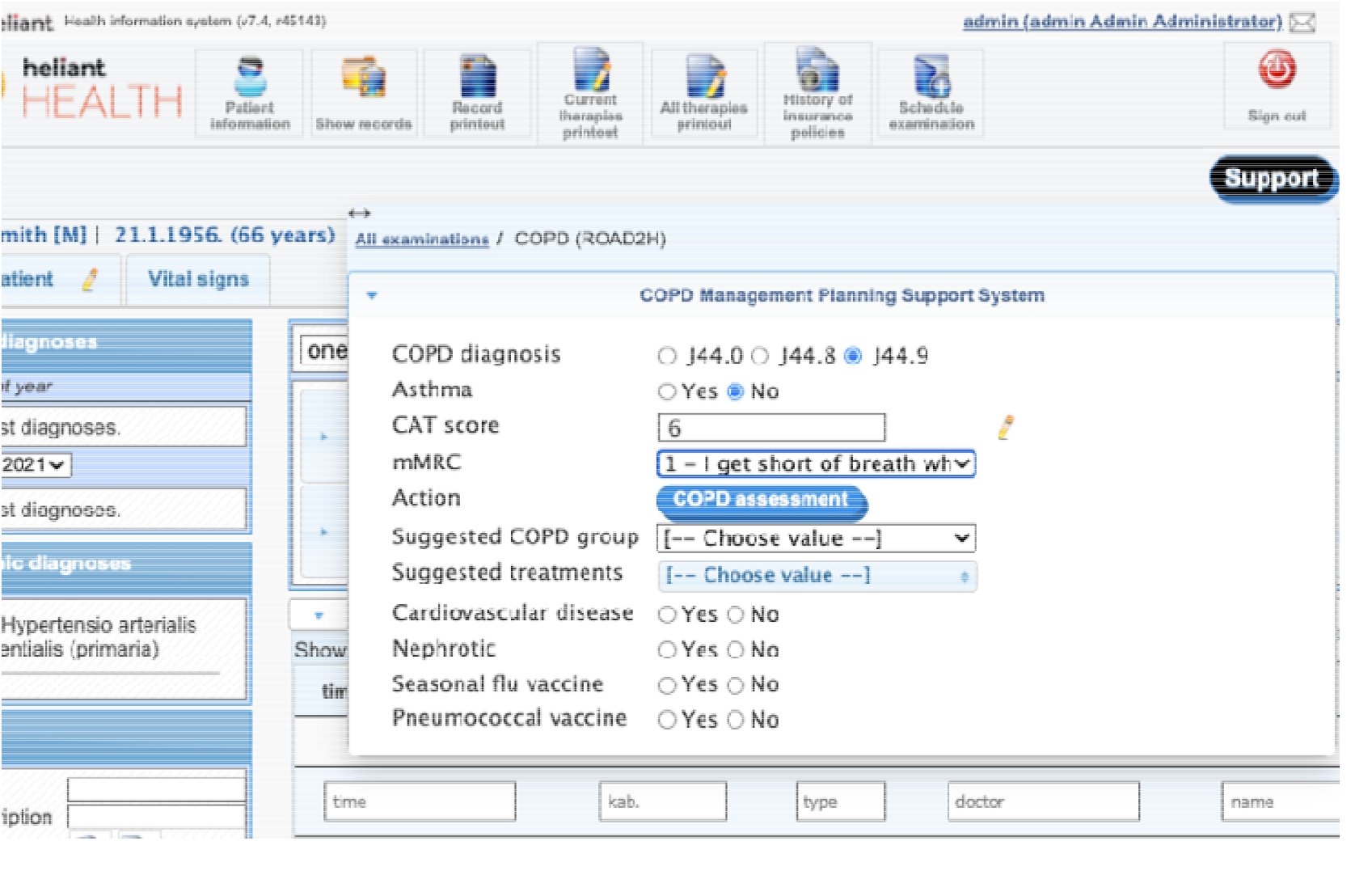
The COPD-CDSS graphical user interface displayed on the COPD tab of Heliant’s EHR. The opened EHR belongs to a male patient with no previous COPD history. The result of his first COPD symptom severity assessment, shown in the interface, is entered by the clinician. The ‘COPD assessment’ button triggers hook ‘copd-assess’. The hook’s response provides content to fields ‘Suggested COPD group’ and ‘Suggested treatments’.

Figure 5 shows the clinical contents of the card responding to hook ‘copd-assess’ integrated with the GUI, i.e., the personalized GOLD group and treatments preference order suggested by the COPD-CDSS. However, no preference ordering is maintained when launching hook ‘copd-careplan-review’ via button ‘Personalised care plan’, i.e., ticked checkboxes linked to field ‘Suggested treatments’ are considered equally preferred. This design choice was made by Heliant to simplify user interaction. Consequently, preferences and goals were left out of the COPD-CDSS evaluation.

**Figure 5.**
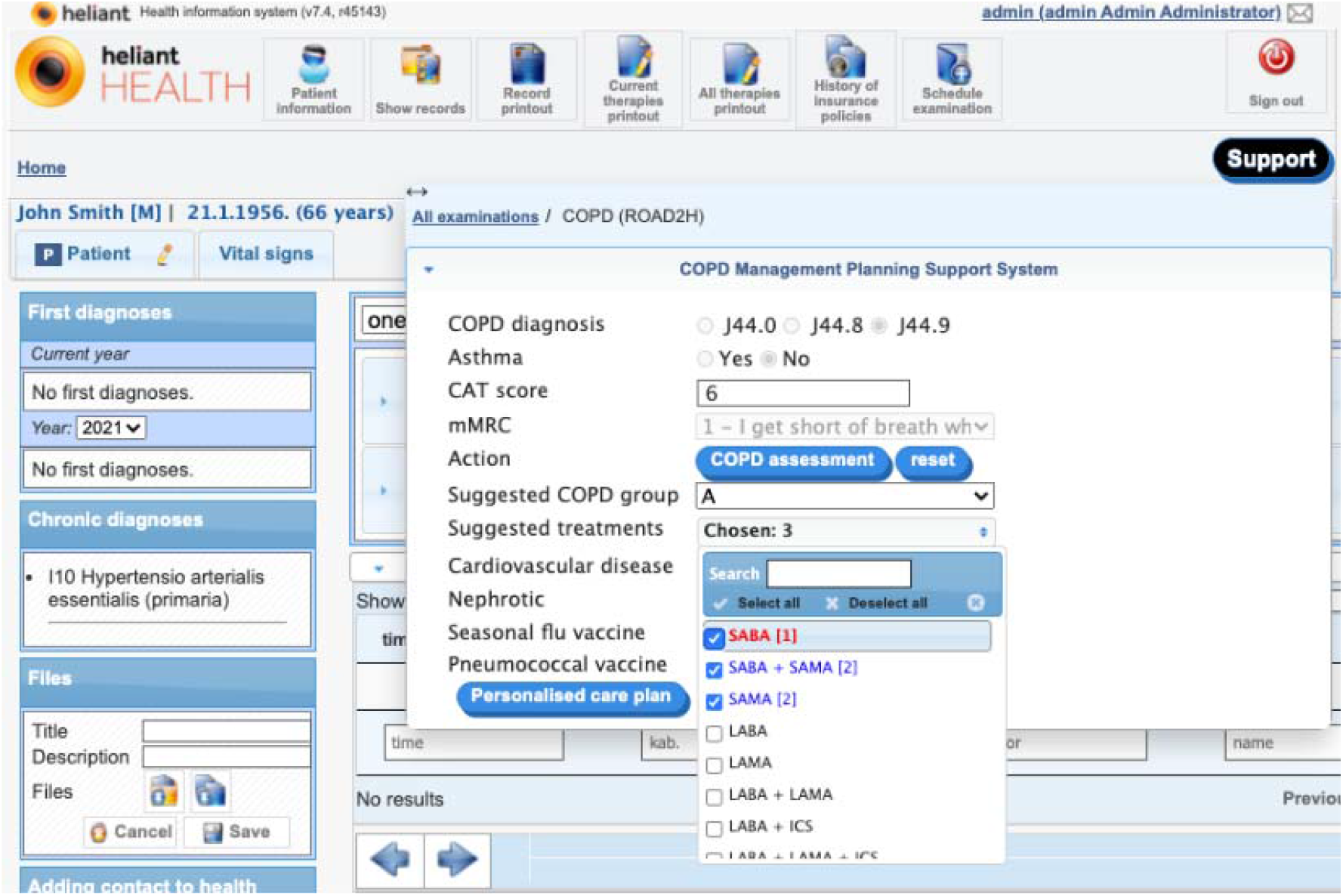
COPD severity assessment response as invoked by the CDS hook ‘copd-assess’ in the COPD-CDSS for the patient introduced in Figure 4.

Figure 6 shows the selection made by the pulmonologist when triggering hook ‘copd-careplan-review’ and the resulting EHR-integrated card. The COPD-CDS response proposed three personalized care plans, the top one has SABA as COPD treatment, which was the sole selection made by the pulmonologist in field ‘Suggested treatments’. The additional proposals provide alternative COPD treatments for the user-selected COPD group as taken from the ‘copd-assess’ response. Additional proposals are added due to detected contradictory recommendations detected involving beta-agonists (see Figure 1). As a beta-agonist, SABA is not recommended for susceptible COPD patients with comorbid CVD.^15^ Application of the SABA-based care plan does not resolve the conflict, some of the other proposals do, so the drug-disease conflict warning was included as an explanation in the proposal with no beta-agonists. Additionally, there are recommendations for pulmonary rehabilitation therapy and to administer the pneumococcal vaccine. The former is common to all COPD patients, the latter is due to the patient’s age (66). The rationale behind each proposed recommendation (Figure 6, right tab) is depicted in a structured form, generated from the encoded terms provided by the explainability algorithm, part of the conflict resolution engine. This design choice was chosen over the generated (English) textual representation (e.g., Table 2, column Explanation) as it enhances scalability and simplifies translation: encoded TMR terms support formal interpretation (e.g., finding SNOMED CT representatives, however mapping back TMR-based knowledge to SNOMED CT was not part of the evaluation). The right tab in Figure 6 enumerates potential interactions found in the SABA-driven care plan (depicted in Figure 1) and how they were resolved (here, by excluding interacting treatments from this proposed care plan). The text utilises a mix of FHIR detectedIssue resources and user-defined (where no FHIR value existed) nomenclature to describe conflicts and their resolutions.

**Figure 6.**
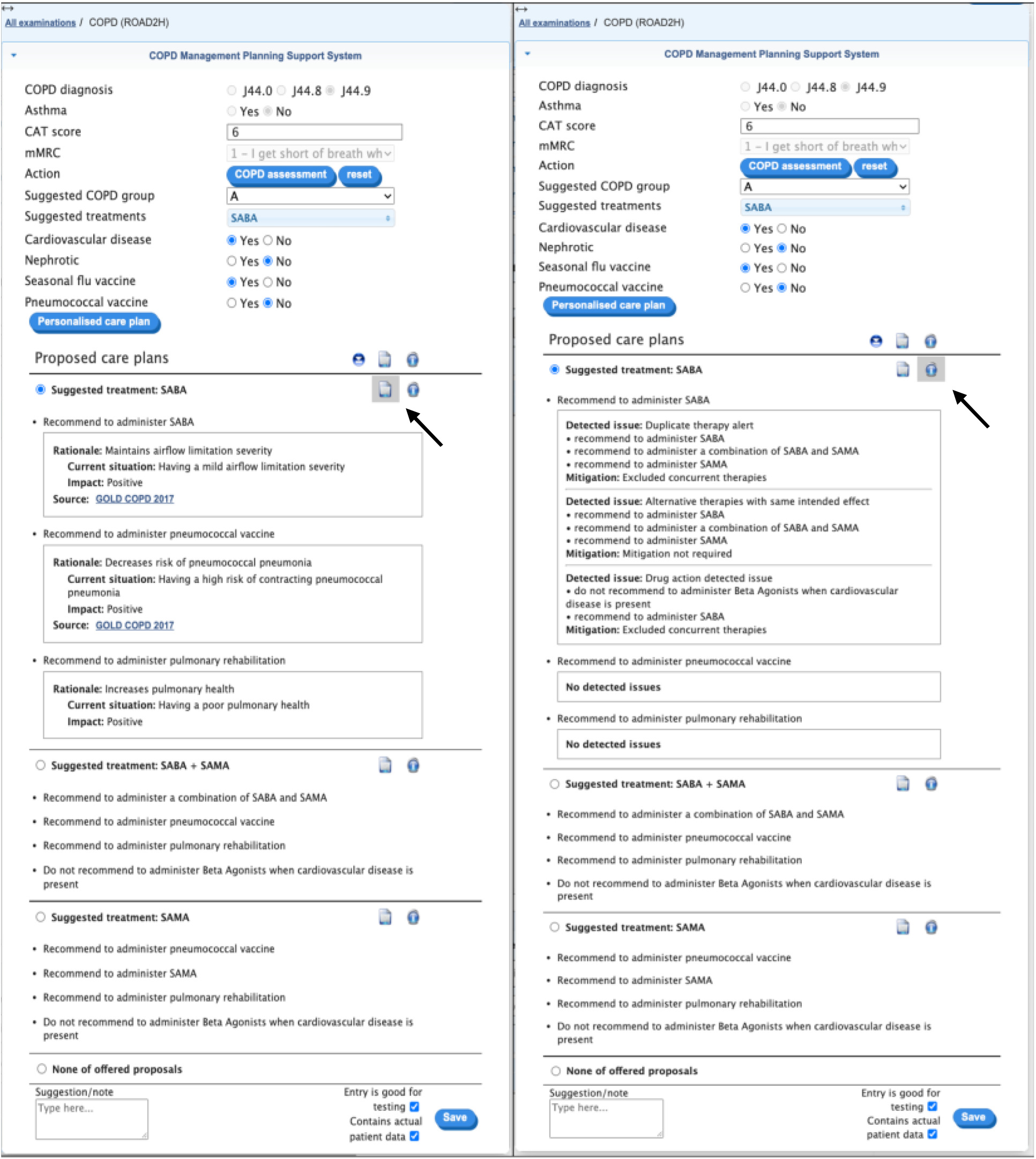
COPD treatment planning response by COPD-CDSS for patient introduced in Figure 4 with COPD assessment from Figure 5. Results are split into two tabs, presented here as one image for the sake of readability. The left tab displays the rationale behind each proposed recommendation, the right tab displays mitigation results for the identified potential interactions among proposed recommendations. A black arrow indicates the button that displays each tab when clicked.

## VI. Evaluation

### A. Agreement with COPD-CDSS results

Data from 99 patient records were returned by Heliant’s EHR. When verifying each case, we found 65 EHRs where entry data did not match that of the provided case vignettes. Many of these modifications were dynamically inserted by pulmonologists at evaluation using the provided GUI, so we deduced they chose to challenge the COPD-CDSS outside the boundaries of the provided cases. However, as we focus on the pulmonologist behaviour for each patient record and associated CDS services, these discrepancies do not affect the overall evaluation. We found two Serbian mistranslations of clinical recommendations that may have left clinicians with a misleading understanding (see Appendix B.2, Table 8). Pulmonologists agreed 97% of the time with the GOLD group assigned by COPD-CDSS to each patient. The remaining 3% found a discrepancy between the use case and the EHR-stored data. Personalised treatments, in the form of COPD drug types and their combinations, were also suggested by the COPD-CDSS when assessing the patient. Suggested treatments were marked with a priority order. Our analysis indicated pulmonologists corroborated the most favoured treatment 31·3% whereas second, third, and fourth choices had 33·3, 24·2, and 9·1 selection rates, respectively. The rejection of all suggested treatments was 2%. These cases corresponded to the previously rejected GOLD groups assessed by the COPD-CDSS.

A selection of alternative care plan proposals that considered the pulmonologists’ treatments preference and the related responses of hook ‘copd-assess’ was presented to the pulmonologists for each use case. Each proposal incorporated personalised recommendations and warnings. Pulmonologists rejected all proposals in the same selection for 1% of the cases while at least one proposal was deemed suitable for 71·7% of the cases. A satisfactory proposal was derived by the pulmonologists for the remaining 27·3% by de-selecting a pair of mistranslated recommendations (English to Serbian) from some results. Incorporating recommendations from alternative proposals was allowed; however, none of the pulmonologists deemed it necessary to augment their selected proposal. Interestingly, clinicians selected the (unordered) topmost displayed proposal for 72% of the cases, the ordering being determined by the operation of the conflict resolution service.

### B. Qualitative analysis

Following use of the COPD-CDSS integrated with Heliant’s EHR and the 20 vignettes, five pulmonologists were invited to take part in individual interviews and responses were documented by the researcher. A structured protocol was used as a guide to explore perceptions in using the COPD-CDSS with case vignettes. For a full list of evaluation questions and answers, see Appendix B.3. The following themes were developed from examining all clinicians’ responses:

#### 1. Treatment and management options

All five pulmonologists agreed the CDS offered a wide range of treatment options. One respondent suggested the CDS offered appropriate treatment choices including alternative therapeutic options.

> *“Offered options for medication, based on the entered parameters on severity and discomfort of disease. Also, the idea of non-medical recommendations*.*” (Doctor 2)*.

One respondent suggested the drug therapies presented could be more precise.

> *“The data must be more precise. E*.*g*., *drugs from the group of beta-agonists are divided into SABA and LABA, and it must be clear to which therapeutic option the data refers to…” (Doctor 1)*

#### 2. Functionality and usability

All five clinicians agreed there were no features of the COPD-CDSS they found unhelpful or ‘least useful’.

The CDS was positively regarded as a *“Clear and concise software” (Doctor 3)*. Although technical issues related to data entry were raised, one respondent stated these were easily resolved.

> *“*… *Minor technical problems (that) were easily resolved in consultations and with the support of the IT specialist” (Doctor 3)*.

One respondent suggested future development of the CDS should aim to be more streamlined.

> *“The goal of every system is simplicity, speed, and efficiency, with as few unnecessary pop-ups and additional questions as possible*.*” (Doctor 5)*.

One way of using the CDS efficiently is integration into existing systems to reduce workload.

> *“Integration with the existing hospital information system, so that all required data for the patient is already present in the EHR system … Avoiding the unnecessary entering of the same data is a waste of time” (Doctor 2)*.

#### 3. Patient monitoring and data capture

The CDS was thought to be useful for continual health monitoring purposes.

> *“In situations when the patient comes regularly for examinations, for better monitoring” (Doctor 4)*.

However, most respondents suggested a wider range of patient data should be captured including co-morbidities, diagnostic investigations, and other therapeutic options.

> *“As much patient data as possible should be entered into the system (from the EHR)” (Doctor 4). “Not enough options for entering the data on associated diseases of importance” (Doctor 1). “Functions of including other aspects (findings) such as spirometry, lab tests, X-rays” (Doctor 3)*.

> *“No therapeutic option for ICS in deciding on a therapeutic option when entering data for patients who also have asthma” (Doctor 3)*.

#### 4. Additional uses of the CDS

One respondent thought the CDS would be an effective companion tool for less experienced clinicians.

> *“The system is a very good idea and a kind of guideline for doctors with little clinical experience at the very beginning of independent management of patients, and then their outpatient examinations*.*” (Doctor 5)*.

Respondents also said that CDS could be used to aid decisions for other diseases.

> *“It can serve as an advising tool and as a reminder of other guidelines if they exist in other diseases” (Doctor 2)*.

> *“It would not be bad to expand the field of action to other specialties (gastroenterology, endocrinology, cardiology*…*) and thus help in deciding the patient with co-morbidities initially during outpatient consultations*.*” (Doctor 5)*.

## VII. Discussion

We successfully extended an ontological framework for expressing guideline recommendations (TMR) to provide individual patient-based reasoning and explanation via argumentation. In addition, a CDS microservices architecture based on open standards (SNOMED CT, HL7 FHIR and CDS Hooks) integrated the resulting TMR-based CDS framework with a commercial EHR. Although the system could manage more than two interactions at a time our evaluation was limited to interactions arising from a single guideline. In clinical practice some patients will have multiple guidelines in operation with potential increased complexity of interactions. Further development of the TMR-based CDS framework to cover multiple guidelines is needed to determine the robustness of the approach. Furthermore, on account of the COVID-19 workload and restrictions in Serbia, the evaluation was delayed for 12 months and was eventually only able to go ahead with five pulmonologists and 20 clinical vignettes rather than live in the COPD clinic as planned. The EHR vendor also introduced several restrictions in that some of the clinical vignette data had to be entered by the clinician at the start, rather than being already in the record, and the ability to order recommendations by prior patient preference and overall clinical goals (such as cost/effectiveness) was omitted. Many of the comments in the qualitative analysis reflect these limitations rather than inherent issues with the approach.

In the practical application of the approach, there are several limitations. Firstly, the representation of the GOLD statements in TMR is time-consuming, requiring input from experienced clinicians and knowledge engineers. This problem is common to all knowledge representation approaches including PROFORMA^26^, Arden Syntax,^27^ and CQL^28^. However, as TMR relationships are defined using the semantic web, this may offer a potential route for semi-automation of the guideline representation process. A combination of natural language processing and expression of found concepts and relationships as knowledge graphs, constrained by the TMR ontology, would be the next research step. In terms of integration with EHRs, FHIR is being increasingly adopted and CDS Hooks is a non-proprietary standard, so the approach we took with Heliant should be widely replicable in other LMIC where open-source EHRs based on standards are of particular value. Guidelines represented in TMR are also a shareable open resource, with potential for local adaptation and optimisation in a transparent way. Although existing syntaxes such as CQL have much greater maturity than TMR, TMR being an ontological approach is much more extensible (as in our addition of argumentation) and a better fit with advances in ontology-based knowledge extraction methods such as neural-symbolic reasoning.^29^

Arguably, the ABA+G explanations delineated above do not make use of the full spectrum of explanation techniques available in argumentation. Nevertheless, together with the explainable nature of argumentation, they are a steppingstone in meeting explainability guidelines for the deployment of AI-assisted systems produced by the UK Information Commissioner’s Office and the Turing Institute. These explanations indicate the reasoning underlying the recommendations in the spirit of Explainable AI methods drawn from argumentative abstractions. As future work, we would explore whether interactive forms of explanations, as in asking questions, naturally supported by argumentation, could be an alternative approach to providing explanation in our context.^30^

## VIII. Conclusion

We approached the problem of supplying explainable AI in the form of a CDSS for guidelines by representing guideline statements and an assessment of the patient’s disease stage using the exemplar of COPD and building argumentation as an added reasoning layer on top of an existing ontological model of guideline recommendations, the TMR model. We proved that it is possible to transact both guideline statements, recommendations and contraindications using this approach and to implement it using a microservice model incorporating widely used standards (SNOMED CT, FHIR API and CDS Hooks). In addition, the system was implemented in integration with a commercial EHR widely used in Serbia and other European LMICs. A mixed-method evaluation using vignettes also showed high agreement with pulmonologists and favourable views on the implementation and the potential of such systems. The aim of providing explainable CDS that integrates in real-time with clinical systems and using a reproducible and non-proprietary system that can be scaled across both clinical problems and systems shows promise.

## Data Availability

All data produced are available online at

https:github/susoDominguez/COPD-CDS.git

