## Appendices for "Development and evaluation of an open-source, standards-based approach to explainable artificial intelligence for managing co-morbidity and clinical guidelines using argumentation techniques and the Transition-based Medical Recommendation model"

#### Appendix A: Background context on the COPD-CDSS

##### A.1 Formalisation of GOLD statements

In the UK, COPD management and control is based on two main clinical guidelines, the NICE guideline for acute and chronic COPD, and the GOLD guideline. The NICE guideline outlines management of the disease in the UK context of the NHS as well as local recommendations for pharmacological and non-pharmacological treatments and therapies. The GOLD guideline is an internationally recognised guideline for COPD management that is used by most other countries where a localised approach is not specified. Both guidelines offer similar advice in most cases, with the same pharmacological treatments being recommended, and only minor differences in management of co-morbidities.

With consideration of both the international aspect of the ROAD2H study and the application of the developed multimorbidity CDS in healthcare settings outside the UK, we have chosen to focus on adapting the recommendations presented in the GOLD guideline. The GOLD guideline offers a comprehensive overview of the entire treatment pathway that a patient would follow throughout the course of the disease, covering both pharmacological, and non-pharmacological treatments as well as, monitoring, follow-up, exacerbation management, and management of COPD in the presence of co-morbidities. As the computerisation formalisation of the entire GOLD guideline fell outside of the scope of this project, we have chosen to focus on the pharmacological treatment of stable COPD using bronchodilators, non-pharmacological treatment through physiotherapy and smoking cessation therapy, management of exacerbation risk through immunisation, and the computerisation formalisation of the COPD treatment algorithms outlined in the GOLD guideline.

The GOLD guideline outlines a severity scale for COPD patients that describe the degree of their airflow limitation using spirometry. Additionally, there is a symptoms assessment and a separate assignment algorithm that determines the severity of the COPD patient’s symptoms and their risk of future exacerbations to classify the patient within one of the four risk groups.

Spirometry is a pulmonary function test that is used to measure the patient’s ability to inhale and exhale air, described as a value of Forced Expiratory Volume in 1 second (FEV1). The GOLD severity scale compares the amount of air a patient can forcibly expel in 1 second to the expected value dependent on the patients age, gender, and height, weight, and ethnicity.

The GOLD ABCD scale assessment tool is based on scores of the Modified Medical Research Council (mMRC) dyspnoea scale, the COPD Assessment Test (CAT), and the recent history of acute exacerbations experienced by the patient within the one year prior to their COPD review. The mMRC dyspnoea scale is a self-rating tool used to measure the effect of breathlessness that a patient experiences in everyday life and has a scale of 0 to 4.

The CAT score is a questionnaire that measures the symptoms burden of a patient’s COPD presented by the severity of their cough, phlegm, chest tightness, breathlessness, confidence, quality of sleep, energy, and the effect the condition has head on everyday activities. The CAT score has a range from 0 to 40.

Using the tests described above, a patient is assigned by the COPD-CDSS into one of the four COPD groups (A – D). In a clinical setting, a patient's assignment of COPD severity group is done at every new call of the decision support system. COPD symptom severity assessment is commonly reviewed, with the assumption that the clinician follows the recommended follow-up scheduling of an appointment every 6 months. As a result, the GOLD group classification, and thus the COPD treatment pathway, of the patient may require an update, following the algorithms mentioned above. Updating both COPD classification and treatment depends on the measurements from the current follow-up visit as well as the data recorded at the previous COPD follow-up appointment. The process of reviewing and (possibly) updating the COPD care plan of a patient has been implemented as part of the CDSS. Thus, a patient may move between groups between appointments, which the COPD-CDSS considers by comparing the newly assigned severity group to the severity group recorded in the EHR.

Next, the COPD-CDSS assigns an appropriate COPD treatment based on the GOLD guideline treatment preference algorithm. The assignment is based on the previous and current COPD severity assessment results, the active COPD treatment (if any) the patient is undertaking, and any relevant co-morbidities that are present.

As part of the GOLD guidelines, a list of recommendations is included for each treatment group that describes the pharmacological treatment. As an example, one of the recommendations listed for group B is as follows:

- For patients with severe breathlessness initial therapy with two bronchodilators may be considered.

For this statement to be formalised as clinical knowledge using the TMR model, the initial response would be to design a pair of TMR recommendations, one per each care action involving a bronchodilator; however, since the therapy involves the combination of both, we define a TMR drug class that represents the combination of both bronchodilators, to then instantiate the class with two drug types corresponding to each of the bronchodilators. Grouping bronchodilators that are part of the same therapy as part of the same drug class is more logical and avoids the detection of spurious interactions by the reasoning engine of TMR (notice that, if split into separate TMR-based recommendations, the TMR reasoning tool would define them as alternative recommendations).

In addition to therapeutical recommendations, we have also considered for formalisation adverse reactions to pharmacological treatments. For instance, as stated in the GOLD guideline:

*“Stimulation of beta2-adrenergic receptors can produce resting sinus tachycardia and has the potential to precipitate cardiac rhythm disturbances in susceptible patients”*

We included this information as a TMR recommendation, in the form of a warning or alert, so that if the patient has a history of cardiac disease, the TMR warning would be triggered by the COPD_CDSS, leading to the reasoning and mitigation services to detect and recommend alternative muscarinic antagonist treatments (whenever possible) over beta agonists.

##### A.2 The ontology- and standards-based CDS microservice architecture

The CDS microservice architecture on which the COPD-CDSS is built aims to enforce a discipline for adding external guideline-based CDS to EHR systems by providing a framework that facilitates interoperability with diverse ontology-based CDS services. Notably, the CDS architecture uses a single-entry point for all CDS that leverages widely used HL7 (FHIR and CDS Hooks) and SNOMED CT interoperability standards. As a result, the microservice architecture standardises the integration of executable CGs into EHR systems, allowing for a ‘write once, share everywhere’ CG portability strategy. Additionally, our approach is non-proprietary and modular, thus lowering maintenance costs as well as allowing for ease of scalability in the number of services offered without affecting those already deployed.

The microservice architecture depicted in Figure 2 combines a fully developed CDS Hooks Manager (CDS-HsM) microservice which operates as the single-entry point between EHR systems and subscribed CDS services; one or more CDS Services Manager (CDS-SsM) microservices -at least one instance per each integrated CG formalism- which are invoked by the CDS-HsM microservice. Each separate CDS-SsM microservice encapsulates the functionality required to enact CGs as required by each available CDS service; lastly, the CGs authoring microservice suite, where each instance, one per integrated CG formalism, is invoked by one or more coupled CDS-SsM microservices. The CGs authoring architecture encapsulates existing CGs technology to store, reason about, and interact with ontology-based clinical knowledge. The CGs authoring microservice suite has been described in detail in Chapman and Curcin,^25^ where a TMR-based implementation of their CGs authoring architecture was also provided, adopting the Apache Jena database (found at <https://jena.apache.org/>) as the store service, queried by SPARQL expressions, and the SWI-Prolog environment as the reasoner service. The TMR-based instance of the CDS-SsM microservice has been previously described in Figure 3. Hence, below, we briefly discuss the functionality of the CDS-HsM microservice.

The CDS-HsM is a middleware service to handle CDS Discovery and CDS Services requests for any integrated CGs authoring microservice suite (e.g., Chapman and Curcin’s TMR-based implementation)^25^ and subscribed EHR systems, via the CDS-SsM microservice. The CDS-HsM engine is agnostic to the semantics of the services invoked by any EHR system. This service is instead tasked with querying, and possibly manipulating, the clinical workflow context, which is part of a CDS request call, following instructions from ‘CDS hooks query e-forms’, collections of JSON-like documents (BSON documents) where each collection is grouped under a hook identifier and stored in a NoSQL database (we use MongoDB, found at <https://www.mongodb.com/>). CDS hooks query e-forms leverage existing CDS hooks specifications, and integrated CGs, to provide data for building CDS. Each hook query e-form is created by a knowledge engineer as a read-only record that is reusable across subscribed heterogeneous EHR systems which are FHIR-compliant. E-forms are loaded when a CDS service is invoked via its corresponding CDS hook. Data query instructions are applicable as JSONata (found at <https://jsonata.org/>) query expressions -a declarative open-source query and transformation language for JSON data-, alongside both internal and user-defined actions (e.g., data comparisons or querying an external SNOMED CT browser, for the former, and calculating the body max index of a patient using FHIR-based representations of height and weight, for the latter). The CDS Hooks Manager microservice is a powerful tool capable of querying simple data such as a patient identifier or more complex algorithms like the representation of the COPD assessment tool from GOLD as designed for the COPD-CDSS.

#### Appendix B: Use Cases Documentation and Evaluation Data

##### B.1 Use case vignettes

Patient 1

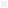

- Age: 62
- Sex: male
- smoking status: current smoker
- GOLD: 2
- Previous therapy: SABA
- Number of exacerbations in past 12 months at last visit: 3 (0 admissions)
- Previous CAT score: 6
- Previous mMRC score: 0
- *Previous COPD group: C*
- Exacerbations in past 12 months at current visit: 2 (0 admissions)
- Current CAT score: 10
- Current mMRC score: 1
- Comorbidities: hypertension
- Asthma: not present
- Flu jab: not-done
- Pneumococcal jab: not-done

Patient 2

- Age: 75
- Sex: male
- smoking status: ex-smoker (quit 2 years ago)
- GOLD: 4
- Previous therapy: LABA + ICS + LAMA
- Number of exacerbations in past 12 months at last visit: 4 (1 admission)
- Previous CAT score: 25
- Previous mMRC score: 3
- *Previous COPD group: D*
- Exacerbations in past 12 months at current visit: 5 (1 admission)
- Current CAT SCORE: 29
- Current mMRC score: 3
- Comorbidities: coronary heart disease, TIA (cerebrovascular disease), chronic kidney disease stage 3, osteoarthritis
- Asthma: not present
- Flu jab: completed
- Pneumococcal jab: completed

Patient 3

- Age: 64
- Sex: female
- smoking status: ex-smoker (quit 6 years ago)
- GOLD: 3
- Previous therapy: LABA
- Number of exacerbations in past 12 months at last visit: 1 (0 admission)
- Previous CAT score: 17
- Previous mMRC score: 1
- *Previous COPD group: B*
- Exacerbations in past 12 months at current visit: 4 (2 admissions)
- Current CAT score: 20
- Current mMRC score: 2
- Comorbidities: diabetes mellitus
- Asthma: not present
- Flu jab: not-done
- Pneumococcal jab: not-done

Patient 4

- Age: 50
- Sex: female
- smoking status: current smoker
- GOLD: 2
- Previous therapy: SABA
- Number of exacerbations in past 12 months at last visit: 1 (0 admissions)
- Previous CAT score: 8
- Previous mMRC score: 1
- *Previous COPD group: A*
- Exacerbations in past 12 months at current visit: 3 (0 admissions)
- Current CAT score: 9
- Current mMRC score: 1
- Comorbidities: rheumatoid arthritis
- Asthma: present
- Flu jab: not-done
- Pneumococcal jab: not-done

Patient 5

- Age: 62
- Sex: female
- smoking status: current smoker
- GOLD: 2
- Previous therapy: LAMA + SABA
- Number of exacerbations in past 12 months at last visit: 1 (0 admissions)
- Previous CAT score: 18
- Previous mMRC score: 2
- *Previous COPD group: B*
- Exacerbations in past 12 months at current visit: 3 (0 admissions)
- Current CAT score: 26
- Current mMRC score: 3
- Comorbidities: coronary heart disease, obesity, hypertension, obstructive sleep apnoea
- Asthma: not present
- Flu jab: completed
- Pneumococcal jab: completed

Patient 6

- Age: 57
- Sex: male
- smoking status: current smoker
- GOLD: 2
- Previous therapy: SABA + ICS
- Number of exacerbations in past 12 months at last visit: 0 (0 admissions)
- Previous CAT score: 6
- Previous mMRC score: 1
- *Previous COPD group: A*
- Exacerbations in past 12 months at current visit: 0 (0 admissions)
- Current CAT score: 11
- Current mMRC score: 1
- Comorbidities: depression
- Asthma: present
- Flu jab: not-done
- Pneumococcal jab: not-done

Patient 7

- Age: 68
- Sex: male
- smoking status: ex-smoker (quit 1 year ago)
- GOLD: 2
- Previous therapy: LAMA
- Number of exacerbations in past 12 months at last visit: 2 (0 admissions)
- Previous CAT score: 9
- Previous mMRC score: 1
- *Previous COPD group: C*
- Exacerbations in past 12 months at current visit: 4 (3 admissions)
- Current CAT score: 16
- Current mMRC score: 1
- Comorbidities: hypertension, hyperlipidaemia, atrial fibrillation, chronic kidney disease stage 3
- Asthma: not present
- Flu jab: not-done
- Pneumococcal jab: not-done

Patient 8

- Age: 78
- Sex: male
- smoking status: ex-smoker (quit 10 years ago)
- GOLD: 3
- Previous therapy: LAMA
- Number of exacerbations in past 12 months at last visit: 1 (0 admission)
- Previous CAT score: 25
- Previous mMRC score: 3
- *Previous COPD group: B*
- Exacerbations in past 12 months at current visit: 2 (1 admission)
- Current CAT score: 27
- Current mMRC score: 4
- Comorbidities: previous myocardial infarction, heart failure, hypertension, prostate cancer
- Asthma: not present
- Flu jab: completed
- Pneumococcal jab: not-done

Patient 9

- Age: 69
- Sex: female
- smoking status: current smoker
- GOLD: 2
- Previous therapy: LABA + ICS
- Number of exacerbations in past 12 months at last visit: 0 (0 admissions)
- Previous CAT score: 22
- Previous mMRC score: 1
- *Previous COPD group: B*
- Exacerbations in past 12 months at current visit: 0 (0 admissions)
- Current CAT score: 20
- Current mMRC score: 1
- Comorbidities: anxiety, hypothyroidism
- Asthma: not present
- Flu jab: not-done
- Pneumococcal jab: not-done

Patient 10

- Age: 65
- Sex: male
- smoking status: ex-smoker (quit 5 years ago)
- GOLD: 1
- Previous therapy: none
- Number of exacerbations in past 12 months at last visit: ·· (first visit)
- Previous CAT score: none
- Previous mMRC score: none
- *Previous COPD group: ··*
- Exacerbations in past 12 months at current visit: 0 (0 admissions)
- Current CAT score: 12
- Current mMRC score: 1
- Comorbidities: hypertension, hypercholesterolaemia, angina (coronary heart disease)
- Asthma: not present
- Flu jab: not-done
- Pneumococcal jab: not-done

Patient 11

- Age: 71
- Sex: female
- smoking status: current smoker
- GOLD: 3
- Previous therapy: LAMA + LABA
- Number of exacerbations in past 12 months at last visit: 2 (0 admissions)
- Previous CAT score: 9
- Previous mMRC score: 1
- *Previous COPD group: C*
- Exacerbations in past 12 months at current visit: 1 (0 admissions)
- Current CAT score: 26
- Current mMRC score: 2
- Comorbidities: previous CVA (cerebrovascular disease), hypertension, osteoporosis, anaemia, depression, chronic kidney disease stage 3
- Asthma: not present
- Flu jab: completed
- Pneumococcal jab: not-done

Patient 12

- Age: 76
- Sex: male
- smoking status: ex-smoker (quit 3 years ago)
- GOLD: 3
- Previous therapy: SAMA
- Number of exacerbations in past 12 months at last visit: 1 (0 admission)
- Previous CAT score: 5
- Previous mMRC score: 1
- *Previous COPD group: A*
- Exacerbations in past 12 months at current visit: 1 (1 admission)
- Current CAT score: 8
- Current mMRC score: 2
- Comorbidities: diabetes mellitus, previous CVA (cerebrovascular disease), hypertension, chronic kidney disease stage 4
- Asthma: not present
- Flu jab: completed
- Pneumococcal jab: not-done

Patient 13

- Age: 42
- Sex: female
- smoking status: current smoker
- GOLD: 2
- Previous therapy: none
- Number of exacerbations in past 12 months at last visit: ·· (first visit)
- Previous CAT score: none
- Previous mMRC score: none
- *Previous COPD group: ··*
- Exacerbations in past 12 months at current visit: 1 (0 admissions)
- Current CAT score: 6
- Current mMRC score: 1
- Comorbidities: none
- Asthma: present
- Flu jab: not-done
- Pneumococcal jab: not-done

Patient 14

- Age: 59
- Sex: female
- smoking status: current smoker
- GOLD: 2
- Previous therapy: LABA + SABA
- Number of exacerbations in past 12 months at last visit: 1 (0 admission)
- Previous CAT score: 20
- Previous mMRC score: 1
- *Previous COPD group: B*
- Exacerbations in past 12 months at current visit: 3 (0 admission)
- Current CAT score: 21
- Current mMRC score: 2
- Comorbidities: obesity, hypercholesterolaemia, peripheral vascular disease
- Asthma: not present
- Flu jab: not-done
- Pneumococcal jab: not-done

Patient 15

- Age: 61
- Sex: male
- smoking status: ex-smoker (quit 4 years ago)
- GOLD: 2
- Previous therapy: SABA
- Number of exacerbations in past 12 months at last visit: 0 (0 admission)
- Previous CAT score: 9
- Previous mMRC score: 1
- *Previous COPD group: A*
- Exacerbations in past 12 months at current visit: 0 (0 admission)
- Current CAT score: 14
- Current mMRC score: 2
- Comorbidities: diabetes mellitus, previous community acquired pneumonia
- Asthma: not present
- Flu jab: completed
- Pneumococcal jab: not-done

Patient 16

- Age: 46
- Sex: male
- smoking status: ex-smoker (quit 1 year ago)
- GOLD: 1
- Previous therapy: LABA + ICS
- Number of exacerbations in past 12 months at last visit: 0 (0 admissions)
- Previous CAT score: 8
- Previous mMRC score: 0
- *Previous COPD group: A*
- Exacerbations in past 12 months at current visit: 3 (0 admissions)
- Current CAT score: 9
- Current mMRC score: 1
- Comorbidities: allergic rhinitis
- Asthma: present
- Flu jab: not-done
- Pneumococcal jab: not-done

Patient 17

- Age: 77
- Sex: female
- smoking status: current smoker
- GOLD: 3
- Previous therapy: LAMA + LABA
- Number of exacerbations in past 12 months at last visit: 2 (0 admissions)
- Previous CAT score: 27
- Previous mMRC score: 2
- *Previous COPD group: D*
- Exacerbations in past 12 months at current visit: 4 (1 admission)
- Current CAT score: 26
- Current mMRC score: 3
- Comorbidities: previous breast cancer, chronic kidney disease stage 4
- Asthma: not present
- Flu jab: completed
- Pneumococcal jab: completed

Patient 18

- Age: 62
- Sex: female
- smoking status: ex-smoker (quit 6 months ago)
- GOLD: 3
- Previous therapy: LABA + ICS
- Number of exacerbations in past 12 months at last visit: 2 (0 admissions)
- Previous CAT score: 6
- Previous mMRC score: 1
- *Previous COPD group: C*
- Exacerbations in past 12 months at current visit: 4 (0 admissions)
- Current CAT score: 7
- Current mMRC score: 1
- Comorbidities: osteoporosis
- Asthma: not present
- Flu jab: not-done
- Pneumococcal jab: not-done

Patient 19

- Age: 58
- Sex: male
- smoking status: ex-smoker (quit 2 months ago)
- GOLD: 4
- Previous therapy: LABA + ICS + LAMA
- Number of exacerbations in past 12 months at last visit: 4 (2 admissions)
- Previous CAT score: 32
- Previous mMRC score: 2
- *Previous COPD group: D*
- Exacerbations in past 12 months at current visit: 6 (5 admissions)
- Current CAT score: 37
- Current mMRC score: 3
- Comorbidities: hypertension, ischaemic heart disease, diabetes mellitus, chronic kidney disease stage 3, sciatica
- Asthma: not present
- Flu jab: completed
- Pneumococcal jab: completed

Patient 20

- Age: 55
- Sex: male
- smoking status: current smoker
- GOLD: 2
- Previous therapy: SAMA
- Number of exacerbations in past 12 months at last visit: 1 (0 admissions)
- Previous CAT score: 7
- Previous mMRC score: 0
- *Previous COPD group: A*
- Exacerbations in past 12 months at current visit: 2 (0 admissions)
- Current CAT score: 6
- Current mMRC score: 1
- Comorbidities: hypertension, hyperlipidaemia, previous myocardial infarction
- Asthma: present
- Flu jab: not-done
- Pneumococcal jab: not-done

##### B.2 Evaluation data

We present the evaluation data collected from the application of the COPD-CDSS to the use cases detailed above by each of the five Serbian pulmonologists. Prior to that, we introduce in Table 8 the textual representation of the selected GOLD statements, both in English and Serbian. Additionally, in Table 9 we provide the semantics of the headers for each column in the evaluation data. Both tables are to be used as support for the reader when reviewing the collected evaluation data.

Table 8. **TMR-based care action recommendations representing selected GOLD statements**. To be used as part of the dictionary for the COPD-CDSS evaluation data shown below. Observe how the Serbian translations (column English translation of Serbian textual display for non-Serbian readers) with URI references BetaAgonists and LabaLamaCkd do not mention the COPD co-morbidities stated in their English counterparts.

| **Care action URI reference** | **English textual display** | **Serbian textual display** | **English translation of Serbian textual display** |
| --- | --- | --- | --- |
| Gluccorticds | do not recommend to administer inhaled corticosteroids | Ne preporučuje se primena inhalacije kortikosteroidima | Corticosteroid inhalation is not recommended |
| PneumnVac | recommend to administer pneumococcal vaccine | Preporučuje se primena pneumokokne vakcine | The use of pneumococcal vaccine is recommended |
| FluVac | recommend to administer influenza vaccine | Preporučuje se primena vakcine protiv gripa | It is recommended to use the flu vaccine |
| SmokingTher | recommend to administer smoking cessation therapy | Preporučuje se primena terapije za prestanak pušenja | Smoking cessation therapy is recommended |
| BetaAgonists | do not recommend to administer Beta Agonists when cardiovascular disease is present | Ne preporučuje se primena bronhodilatatora iz grupe beta agonista | The use of bronchodilators from the group of beta agonists is not recommended |
| PulmonRehab | recommend to administer pulmonary rehabilitation | Preporučuje se sprovođenje plućne rehabilitacije | Pulmonary rehabilitation is recommended |
| LabaLamaCkd | do not recommend to administer LABA or LAMA when renall or nephrotic disease is present | Ne preporučuje se primena LABA i LAMA bronhodilatatora | The use of LABA and LAMA bronchodilators is not recommended |
| Saba | recommend to administer SABA | Preporučuje se primena SABA bronhodilatatora | The use of SABA is recommended |
| Sama | recommend to administer SAMA | Preporučuje se primena SAMA bronhodilatatora | The use of SAMA is recommended |
| SabaSama | recommend to administer a combination of SABA and SAMA | Preporučuje se primena SABA i SAMA bronhodilatatora | The use of SABA and SAMA is recommended |
| Laba | recommend to administer LABA |  | The use of LABA is recommended |
| Lama | recommend to administer LAMA | Preporučuje se primena LAMA bronhodilatatora | The use of LAMA is recommended |
| LabaLama | recommend to administer a combination of LABA and LAMA |  | The use of LABA and LAMA is recommended |
| LabaIcs | recommend to administer a combination of LABA and ICS | Preporučuje se primena LABA i ICS | The use of LABA and ICS is recommended |
| LabaLamaIcs | recommend to administer a combination of LABA, LAMA and ICS | Preporučuje se primena LABA i LAMA i ICS | The use of LABA and LAMA and ICS is recommended |

Table 9. **Semantics of the headers in the COPD-CDSS evaluation data**. Column keyword identifies each of the column labels that are headers in the evaluation data tables below. Column semantics describes the meaning of each column label. To be used as part of the dictionary for the COPD-CDSS evaluation data shown below.

| **Keyword** | **Semantics** |
| --- | --- |
| ID | Use case identifier |
| previous CAT score | CAT score recorded at previous COPD review |
| previous mMRC score | mMRC dyspnoea scale score recorded at previous COPD review |
| previous number of exacerbations | recorded number of exacerbations within one year prior to the previous COPD review |
| active COPD treatment | currently active COPD drug type, or drug class (i.e., combination of COPD drug types) |
| current CAT score | CAT score measured at current COPD review |
| current mMRC score | mMRC dyspnoea scale score measured at current COPD review |
| Current number of exacerbations | recorded number of exacerbations within one year prior to the current COPD review |
| Has asthma? | is asthma present in EHR? |
| previous GOLD group | GOLD COPD group recorded at previous COPD review |
| copd-assess result: GOLD group | GOLD COPD group as suggested by COPD-CDSS and based on GOLD guideline |
| user-selected GOLD group | GOLD COPD group related to the patient’s COPD symptom severity as selected by the pulmonologist |
| copd-assess result: personalized COPD treatments | personalised COPD treatment preference list as suggested by COPD-CDSS, and based on GOLD guideline, via ‘copd-assess’ CDS service taking into account the pulmonologist’s GOLD group selection |
| had annual influenza vaccine? |  |
| had annual pneumococcal vaccine? | is there a completed pneumococcal vaccination recorded in the EHR? |
| Age | patient's current age |
| is a smoker? | is the patient currently a smoker? |
| Has CKD? | is chronic kidney disease a recorded condition in the EHR of the patient? |
| Has CVD? | is cardiovascular disease a recorded condition in the EHR of the patient? |
| user-selected treatments | Selection by pulmonologist to design personalised COPD care plan |
| COPD care plan proposal 1— 4 | personalised COPD care plans as proposed by CDS system and ordered as displayed by Heliant's graphical user interface (no priority order) |
| Selected proposal/COPD treatment | COPD care plan proposal selected by pulmonologist as most suitable for current use case (if any)/COPD treatment suggested as part of this care plan proposal |
| Context-suitable recs | Recommendations selected by the pulmonologist as suitable to manage the COPD of the current patient |

##### Pulmonologist 1: Evaluation data

Table 10. **Use cases clinical context as collected from the EHR of pulmonologist 1**. Cell data with format m(n) where m,n are numbers denotes contextual use case data entered incorrectly into the EHR (represented outside the brackets) beside the contextual use case data given in the documentation (represented inside the brackets). Keyword MISSING denotes the data was not entered into the EHR.

| **Use case ID** | **previous CAT score** | **previous mMRC score** | **previous number of exacerbations** | **active COPD treatment** | **current CAT score** | **current mMRC score** | **Current number of Exacerbations** | **Has asthma?** | **previous GOLD group** |
| --- | --- | --- | --- | --- | --- | --- | --- | --- | --- |
| 1 | 10 (6) | 1 (0) | 5 (3) | Saba | 10 | 1 | 0 (2) | FALSE | B (C) |
| 2 | 29 (25) | 3 | 9 (4) | Lama (Laba+Lama+Ics) | 29 | 3 | 0 (5) | FALSE | B (D) |
| 3 | 20 (17) | 2 (1) | 5 (1) | Laba | 20 | 1 (2) | 0 (4) | FALSE | B |
| 4 | 9 (8) | 1 | 5 (1) | Saba | 9 | 1 | 0 (3) | TRUE | C (A) |
| 5 | 20 (18) | 1 (2) | 0 (1) | Saba (Lama) | 26 | 2 (3) | 0 (3) | FALSE | A (B) |
| 6 | 11 (6) | 1 (1) | 0 | Saba | 11 | 1 | 0 | TRUE | A |
| 7 | 16 (9) | 1 (1) | 6 (2) | Lama | 16 | 1 | 0 (4) | FALSE | D (C) |
| 8 | 27 (25) | 4 (3) | 3 (1) | Lama | 27 | 4 | 0 (2) | FALSE | D |
| 9 | 20 (22) | 1 | 0 | Saba (Laba+Ics) | 20 | 1 | 0 | FALSE | A (B) |
| 10 | ·· | ·· | 0 | ·· | 12 | 1 | 0 | FALSE | ·· |
| 11 | 26 (9) | 2 (1) | 0 (2) | Saba (Laba+Lama) | 26 | 2 | 0 (1) | FALSE | A (C) |
| 12 | 5 | 1 | 2 (1) | Saba+Sama (Sama) | 8 | 2 | 0 (1) | FALSE | C (A) |
| 13 | 6 | 1 | 0 | ·· | 6 | 1 | 0 (1) | TRUE | ·· |
| 14 | 21 (20) | 2 (1) | 0 (1) | Saba (Laba) | 21 | 2 | 0 (3) | FALSE | A (B) |
| 15 | 14 (9) | 2 (1) | 0 | Saba | 14 | 2 | 0 | FALSE | B (A) |
| 16 | 9 (8) | 0 | 0 | Laba+Ics | 9 | 1 | 3 | TRUE | A |
| 17 | 27 | 2 | 0 (2) | Laba+Lama | 26 | 3 | 6 (4) | FALSE | D |
| 18 | 7 (6) | 1 | 0 (2) | Saba (Laba+Ics) | 7 | 1 | 0 (4) | FALSE | A (C) |
| 19 | MISSING | MISSING | MISSING | MISSING | MISSING | MISSING | MISSING | MISSING | MISSING |
| 20 | 6 (7) | 1 (0) | 3 (1) | Sama | 7 (6) | 0 (1) | 0 (2) | TRUE | C (A) |

Table 11. **Results from CDS service ‘copd-assess’ and input for CDS service ‘copd-careplan-review’, as collected from both the EHR of pulmonologist 1 and COPD-CDSS logs**. Keyword MISSING denotes the contextual data was not entered into the EHR.

| **use case ID** | **copd-assess result: GOLD group** | **user-selected GOLD group** | **copd-assess result: personalized COPD treatments** | **had annual influenza vaccine?** | **had pneumococcal vaccine?** | **age** | **is a smoker?** | **Has CKD?** | **history of CVD?** |
| --- | --- | --- | --- | --- | --- | --- | --- | --- | --- |
| 1 | B | B | [[Laba],[Lama],[Laba+Lama]] | FALSE | FALSE | 62 | TRUE | FALSE | TRUE |
| 2 | B | D | [ [Lama], [Laba, Laba+Lama] ] | TRUE | TRUE | 76 (75) | FALSE | FALSE | TRUE |
| 3 | B | B | [ [Laba],[Lama, Laba+Lama] ] | FALSE | FALSE | 64 | FALSE | FALSE | FALSE |
| 4 | A | A | [ [Saba], [Saba+Sama, Sama] ] | FALSE | FALSE | 50 | TRUE | FALSE | FALSE |
| 5 | D | D | [ [Laba+Lama], [Laba+Lama+Ics], [Laba+Ics], [Lama] ] | TRUE | TRUE | 69 (62) | TRUE | FALSE | TRUE |
| 6 | B | B | [[Laba],[Lama],[Laba+Lama]] | FALSE | FALSE | 57 | TRUE | FALSE | FALSE |
| 7 | B | B | [[Lama],[Laba, Laba+Lama]] | FALSE | FALSE | 68 | FALSE | FALSE (TRUE) | TRUE (FALSE) |
| 8 | B | B | [[Lama],[Laba, Laba+Lama]] | TRUE | FALSE | 78 | FALSE | FALSE | TRUE |
| 9 | B | B | [[Laba],[Lama],[Laba+Lama]] | FALSE | FALSE | 69 | TRUE | FALSE | FALSE |
| 10 | B | B | [[Laba],[Lama],[Laba+Lama]] | FALSE | FALSE | 65 | FALSE | FALSE | FALSE (TRUE) |
| 11 | B | B | [[Laba],[Lama],[Laba+Lama]] | TRUE | FALSE | 71 | TRUE | FALSE (TRUE) | TRUE |
| 12 | A | A | [[Saba+Sama],[Sama,Saba]] | TRUE | FALSE | 76 | FALSE | TRUE | TRUE (FALSE) |
| 13 | A | A | [[Saba],[Saba+Sama,Sama]] | FALSE | FALSE | 42 | TRUE | FALSE | FALSE |
| 14 | B | B | [ [Laba], [Lama], [Laba+Lama] ] | FALSE | FALSE | 59 | TRUE | FALSE | FALSE |
| 15 | B | B | [ [Laba], [Lama], [Laba+Lama] ] | TRUE | FALSE | 61 | FALSE | FALSE | FALSE |
| 16 | C | C | [ [Laba+Ics], [Laba+Lama], [Lama]] | FALSE | FALSE | 46 | FALSE | FALSE | FALSE |
| 17 | D | D | [ [Laba+Lama+Ics], [Laba+Ics], [Laba+Lama], [Lama]] | TRUE | TRUE | 77 | TRUE | TRUE | FALSE |
| 18 | C | C | [ [Lama], [Laba+Lama, Laba+Ics] ] | FALSE | FALSE | 62 | FALSE | FALSE | FALSE |
| 19 | MISSING | MISSING | MISSING | MISSING | MISSING | MISSING | MISSING | MISSING | MISSING |
| 20 | A | A | [ [Sama], [Saba+Sama, Saba]] | FALSE | FALSE | 55 | TRUE | FALSE | TRUE |

Table 12. **User-selected treatment(s) for CDS and corresponding results, with pulmonologist comments, for CDS service ‘copd-careplan-review’ and pulmonologist 1.**

| **use case ID** | **user-selected treatments** | **COPD care plan proposal 1** | **COPD care plan proposal 2** | **COPD care plan proposal 3** | **COPD care plan proposal 4** | **User-selected proposal/COPD treatment** | **Context-suitable recommendations** | **Pulmonologist comments** |
| --- | --- | --- | --- | --- | --- | --- | --- | --- |
| 1 | ALL CDS-SUGGESTED | LabaLama,PulmonRehab,SmokingTher,FluVac,Gluccorticds,BetaAgonists | Lama,PulmonRehab,SmokingTher,FluVac,Gluccorticds,BetaAgonists | Laba,PulmonRehab,SmokingTher,FluVac,Gluccorticds | ·· | 1/Laba+Lama | ALL PROPOSED | ·· |
| 2 | Laba+Lama, Laba+Lama+Ics, Laba+Ics, Lama | Laba,PulmonRehab,Gluccorticds | Lama,PulmonRehab,Gluccorticds,BetaAgonists | LabaLama,PulmonRehab,Gluccorticds,BetaAgonists | ·· | NONE | LabaLamaIcs, PulmonRehab | ·· |
| 3 | ALL CDS-SUGGESTED | Laba,PulmonRehab,FluVac,Gluccorticds | Lama,PulmonRehab,FluVac,Gluccorticds | LabaLama,PulmonRehab,FluVac,Gluccorticds | ·· | 3/Laba+Lama | ALL PROPOSED | ·· |
| 4 | ALL CDS-SUGGESTED | Saba,PulmonRehab,FluVac,SmokingTher | SabaSama,PulmonRehab,FluVac,SmokingTher | Sama,PulmonRehab,FluVac,SmokingTher | ·· | 2/Saba+Sama | ALL PROPOSED | Ova pacijentkinja ima i astmu te je neophodna terapija IKS (This patient also has asthma and needs Ics therapy) |
| 5 | ALL CDS-SUGGESTED | LabaLama,PulmonRehab,SmokingTher,Gluccorticds,BetaAgonists | Lama,PulmonRehab,SmokingTher,Gluccorticds,BetaAgonists | Laba,PulmonRehab,SmokingTher,Gluccorticds | ·· | 1/Laba+Lama | ALL PROPOSED | ·· |
| 6 | ALL CDS-SUGGESTED | Lama,PulmonRehab,SmokingTher,Gluccorticds, FluVac | LabaLama,PulmonRehab,SmokingTher,Gluccorticds, FluVac | Laba,PulmonRehab,SmokingTher,Gluccorticds, FluVac | ·· | 2/Laba+Lama | LabaLama,PulmonRehab,SmokingTher, FluVac | Ova pacijentkinja ima i astmu te je neophodna terapija IKS (This patient also has asthma and needs Ics therapy) |
| 7 | ALL CDS-SUGGESTED | Lama,PulmonRehab,Gluccorticds, FluVac,BetaAgonists,PneumnVac | Laba,PulmonRehab,Gluccorticds, FluVac,PneumnVac | LabaLama,PulmonRehab,Gluccorticds, FluVac,BetaAgonists,PneumnVac | ·· | 3/Laba+Lama | ALL PROPOSED | ·· |
| 8 | ALL CDS-SUGGESTED | Laba,PulmonRehab,Gluccorticds,PneumnVac | Lama,PulmonRehab,Gluccorticds,PneumnVac, BetaAgonists | LabaLama,PulmonRehab,Gluccorticds,BetaAgonists,PneumnVac | ·· | 3/Laba+Lama | ALL PROPOSED | ·· |
| 9 | ALL CDS-SUGGESTED | Lama,PulmonRehab,SmokingTher,FluVac,Gluccorticds,PneumnVac | LabaLama,PulmonRehab,SmokingTher,FluVac,Gluccorticds,PneumnVac | Laba,PulmonRehab,SmokingTher,FluVac,Gluccorticds,PneumnVac | ·· | 1/Lama | ALL PROPOSED | ·· |
| 10 | ALL CDS-SUGGESTED | LabaLama,PulmonRehab,PneumnVac,FluVac,Gluccorticds | Laba,PulmonRehab,PneumnVac,FluVac,Gluccorticds | Lama,PulmonRehab,PneumnVac,FluVac,Gluccorticds | ·· | 1/Laba+Lama | ALL PROPOSED | ·· |
| 11 | ALL CDS-SUGGESTED | Lama,PulmonRehab,SmokingTher,PneumnVac,Gluccorticds,BetaAgonists | LabaLama,PulmonRehab,SmokingTher,PneumnVac,Gluccorticds,BetaAgonists | Laba,PulmonRehab,SmokingTher,PneumnVac,Gluccorticds | ·· | 1/Laba+Lama | ALL PROPOSED | ·· |
| 12 | ALL CDS-SUGGESTED | SabaSama,PulmonRehab,PneumnVac,LabaLamaCkd,BetaAgonists | Saba,PulmonRehab,PneumnVac,LabaLamaCkd | Sama,PulmonRehab,PneumnVac,LabaLamaCkd,BetaAgonists | ·· | NONE | LabaLama,PulmonRehab, PneumnVac,LabaLamaCkd | ·· |
| 13 | ALL CDS-SUGGESTED | Saba,PulmonRehab,SmokingTher,FluVac | SabaSama,PulmonRehab,SmokingTher,FluVac | Sama,PulmonRehab,SmokingTher,FluVac | ·· | 1/Saba | ALL PROPOSED | ·· |
| 14 | ALL CDS-SUGGESTED | Lama,PulmonRehab,SmokingTher,FluVac,Gluccorticds | LabaLama,PulmonRehab,SmokingTher,FluVac,Gluccorticds | Laba,PulmonRehab,SmokingTher,FluVac,Gluccorticds | ·· | 2/Laba+Lama | ALL PROPOSED | ·· |
| 15 | ALL CDS-SUGGESTED | Laba,PulmonRehab,Gluccorticds | Lama,PulmonRehab,Gluccorticds | LabaLama,PulmonRehab,Gluccorticds | ·· | 2/Lama | ALL PROPOSED | ·· |
| 16 | ALL CDS-SUGGESTED | Lama,PulmonRehab,Gluccorticds,FluVac | LabaLama,PulmonRehab,Gluccorticds,FluVac | LabaIcs,PulmonRehab,FluVac | ·· | 3/Laba+Ics | ALL PROPOSED | ·· |
| 17 | ALL CDS-SUGGESTED | LabaLama,PulmonRehab,Gluccorticds,SmokingTher,LabaLamaCkd | Lama,PulmonRehab,Gluccorticds,SmokingTher | LabaLamaIcs,PulmonRehab,SmokingTher,LabaLamaCkd | LabaIcs,PulmonRehab,SmokingTher,LabaLamaCkd | 3/Laba+Lama+Ics | ALL PROPOSED | ·· |
| 18 | ALL CDS-SUGGESTED | Lama,PulmonRehab,Gluccorticds,FluVac | LabaLama,PulmonRehab,Gluccorticds,FluVac | LabaIcs,PulmonRehab,FluVac | ·· | 2/Laba+Lama | ALL PROPOSED | ·· |
| 19 | MISSING | MISSING | MISSING | MISSING | MISSING | MISSING | MISSING | MISSING |
| 20 | ALL CDS-SUGGESTED | Sama,PulmonRehab,SmokingTher,FluVac,BetaAgonists | Saba,PulmonRehab,SmokingTher,FluVac,BetaAgonists | SabaSama,PulmonRehab,SmokingTher,FluVac | ·· | 1/Sama | ALL PROPOSED | ovaj pacijent ima astmu treba mu IKS i Laba (This patient has asthma he needs Ics and Laba) |

##### Pulmonologist 2: Evaluation data

Table 13. **Use cases clinical context as collected from the EHR of pulmonologist 2**. Cell data with format m(n) where m,n are numbers denotes contextual use case data entered incorrectly into the EHR (represented outside the brackets) beside the contextual use case data given in the documentation (represented inside the brackets).

| **Use case ID** | **previous CAT score** | **previous mMRC score** | **previous number of exacerbations** | **active COPD treatment** | **current CAT score** | **current mMRC score** | **Current number of Exacerbations** | **Has asthma?** | **previous GOLD group** |
| --- | --- | --- | --- | --- | --- | --- | --- | --- | --- |
| *1* | 6 | 0 | 3 | Saba | 10 | 1 | 2 | FALSE | C |
| *2* | 29 (25) | 3 | 5 (4) | Saba | 29 | 1 (3) | 5 | FALSE | A (D) |
| *3* | 17 | 1 | 1 | Laba | 20 | 1 (2) | 4 | FALSE | B |
| *4* | 9 (8) | 1 | 3 (1) | Saba | 9 | 1 | 3 | TRUE | A |
| *5* | 18 | 2 | 1 | Lama | 26 | 3 | 3 | FALSE | A (B) |
| *6* | 11 (6) | 1 | 0 | Saba | 11 | 1 | 0 | TRUE | A |
| *7* | 9 | 1 | 2 | Lama | 16 | 1 | 4 | FALSE | C |
| *8* | 25 | 3 | 1 | Lama | 27 | 4 | 2 | FALSE | B |
| *9* | 22 | 1 | 0 | Laba+Ics | 20 | 1 | 0 | FALSE | B |
| *10* | ·· | ·· | 0 | ·· | 12 | 1 | 0 | FALSE | ·· |
| *11* | 9 | 1 | 2 | Laba+Lama | 26 | 2 | 1 | FALSE | C |
| *12* | 5 | 1 | 1 | Sama | 8 | 2 | 1 | FALSE | A |
| *13* | ·· | ·· | 0 | ·· | 6 | 1 | 0 (1) | TRUE | ·· |
| *14* | 20 | 1 | 1 | Laba | 21 | 2 | 3 | FALSE | D (B) |
| *15* | 9 | 1 | 0 | Saba | 14 | 2 | 0 | FALSE | A |
| *16* | 8 | 0 | 0 | Laba+Ics | 9 | 1 | 3 | TRUE | A |
| *17* | 26 (27) | 3 (2) | 4 (2) | Laba+Lama | 26 | 3 | 4 | FALSE | A (D) |
| *18* | 6 | 1 | 2 | Laba+Ics | 14 (7) | 2 (1) | 4 | FALSE | C |
| *19* | 37 (32) | 3 (2) | 6 (4) | Saba (Laba+Lama+Ics) | 37 | 3 | 6 | FALSE | A (D) |
| *20* | 7 | 0 | 1 | Sama | 6 | 1 | 2 | TRUE | A |

Table 14. **Results from CDS service ‘copd-assess’ and input for CDS service ‘copd-careplan-review’, as collected from both the EHR of pulmonologist 2 and COPD-CDSS logs**. Keyword MISSING denotes the contextual data was not entered into the EHR.

| **use case ID** | **copd-assess result: GOLD group** | **user-selected GOLD group** | **copd-assess result: personalized COPD treatments** | **had annual influenza vaccine?** | **had pneumococcal vaccine?** | **age** | **is a smoker?** | **Has CKD?** | **Has CVD?** |
| --- | --- | --- | --- | --- | --- | --- | --- | --- | --- |
| 1 | D | D | [ [Laba+Lama], [Laba+Lama+Ics], [Laba+Ics], [Lama]] | FALSE | FALSE | 62 | TRUE | FALSE | TRUE |
| 2 | D | D | [ [Laba+Lama], [Laba+Lama+Ics], [Laba+Ics], [Lama] ] | TRUE | TRUE | 75 | FALSE | FALSE | TRUE |
| 3 | D | D | [ [Laba+Lama], [Laba+Lama+Ics], [Laba+Ics], [Lama]] | FALSE | FALSE | 64 | FALSE | FALSE | FALSE |
| 4 | C | C | [[Lama],[Laba+Ics, Laba+Lama]] | FALSE | FALSE | 50 | TRUE | FALSE | FALSE |
| 5 | D | D | [ [Laba+Lama], [Laba+Lama+Ics], [Laba+Ics], [Lama] ] | TRUE | TRUE | 62 | TRUE | FALSE | TRUE |
| 6 | B | B | [[Laba],[Lama],[Laba+Lama]] | FALSE | FALSE | 57 | TRUE | FALSE | FALSE |
| 7 | D | D | [ [Laba+Lama], [Laba+Lama+Ics], [Laba+Ics], [Lama]] | FALSE | FALSE | 68 | FALSE | TRUE | TRUE (FALSE) |
| 8 | D | D | [ [Laba+Lama], [Laba+Lama+Ics], [Laba+Ics], [Lama]] | TRUE | FALSE | 78 | FALSE (TRUE) | FALSE | TRUE |
| 9 | B | B | [ [Laba+Ics] ,[Laba+Lama], [Laba,Lama] ] | FALSE | FALSE | 69 | TRUE | FALSE | FALSE |
| 10 | B | B | [ [Laba], [Lama], [Laba+Lama] ] | FALSE | FALSE | 65 | FALSE | FALSE | TRUE |
| 11 | B | B | [ [Lama, Laba], [Laba+Lama]] | TRUE | FALSE | 71 | TRUE | TRUE | TRUE |
| 12 | A | A | [[Saba+Sama, Sama],[Saba]] | TRUE | FALSE | 76 | FALSE | FALSE (TRUE) | TRUE (FALSE) |
| 13 | A | A | [[Saba],[Saba+Sama,Sama]] | FALSE | FALSE | 42 | TRUE | FALSE | FALSE |
| 14 | D | D | [ [Laba+Lama], [Laba+Lama+Ics], [Laba+Ics], [Lama] ] | FALSE | FALSE | 59 | TRUE | FALSE | TRUE |
| 15 | B | B | [ [Laba], [Lama], [Laba+Lama] ] | TRUE | FALSE | 61 | FALSE | FALSE | FALSE |
| 16 | C | C | [ [Laba+Ics], [Laba+Lama], [Lama]] | FALSE | FALSE | 46 | FALSE | FALSE | FALSE |
| 17 | D | D | [ [Laba+Lama], [Laba+Lama+Ics], [Laba+Ics], [Lama]] | TRUE | TRUE | 77 | TRUE | FALSE (TRUE) | FALSE |
| 18 | D | D | [ [Laba+Lama], [Laba+Lama+Ics], [Laba+Ics], [Lama] ] | FALSE | FALSE | 62 | FALSE | FALSE | FALSE |
| 19 | D | D | [ [Laba+Lama],[Laba+Lama+Ics],[Laba+Ics],[Lama] ] | TRUE | TRUE | 58 | FALSE | TRUE | TRUE |
| 20 | C | C | [ [Lama], [Laba+Lama, Laba+Ics]] | FALSE | FALSE | 55 | TRUE | FALSE | TRUE |

Table 15. **User-selected treatment(s) for CDS and corresponding results, with pulmonologist comments, for CDS service ‘copd-careplan-review’ and pulmonologist 2.**

| **use case ID** | **user-selected treatments** | **COPD care plan proposal 1** | **COPD care plan proposal 2** | **COPD care plan proposal 3** | **COPD care plan proposal 4** | **User-selected proposal/COPD treatment** | **Context-suitable recommendations** | **Pulmonologist comments** |
| --- | --- | --- | --- | --- | --- | --- | --- | --- |
| 1 | ALL CDS-SUGGESTED | Lama,PulmonRehab,FluVac,Gluccorticds,BetaAgonists | LabaLamaIcs,PulmonRehab,FluVac,BetaAgonists | LabaIcs,PulmonRehab,FluVac,BetaAgonists | LabaLama,PulmonRehab,FluVac,BetaAgonists,Gluccorticds | 1/Lama | PulmonRehab,FluVac,Lama | ·· |
| 2 | ALL CDS-SUGGESTED | LabaLamaIcs,PulmonRehab,BetaAgonists | Lama,PulmonRehab,Gluccorticds,BetaAgonists | LabaLama,PulmonRehab,Gluccorticds,BetaAgonists | LabaIcs,PulmonRehab,BetaAgonists | 1/Laba+Lama+Ics | LabaLamaIcs, PulmonRehab | ·· |
| 3 | ALL CDS-SUGGESTED | Lama,PulmonRehab,FluVac,Gluccorticds | LabaLama,PulmonRehab,FluVac,Gluccorticds | LabaLamaIcs,PulmonRehab,FluVac | LabaIcs,PulmonRehab,FluVac | 1/Lama | Lama,PulmonRehab,FluVac | ·· |
| 4 | ALL CDS-SUGGESTED | LabaLama,PulmonRehab,FluVac,SmokingTher,Gluccorticds | Lama,PulmonRehab,FluVac,SmokingTher,Gluccorticds | LabaIcs,PulmonRehab,FluVac,SmokingTher | ·· | 3/Laba+Ics | ALL CDS-SUGGESTED | ·· |
| 5 | ALL CDS-SUGGESTED | LabaLamaIcs,PulmonRehab,SmokingTher,BetaAgonists | LabaIcs,BetaAgonists,PulmonRehab,SmokingTher | LabaLama,PulmonRehab,SmokingTher,Gluccorticds,BetaAgonists | Lama,PulmonRehab,SmokingTher,Gluccorticds,BetaAgonists | 1/Laba+Lama+Ics | PulmonRehab,SmokingTher,LabaLamaIcs | ·· |
| 6 | ALL CDS-SUGGESTED | Lama,PulmonRehab,SmokingTher,Gluccorticds, FluVac | Laba,PulmonRehab,SmokingTher,Gluccorticds, FluVac | LabaLama,PulmonRehab,SmokingTher,Gluccorticds, FluVac | ·· | 1/Lama | PulmonRehab,SmokingTher,Lama, FluVac | ·· |
| 7 | ALL CDS-SUGGESTED | LabaIcs,PulmonRehab,LabaLamaCkd, FluVac,BetaAgonists,PneumnVac | LabaLamaIcs,PulmonRehab,LabaLamaCkd, FluVac,BetaAgonists,PneumnVac | LabaLama,PulmonRehab,LabaLamaCkd, FluVac,BetaAgonists,PneumnVac,Gluccorticds | Lama,PulmonRehab, FluVac,PneumnVac,BetaAgonists,Gluccorticds | 1/Laba+Ics | PulmonRehab,LabaLamaCkd,LabaIcs, FluVac,PneumnVac | ·· |
| 8 | ALL CDS-SUGGESTED | Lama,PulmonRehab,BetaAgonists,PneumnVac,Gluccorticds | LabaIcs,PulmonRehab,BetaAgonists,PneumnVac | LabaLamaIcs,PulmonRehab,BetaAgonists,PneumnVac | LabaLama,PulmonRehab,BetaAgonists,PneumnVac,Gluccorticds | 1/Lama | PulmonRehab,Lama,PneumnVac | ·· |
| 9 | ALL CDS-SUGGESTED | Lama,PulmonRehab,SmokingTher,FluVac,Gluccorticds,PneumnVac | Laba,PulmonRehab,SmokingTher,FluVac,Gluccorticds,PneumnVac | LabaLama,PulmonRehab,SmokingTher,FluVac,Gluccorticds,PneumnVac | LabaIcs,PulmonRehab,SmokingTher,FluVac,PneumnVac | 1/Lama | ALL CDS-SUGGESTED | ·· |
| 10 | ALL CDS-SUGGESTED | Laba,PulmonRehab,Gluccorticds, FluVac, PneumnVac | Lama,PulmonRehab,Gluccorticds, FluVac, PneumnVac, BetaAgonists | LabaLama,PulmonRehab,Gluccorticds,BetaAgonists, FluVac, PneumnVac | ·· | 1/Laba | ALL CDS-SUGGESTED | ·· |
| 11 | ALL CDS-SUGGESTED | Laba,PulmonRehab,SmokingTher,PneumnVac,Gluccorticds | Lama,PulmonRehab,SmokingTher,PneumnVac,Gluccorticds,,BetaAgonists | LabaLama,PulmonRehab,SmokingTher,PneumnVac,Gluccorticds,LabaLamaCkd,BetaAgonists | ·· | 1/Laba+Lama | ALL CDS-SUGGESTED | ·· |
| 12 | ALL CDS-SUGGESTED | SabaSama,PulmonRehab,PneumnVac,BetaAgonists | Sama,PulmonRehab,PneumnVac,BetaAgonists | Saba,PulmonRehab,PneumnVac | ·· | 1/Saba+Sama | PulmonRehab, PneumnVac,SabaSama | ·· |
| 13 | ALL CDS-SUGGESTED | Saba,PulmonRehab,SmokingTher,FluVac | Sama,PulmonRehab,SmokingTher,FluVac | SabaSama,PulmonRehab,SmokingTher,FluVac | ·· | 1/Saba | ALL CDS-SUGGESTED | ·· |
| 14 | ALL CDS-SUGGESTED | Lama,PulmonRehab,SmokingTher,FluVac,Gluccorticds,BetaAgonists | LabaLama,PulmonRehab,SmokingTher,FluVac,Gluccorticds,BetaAgonists | LabaLamaIcs,PulmonRehab,SmokingTher,FluVac,BetaAgonists | LabaIcs,PulmonRehab,SmokingTher,FluVac,BetaAgonists | 1/Lama | Lama,PulmonRehab,SmokingTher,FluVac,Gluccorticds | ·· |
| 15 | ALL CDS-SUGGESTED | Laba,PulmonRehab,Gluccorticds | Lama,PulmonRehab,Gluccorticds | LabaLama,PulmonRehab,Gluccorticds | ·· | 1/Laba | ALL CDS-SUGGESTED | ·· |
| 16 | ALL CDS-SUGGESTED | LabaIcs,PulmonRehab,FluVac | Lama,PulmonRehab,Gluccorticds,FluVac | LabaLama,PulmonRehab,FluVac,Gluccorticds | ·· | 1/Laba+Ics | ALL CDS-SUGGESTED | ·· |
| 17 | ALL CDS-SUGGESTED | Lama,PulmonRehab,Gluccorticds,SmokingTher | LabaLama,PulmonRehab,Gluccorticds,SmokingTher | LabaLamaIcs,PulmonRehab,SmokingTher | LabaIcs,PulmonRehab,SmokingTher | 1/Lama | ALL CDS-SUGGESTED | ·· |
| 18 | ALL CDS-SUGGESTED | Lama,PulmonRehab,Gluccorticds,FluVac | LabaLama,PulmonRehab,Gluccorticds,FluVac | LabaLamaIcs,PulmonRehab,FluVac | LabaIcs,PulmonRehab,FluVac | 1/Lama | ALL CDS-SUGGESTED | ·· |
| 19 | ALL CDS-SUGGESTED | LabaIcs,BetaAgonists,LabaLamaCkd,PulmonRehab | LabaLamaIcs,BetaAgonists,LabaLamaCkd,PulmonRehab | Lama,BetaAgonists,LabaLamaCkd,PulmonRehab,Gluccorticds | LabaLama,BetaAgonists,LabaLamaCkd,PulmonRehab,Gluccorticds | 1/Laba+Ics | ALL CDS-SUGGESTED | ·· |
| 20 | ALL CDS-SUGGESTED | LabaIcs,PulmonRehab,SmokingTher,FluVac,BetaAgonists | Lama,PulmonRehab,SmokingTher,FluVac,BetaAgonists,Gluccorticds | LabaLama,PulmonRehab,SmokingTher,FluVac,BetaAgonists | ·· | 1/Laba+Ics | ALL CDS-SUGGESTED | ·· |

##### Pulmonologist 3: Evaluation data

Table 16. **Use cases clinical context as collected from the EHR of pulmonologist 3**. Cell data with format m(n) where m,n are numbers denotes contextual use case data entered incorrectly into the EHR (represented outside the brackets) beside the contextual use case data given in the documentation (represented inside the brackets).

| **Use case ID** | **previous CAT score** | **previous mMRC score** | **previous number of exacerbations** | **active COPD treatment** | **current CAT score** | **current mMRC score** | **Current number of Exacerbations** | **Has asthma?** | **previous GOLD group** |
| --- | --- | --- | --- | --- | --- | --- | --- | --- | --- |
| *1* | 6 | 0 | 3 | Saba | 10 | 1 | 2 | FALSE | C |
| *2* | 29 (25) | 3 | 5 (4) | Saba (Laba+Lama+Ics) | 29 | 3 | 5 | FALSE | A (D) |
| *3* | 17 | 1 | 1 | Laba | 20 | 2 | 4 | FALSE | B |
| *4* | 8 | 1 | 1 | Saba | 9 | 1 | 3 | TRUE | A |
| *5* | 18 | 2 | 1 | Lama | 26 | 3 | 3 | FALSE | B |
| *6* | 11 (6) | 1 | 0 | Saba | 11 | 1 | 0 | TRUE | A |
| *7* | 9 | 1 | 2 | Lama | 16 | 1 | 4 | FALSE | C |
| *8* | 25 | 3 | 1 | Lama | 27 | 4 | 2 | FALSE | B |
| *9* | 22 | 1 | 0 | Laba+Ics | 20 | 1 | 0 | FALSE | B |
| *10* | ·· | ·· | 0 | ·· | 12 | 1 | 0 | FALSE | ·· |
| *11* | 9 | 1 | 2 | Laba+Lama | 26 | 2 | 1 | FALSE | C |
| *12* | 5 | 1 | 1 | Sama | 8 | 2 | 1 | FALSE | A |
| *13* | ·· | ·· | 0 | ·· | 6 | 1 | 0 (1) | TRUE | ·· |
| *14* | 20 | 1 | 1 | Laba | 21 | 2 | 3 | FALSE | D (B) |
| *15* | 9 | 1 | 0 | Saba | 14 | 2 | 0 | FALSE | A |
| *16* | 8 | 0 | 0 | Laba+Ics | 9 | 1 | 3 | TRUE | A |
| *17* | 26 (27) | 3 (2) | 4 (2) | Laba+Lama | 26 | 3 | 4 | FALSE | A (D) |
| *18* | 6 | 1 | 2 | Laba+Ics | 14 (7) | 2 (1) | 4 | FALSE | C |
| *19* | 37 (32) | 3 (2) | 6 (4) | Saba (Laba+Lama+Ics) | 37 | 3 | 6 | FALSE | A (D) |
| *20* | 7 | 0 | 1 | Sama | 6 | 1 | 2 | TRUE | A |

Table 10. **Results from CDS service ‘copd-assess’ and input for CDS service ‘copd-careplan-review’, as collected from both the EHR of pulmonologist 3 and COPD-CDSS logs**. Keyword MISSING denotes the contextual data was not entered into the EHR.

| **use case ID** | **copd-assess result: GOLD group** | **user-selected GOLD group** | **copd-assess result: personalized COPD treatments** | **had annual influenza vaccine?** | **had pneumococcal vaccine?** | **age** | **is a smoker?** | **Has CKD?** | **Has CVD?** |
| --- | --- | --- | --- | --- | --- | --- | --- | --- | --- |
| 1 | D | D | [ [Laba+Lama], [Laba+Lama+Ics], [Laba+Ics], [Lama]] | FALSE | FALSE | 62 | TRUE | FALSE | TRUE |
| 2 | D | D | [ [Laba+Lama],[Laba+Lama+Ics],[Laba+Ics],[Lama] ] | FALSE | FALSE | 75 | FALSE | FALSE (TRUE) | TRUE |
| 3 | D | D | [ [Laba+Lama], [Laba+Lama+Ics], [Laba+Ics], [Lama]] | FALSE | FALSE | 64 | FALSE | FALSE | FALSE |
| 4 | C | C | [[Lama],[Laba+Ics, Laba+Lama]] | FALSE | FALSE | 50 | TRUE | FALSE | FALSE |
| 5 | D | D | [ [Laba+Lama], [Laba+Lama+Ics], [Laba+Ics], [Lama] ] | TRUE | TRUE | 62 | TRUE | FALSE | TRUE |
| 6 | B | B | [[Laba],[Lama],[Laba+Lama]] | FALSE | FALSE | 57 | TRUE | FALSE | FALSE |
| 7 | D | D | [ [Laba+Lama], [Laba+Lama+Ics], [Laba+Ics], [Lama]] | FALSE | FALSE | 68 | FALSE | TRUE | TRUE (FALSE) |
| 8 | D | D | [ [Laba+Lama], [Laba+Lama+Ics], [Laba+Ics], [Lama]] | TRUE | FALSE | 78 | FALSE | FALSE | FALSE (TRUE) |
| 9 | B | B | [ [Laba+Ics], ,[Laba+Lama], [Laba,Lama] ] | FALSE | FALSE | 69 | TRUE | FALSE | FALSE |
| 10 | B | B | [ [Laba], [Lama], [Laba+Lama] ] | FALSE | FALSE | 65 | FALSE | FALSE | TRUE |
| 11 | B | B | [ [Lama, Laba], [Laba+Lama]] | TRUE | FALSE | 71 | TRUE | TRUE (FALSE) | FALSE (TRUE) |
| 12 | A | D | [ [Laba+Lama], [Laba+Lama+Ics], [Laba+Ics], [Lama] ] | TRUE | FALSE | 76 | FALSE | FALSE (TRUE) | TRUE (FALSE) |
| 13 | A | A | [[Saba],[Saba+Sama,Sama]] | FALSE | FALSE | 42 | TRUE | FALSE | FALSE |
| 14 | D | D | [ [Laba+Lama], [Laba+Lama+Ics], [Laba+Ics], [Lama] ] | FALSE | FALSE | 59 | TRUE | FALSE | TRUE |
| 15 | B | B | [ [Laba], [Lama], [Laba+Lama] ] | TRUE | FALSE | 61 | FALSE | FALSE | FALSE |
| 16 | C | C | [ [Laba+Ics], [Laba+Lama], [Lama]] |  | FALSE | 46 | FALSE | FALSE | FALSE |
| 17 | D | D | [ [Laba+Lama+Ics], [Laba+Ics], [Laba+Lama], [Lama]] | TRUE | TRUE | 77 | TRUE | TRUE | FALSE |
| 18 | D | D | [ [Laba+Lama], [Laba+Lama+Ics], [Laba+Ics], [Lama] ] | FALSE | FALSE | 62 | FALSE | FALSE | FALSE |
| 19 | D | D | [ [Laba+Lama],[Laba+Lama+Ics],[Laba+Ics],[Lama] ] | TRUE | TRUE | 58 | FALSE | TRUE | TRUE |
| 20 | C | C | [ [Lama], [Laba+Lama, Laba+Ics]] | FALSE | FALSE | 55 | TRUE | FALSE | TRUE |

Table 17. **User-selected treatment(s) for CDS and corresponding results, with pulmonologist comments, for CDS service ‘copd-careplan-review’ and pulmonologist 3.**

| **use case ID** | **user-selected treatments** | **COPD care plan proposal 1** | **COPD care plan proposal 2** | **COPD care plan proposal 3** | **COPD care plan proposal 4** | **User-selected proposal/COPD treatment** | **Context-suitable recommendations** | **Pulmonologist comments** |
| --- | --- | --- | --- | --- | --- | --- | --- | --- |
| 1 | ALL CDS-SUGGESTED | LabaLamaIcs,PulmonRehab,FluVac,BetaAgonists,SmokingTher | LabaLama,PulmonRehab,FluVac,BetaAgonists,Gluccorticds,SmokingTher | LabaIcs,PulmonRehab,FluVac,BetaAgonists,SmokingTher | Lama,PulmonRehab,FluVac,BetaAgonists,Gluccorticds,SmokingTher | 1/Laba+Lama+Ics | PulmonRehab,FluVac,LabaLamaIcs,SmokingTher | ·· |
| 2 | ALL CDS-SUGGESTED | LabaIcs,PulmonRehab,,FluVac,BetaAgonists, PneumVac | Lama,PulmonRehab,PneumVac,FluVac,BetaAgonists,Gluccorticds | LabaLamaIcs,PulmonRehab,FluVac,BetaAgonists,PneumVac | LabaLama,PulmonRehab,FluVac,BetaAgonists,PneumVac,Gluccorticds | 1/Laba+Ics | LabaIcs,PulmonRehab,FluVac, PneumVac | ·· |
| 3 | ALL CDS-SUGGESTED | LabaLamaIcs,PulmonRehab,FluVac | LabaLama,PulmonRehab,FluVac,Gluccorticds | LabaIcs,PulmonRehab,FluVac | Lama,PulmonRehab,FluVac,Gluccorticds | 1/Laba+Lama+Ics | ALL PROPOSED | ·· |
| 4 | ALL CDS-SUGGESTED | LabaIcs,PulmonRehab,FluVac,SmokingTher | LabaLama,PulmonRehab,FluVac,SmokingTher,Gluccorticds | Lama,PulmonRehab,FluVac,SmokingTher,Gluccorticds | ·· | 1/Laba+Ics | ALL PROPOSED | ·· |
| 5 | ALL CDS-SUGGESTED | Lama,PulmonRehab,SmokingTher,Gluccorticds,BetaAgonists | LabaLama,PulmonRehab,SmokingTher,Gluccorticds,BetaAgonists | LabaIcs,BetaAgonists,PulmonRehab,SmokingTher | LabaLamaIcs,PulmonRehab,SmokingTher,BetaAgonists | 2/Laba+Lama | PulmonRehab,SmokingTher,Gluccorticds,LabaLama | ·· |
| 6 | ALL CDS-SUGGESTED | Laba,PulmonRehab,SmokingTher,Gluccorticds, FluVac | LabaLama,PulmonRehab,SmokingTher,Gluccorticds, FluVac | Lama,PulmonRehab,SmokingTher,Gluccorticds, FluVac | ·· | 1/Laba | PulmonRehab,SmokingTher,Laba, FluVac | ·· |
| 7 | ALL CDS-SUGGESTED | LabaIcs,PulmonRehab, FluVac,PneumnVac,BetaAgonists | Lama,PulmonRehab, FluVac,BetaAgonists,PneumnVac,Gluccorticds | LabaLamaIcs,PulmonRehab, FluVac,BetaAgonists,PneumnVac | LabaLama,PulmonRehab, FluVac,BetaAgonists,PneumnVac,Gluccorticds | 1/Laba+Ics | PulmonRehab,LabaIcs, FluVac,PneumnVac | ·· |
| 8 | ALL CDS-SUGGESTED | LabaLamaIcs,PulmonRehab,,PneumnVac | Lama,PulmonRehab,PneumnVac,Gluccorticds | LabaIcs,PulmonRehab,PneumnVac | LabaLama,PulmonRehab,PneumnVac,Gluccorticds | 1/Laba+Lama+Ics | ALL PROPOSED | ·· |
| 9 | ALL CDS-SUGGESTED | Lama,PulmonRehab,SmokingTher,FluVac,Gluccorticds,PneumnVac | LabaLama,PulmonRehab,SmokingTher,FluVac,Gluccorticds,PneumnVac | Laba,PulmonRehab,SmokingTher,FluVac,Gluccorticds,PneumnVac | LabaIcs,PulmonRehab,SmokingTher,FluVac,PneumnVac | 1/Lama | PulmonRehab,SmokingTher,FluVac,Lama,PneumnVac | ·· |
| 10 | ALL CDS-SUGGESTED | LabaLama,PulmonRehab,Gluccorticds, FluVac, PneumnVac,BetaAgonists | Lama,PulmonRehab,Gluccorticds, FluVac, PneumnVac, BetaAgonists | Laba,PulmonRehab,Gluccorticds, FluVac, PneumnVac | ·· | 1/Laba+Lama | PulmonRehab,LabaLama, FluVac, PneumnVac | ·· |
| 11 | ALL CDS-SUGGESTED | Laba,PulmonRehab,SmokingTher,PneumnVac,Gluccorticds | Lama,PulmonRehab,SmokingTher,PneumnVac,Gluccorticds,,BetaAgonists | LabaLama,PulmonRehab,SmokingTher,PneumnVac,Gluccorticds,BetaAgonists | ·· | 1/Laba+Lama | SmokingTher,PneumnVac,Laba | ·· |
| 12 | ALL CDS-SUGGESTED | LabaLamaIcs,PulmonRehab,PneumnVac,BetaAgonists | Lama,PulmonRehab,PneumnVac,BetaAgonists,Gluccorticds | LabaLama,PulmonRehab,PneumnVac,BetaAgonists,Gluccorticds | LabaIcs,PulmonRehab,PneumnVac,BetaAgonists | 1/Laba+Lama | ALL PROPOSED | ·· |
| 13 | ALL CDS-SUGGESTED | Sama,PulmonRehab,SmokingTher,FluVac | SabaSama,PulmonRehab,SmokingTher,FluVac | Saba,PulmonRehab,SmokingTher,FluVac | ·· | 1/Sama | ALL PROPOSED | ·· |
| 14 | ALL CDS-SUGGESTED | LabaLamaIcs,PulmonRehab,SmokingTher,FluVac,BetaAgonists | LabaLama,PulmonRehab,SmokingTher,FluVac,Gluccorticds,BetaAgonists | LabaIcs,PulmonRehab,SmokingTher,FluVac,BetaAgonists | Lama,PulmonRehab,SmokingTher,FluVac,BetaAgonists,Gluccorticds | 1/Laba+Lama+Ics | LabaLamaIcs,PulmonRehab,SmokingTher,FluVac | ·· |
| 15 | ALL CDS-SUGGESTED | Laba,PulmonRehab,Gluccorticds | LabaLama,PulmonRehab,Gluccorticds | Lama,PulmonRehab,Gluccorticds | ·· | 1/Laba | ALL PROPOSED | ·· |
| 16 | ALL CDS-SUGGESTED | Lama,PulmonRehab,Gluccorticds,FluVac | LabaLama,PulmonRehab,FluVac,Gluccorticds | LabaIcs,PulmonRehab,FluVac | ·· | 1/Lama | Lama,PulmonRehab,FluVac | ·· |
| 17 | ALL CDS-SUGGESTED | Lama,PulmonRehab,Gluccorticds,SmokingTher | LabaLama,PulmonRehab,Gluccorticds,SmokingTher, LabaLamaCkd | LabaLamaIcs,PulmonRehab,SmokingTher, LabaLamaCkd | LabaIcs,PulmonRehab,SmokingTher, LabaLamaCkd | 1/Lama | ALL PROPOSED | ·· |
| 18 | ALL CDS-SUGGESTED | LabaLamaIcs,PulmonRehab,FluVac | Lama,PulmonRehab,Gluccorticds,FluVac | LabaLama,PulmonRehab,Gluccorticds,FluVac | LabaIcs,PulmonRehab,FluVac | 1/Laba+Lama+Ics | ALL PROPOSED | ·· |
| 19 | ALL CDS-SUGGESTED | LabaIcs,BetaAgonists,LabaLamaCkd,PulmonRehab | Lama,BetaAgonists,LabaLamaCkd,PulmonRehab,Gluccorticds | LabaLama,BetaAgonists,LabaLamaCkd,PulmonRehab,Gluccorticds | LabaLamaIcs,BetaAgonists,LabaLamaCkd,PulmonRehab | 1/Laba+Ics | ALL PROPOSED | ·· |
| 20 | ALL CDS-SUGGESTED | Lama,PulmonRehab,SmokingTher,FluVac,BetaAgonists,Gluccorticds | LabaLama,PulmonRehab,SmokingTher,FluVac,BetaAgonists,Gluccorticds | LabaIcs,PulmonRehab,SmokingTher,FluVac,BetaAgonists | ·· | 1/Lama | ALL PROPOSED | ·· |

##### Pulmonologist 4: Evaluation data

Table 18. **Use cases clinical context as collected from the EHR of pulmonologist 4**. Cell data with format m(n) where m,n are numbers denotes contextual use case data entered incorrectly into the EHR (represented outside the brackets) beside the contextual use case data given in the documentation (represented inside the brackets).

| **Use case ID** | **previous CAT score** | **previous mMRC score** | **previous number of exacerbations** | **active COPD treatment** | **current CAT score** | **current mMRC score** | **Current number of Exacerbations** | **Has asthma?** | **previous GOLD group** |
| --- | --- | --- | --- | --- | --- | --- | --- | --- | --- |
| *1* | 10 (6) | 1 (0) | 0 (3) | 10 | 1 | 0 (2) | FALSE | FALSE (TRUE) | A (C) |
| *2* | 29 (25) | 3 | 0 (4) | 29 | 3 | 5 | FALSE | FALSE (TRUE) | A |
| *3* | 17 | 1 | 1 | 20 | 2 | 0 (4) | FALSE | FALSE | D (B) |
| *4* | 9 (8) | 1 | 3 (1) | 9 | 1 | 3 | TRUE | TRUE | A |
| *5* | 20 (18) | 2 | 3 (1) | 26 | 3 | 0 (3) | FALSE | TRUE | D (B) |
| *6* | 11 (6) | 1 | 0 | 11 | 1 | 0 | TRUE | TRUE | A |
| *7* | 9 | 1 | 2 | 16 | 1 | 4 | FALSE | FALSE | C |
| *8* | 25 | 3 | 1 | 27 | 4 | 2 | FALSE | FALSE | B |
| *9* | 22 | 1 | 0 | 20 | 1 | 0 | FALSE | TRUE | B |
| *10* | ·· | ·· | 0 | 12 | 1 | 0 | FALSE | FALSE | ·· |
| *11* | 9 | 1 | 2 | 26 | 2 | 1 | FALSE | TRUE | C |
| *12* | 5 | 1 | 1 | 8 | 2 | 1 | FALSE | FALSE | A |
| *13* | ·· | ·· | 0 | 6 | 1 | 0 (1) | TRUE | FALSE (TRUE) | ·· |
| *14* | 20 | 1 | 1 | 21 | 2 | 3 | FALSE | TRUE | B |
| *15* | 9 | 1 | 0 | 14 | 2 | 0 | FALSE | FALSE | A |
| *16* | 8 | 0 | 0 | 9 | 1 | 3 | TRUE | FALSE | A |
| *17* | 26 (27) | 3 (2) | 4 (2) | 26 | 3 | 4 | FALSE | TRUE | A (D) |
| *18* | 6 | 1 | 2 | 14 (7) | 2 (1) | 4 | FALSE | FALSE | C |
| *19* | 37 (32) | 3 (2) | 6 (4) | 37 | 3 | 6 | FALSE | FALSE | A (D) |
| *20* | 7 | 0 | 1 | 6 | 1 | 2 | TRUE | TRUE | A |

Table 19. **Results from CDS service ‘copd-assess’ and input for CDS service ‘copd-careplan-review’, as collected from both the EHR of pulmonologist 4 and COPD-CDSS logs**. Keyword MISSING denotes the contextual data was not entered into the EHR.

| **use case ID** | **copd-assess result: GOLD group** | **user-selected GOLD group** | **copd-assess result: personalized COPD treatments** | **had annual influenza vaccine?** | **had pneumococcal vaccine?** | **age** | **is a smoker?** | **Has CKD?** | **Has CVD?** |
| --- | --- | --- | --- | --- | --- | --- | --- | --- | --- |
| 1 | B (D) | B (D) | [ [Laba], [Lama], [Laba+Lama]] | FALSE | FALSE | 62 | FALSE (TRUE) | FALSE | TRUE |
| 2 | B (D) | B | [ [Laba], [Lama], [Laba+Lama] ] | TRUE | TRUE | 75 | FALSE | TRUE | TRUE |
| 3 | B (D) | B | [ [Laba], [Laba+Lama, Lama] ] | FALSE | FALSE | 64 | FALSE | FALSE | FALSE |
| 4 | C | C | [[Lama],[Laba+Ics, Laba+Lama]] | FALSE | FALSE | 50 | TRUE | FALSE | FALSE |
| 5 | B (D) | B (D) | [ [Laba, Laba+Lama], [Lama] ] | TRUE | TRUE | 62 | TRUE | FALSE | TRUE |
| 6 | B | B | [[Laba],[Lama],[Laba+Lama]] | FALSE | FALSE | 57 | TRUE | FALSE | FALSE |
| 7 | D | D | [ [Laba+Lama], [Laba+Lama+Ics], [Laba+Ics], [Lama]] | FALSE | FALSE | 68 | FALSE | TRUE | TRUE (FALSE) |
| 8 | D | D | [ [Laba+Lama], [Laba+Lama+Ics], [Laba+Ics], [Lama]] | TRUE | FALSE | 78 | FALSE | FALSE | TRUE |
| 9 | B | B | [ [Laba+Ics],[Laba+Lama], [Laba,Lama] ] | FALSE | FALSE | 69 | TRUE | FALSE | FALSE |
| 10 | B | B | [ [Laba], [Lama], [Laba+Lama] ] | FALSE | FALSE | 65 | FALSE | FALSE | TRUE |
| 11 | A (B) | A | [ [Lama, Laba], [Laba+Lama]] | TRUE | FALSE | 71 | TRUE | TRUE | TRUE (FALSE) |
| 12 | A (D) | D | [ [Laba+Lama], [Laba+Lama+Ics], [Laba+Ics], [Lama] ] | TRUE | FALSE | 76 | FALSE | TRUE | TRUE |
| 13 | A | A | [[Saba],[Saba+Sama,Sama]] | FALSE | FALSE | 42 | FALSE (TRUE) | FALSE | FALSE |
| 14 | D | D | [ [Laba+Lama], [Laba+Lama+Ics], [Laba+Ics], [Lama] ] | FALSE | FALSE | 59 | TRUE | FALSE | TRUE |
| 15 | B | B | [ [Laba], [Lama], [Laba+Lama] ] | TRUE | FALSE | 61 | FALSE | FALSE | FALSE |
| 16 | C | C | [ [Laba+Ics], [Laba+Lama], [Lama]] | FALSE | FALSE | 46 | FALSE | FALSE | FALSE |
| 17 | D | D | [ [Laba+Lama], [Laba+Lama+Ics], [Laba+Ics], [Lama] ] | TRUE | TRUE | 77 | TRUE | TRUE | FALSE |
| 18 | D (C) | D | [ [Laba+Lama], [Laba+Lama+Ics], [Laba+Ics], [Lama] ] | FALSE | FALSE | 62 | FALSE | FALSE | FALSE |
| 19 | D | D | [ [Laba+Lama],[Laba+Lama+Ics],[Laba+Ics],[Lama] ] | TRUE | TRUE |  | FALSE | TRUE | TRUE |
| 20 | C | C | [ [Lama], [Laba+Lama, Laba+Ics]] | FALSE | FALSE | 55 | TRUE | FALSE | TRUE |

Table 20. **User-selected treatment(s) for CDS and corresponding results, with pulmonologist comments, for CDS service ‘copd-careplan-review’ and pulmonologist 4.**

| **use case ID** | **user-selected treatments** | **COPD care plan proposal 1** | **COPD care plan proposal 2** | **COPD care plan proposal 3** | **COPD care plan proposal 4** | **User-selected proposal/COPD treatment** | **Context-suitable recommendations** | **Pulmonologist comments** |
| --- | --- | --- | --- | --- | --- | --- | --- | --- |
| 1 | ALL CDS-SUGGESTED | Laba,PulmonRehab,FluVac,Gluccorticds | Laba,PulmonRehab,FluVac,Gluccorticds,BetaAgonists | LabaLama,PulmonRehab,FluVac,Gluccorticds,BetaAgonists | ·· | 1/Laba | ALL PROPOSED | ·· |
| 2 | ALL CDS-SUGGESTED | Laba,PulmonRehab,Gluccorticds | LabaLama,PulmonRehab,Gluccorticds,BetaAgonists | Lama,PulmonRehab,Gluccorticds,BetaAgonists, | ·· | 1/Laba | ALL PROPOSED | ·· |
| 3 | ALL CDS-SUGGESTED | Laba,PulmonRehab,FluVac,Gluccorticds | LabaLama,PulmonRehab,FluVac,Gluccorticds | Lama,PulmonRehab,FluVac,Gluccorticds | ·· | 1/Laba | ALL PROPOSED | ·· |
| 4 | ALL CDS-SUGGESTED | Lama,PulmonRehab,FluVac,SmokingTher,Gluccorticds | LabaLama,PulmonRehab,FluVac,SmokingTher,Gluccorticds | LabaIcs,PulmonRehab,FluVac,SmokingTher | ·· | 1/Lama | Lama,PulmonRehab,FluVac,SmokingTher | ·· |
| 5 | ALL CDS-SUGGESTED | LabaLama,PulmonRehab,SmokingTher,BetaAgonists,Gluccorticds | Lama,PulmonRehab,SmokingTher,BetaAgonists,Gluccorticds | Laba,PulmonRehab,SmokingTher,Gluccorticds | ·· | 1/Laba+Lama | ALL PROPOSED | ·· |
| 6 | ALL CDS-SUGGESTED | Laba,PulmonRehab,SmokingTher,Gluccorticds, FluVac | Lama,PulmonRehab,SmokingTher,Gluccorticds, FluVac | LabaLama,PulmonRehab,SmokingTher,Gluccorticds, FluVac | ·· | 1/Laba | PulmonRehab,SmokingTher,Lama, FluVac | ·· |
| 7 | ALL CDS-SUGGESTED | LabaLamaIcs,PulmonRehab,LabaLamaCkd, FluVac,BetaAgonists,PneumnVac | LabaIcs,PulmonRehab,LabaLamaCkd, FluVac,BetaAgonists,PneumnVac | Lama,PulmonRehab, FluVac,PneumnVac,BetaAgonists,Gluccorticds | LabaLama,PulmonRehab,LabaLamaCk, FluVac,BetaAgonists,PneumnVac,Gluccorticds | 1/Laba+Lama+Ics | PulmonRehab,LabaLamaIcs, FluVac,PneumnVac,BetaAgonists | ·· |
| 8 | ALL CDS-SUGGESTED | LabaIcs,PulmonRehab,BetaAgonists,PneumnVac | LabaLamaIcs,PulmonRehab,BetaAgonists,PneumnVac | LabaLama,PulmonRehab,BetaAgonists,PneumnVac,Gluccorticds | Lama,PulmonRehab,BetaAgonists,PneumnVac,Gluccorticds | 1/Laba+Ics | PulmonRehab,BetaAgonists,PneumnVac | Pulmonologist did not select a COPD treatment in column 'Context suitable recs' |
| 9 | ALL CDS-SUGGESTED | LabaLama,PulmonRehab,SmokingTher,FluVac,Gluccorticds,PneumnVac | Laba,PulmonRehab,SmokingTher,FluVac,Gluccorticds,PneumnVac | Lama,PulmonRehab,SmokingTher,FluVac,Gluccorticds,PneumnVac | LabaIcs,PulmonRehab,SmokingTher,FluVac,PneumnVac | 1/Laba+Lama | ALL PROPOSED | ·· |
| 10 | ALL CDS-SUGGESTED | LabaLama,PulmonRehab,Gluccorticds, FluVac, PneumnVac, BetaAgonists | Lama,PulmonRehab,Gluccorticds, FluVac, PneumnVac, BetaAgonists | Laba,PulmonRehab,Gluccorticds, FluVac, PneumnVac | ·· | 1/Laba+Lama | ALL PROPOSED | ·· |
| 11 | ALL CDS-SUGGESTED | LabaLama,PulmonRehab,SmokingTher,PneumnVac,LabaLamaCkd,Gluccorticds,BetaAgonists | LabaLama,PulmonRehab,SmokingTher,PneumnVac,Gluccorticds | LabaLama,PulmonRehab,SmokingTher,PneumnVac,Gluccorticds,BetaAgonists | ·· | 1/Laba+Lama | PulmonRehab,SmokingTher,PneumnVac,Gluccorticds,LabaLama,BetaAgonists | ·· |
| 12 | ALL CDS-SUGGESTED | LabaLama,PulmonRehab,PneumnVac,LabaLamaCkd,BetaAgonists,Gluccorticds | LabaIcs,PulmonRehab,PneumnVac,LabaLamaCkd,BetaAgonists | LabaLamaIcs,PulmonRehab,PneumnVac,LabaLamaCkd,BetaAgonists | Lama,PulmonRehab,PneumnVac,BetaAgonists,Gluccorticds | 1/Laba+Lama | PulmonRehab,PneumnVac,LabaLama,BetaAgonists,Gluccorticds | ·· |
| 13 | ALL CDS-SUGGESTED | Saba,PulmonRehab,FluVac | Sama,PulmonRehab,FluVac | SabaSama,PulmonRehab,FluVac | ·· | 1/Saba | ALL PROPOSED | ·· |
| 14 | ALL CDS-SUGGESTED | LabaIcs,PulmonRehab,SmokingTher,FluVac,BetaAgonists | LabaLama,PulmonRehab,SmokingTher,FluVac,BetaAgonists,Gluccorticds | Lama,PulmonRehab,SmokingTher,FluVac,BetaAgonists,Gluccorticds | LabaLamaIcs,PulmonRehab,SmokingTher,FluVac,BetaAgonists | 1/Laba+Ics | ALL PROPOSED | ·· |
| 15 | ALL CDS-SUGGESTED | Laba,PulmonRehab,Gluccorticds | Lama,PulmonRehab,Gluccorticds | LabaLama,PulmonRehab,Gluccorticds | ·· | 1/Laba | ALL PROPOSED | ·· |
| 16 | ALL CDS-SUGGESTED | LabaIcs,PulmonRehab,FluVac | LabaLama,PulmonRehab,FluVac,Gluccorticds | Lama,PulmonRehab,Gluccorticds,FluVac | ·· | 1/Laba+Ics | ALL PROPOSED | ·· |
| 17 | ALL CDS-SUGGESTED | LabaLama,PulmonRehab,Gluccorticds,SmokingTher, LabaLamaCkd | LabaLamaIcs,PulmonRehab,SmokingTher, LabaLamaCkd | LabaIcs,PulmonRehab,SmokingTher, LabaLamaCkd | Lama,PulmonRehab,Gluccorticds,SmokingTher | 1/Laba+Lama | LabaLama,PulmonRehab,Gluccorticds,SmokingTher | ·· |
| 18 | ALL CDS-SUGGESTED | Lama,PulmonRehab,Gluccorticds,FluVac | LabaLama,PulmonRehab,Gluccorticds,FluVac | LabaLamaIcs,PulmonRehab,FluVac | LabaIcs,PulmonRehab,FluVac | 1/Lama | ALL PROPOSED | ·· |
| 19 | ALL CDS-SUGGESTED | Lama,BetaAgonists,LabaLamaCkd,PulmonRehab,Gluccorticds | LabaLama,BetaAgonists,LabaLamaCkd,PulmonRehab,Gluccorticds | LabaLamaIcs,BetaAgonists,LabaLamaCkd,PulmonRehab | LabaIcs,BetaAgonists,LabaLamaCkd,PulmonRehab | 1/Lama | ALL PROPOSED | ·· |
| 20 | ALL CDS-SUGGESTED | LabaLama,PulmonRehab,SmokingTher,FluVac,BetaAgonists,Gluccorticds | LabaIcs,PulmonRehab,SmokingTher,FluVac,BetaAgonists | Lama,PulmonRehab,SmokingTher,FluVac,BetaAgonists,Gluccorticds | ·· | 1/Laba+Lama | ALL PROPOSED | ·· |

##### Pulmonologist 5: Evaluation data

Table 21. **Use cases clinical context as collected from the EHR of pulmonologist 5**. Cell data with format m(n) where m,n are numbers denotes contextual use case data entered incorrectly into the EHR (represented outside the brackets) beside the contextual use case data given in the documentation (represented inside the brackets).

| **Use case ID** | **previous CAT score** | **previous mMRC score** | **previous number of exacerbations** | **active COPD treatment** | **current CAT score** | **current mMRC score** | **Current number of Exacerbations** | **Has asthma?** | **previous GOLD group** |
| --- | --- | --- | --- | --- | --- | --- | --- | --- | --- |
| *1* | 6 | 0 | 3 | Saba | 10 | 1 | 2 | FALSE | C |
| *2* | 29 (25) | 3 | 5 (4) | Saba (Laba+Lama+Ics) | 29 | 3 | 5 | FALSE | A |
| *3* | 17 | 1 | 1 | Laba | 20 | 2 | 4 | FALSE | B |
| *4* | 8 | 1 | 1 | Saba | 9 | 1 | 3 | TRUE | A |
| *5* | 18 | 2 | 1 | Lama | 26 | 3 | 3 | FALSE | B |
| *6* | 11 (6) | 1 | 0 | Saba | 11 | 1 | 0 | TRUE | A |
| *7* | 9 | 1 | 2 | Lama | 16 | 1 | 4 | FALSE | C |
| *8* | 25 | 3 | 1 | Lama | 27 | 4 | 2 | FALSE | B |
| *9* | 22 | 1 | 0 | Laba+Ics | 20 | 1 | 0 | FALSE | B |
| *10* | ·· | ·· | 0 | ·· | 12 | 1 | 0 | FALSE | ·· |
| *11* | 9 | 1 | 2 | Laba+Lama | 26 | 2 | 1 | FALSE | B(C) |
| *12* | 5 | 1 | 1 | Sama | 8 | 2 | 1 | FALSE | A |
| *13* | ·· | ·· | 0 | ·· | 6 | 1 | 0 (1) | TRUE | ·· |
| *14* | 20 | 1 | 1 | Laba | 21 | 2 | 3 | FALSE | B |
| *15* | 9 | 1 | 0 | Saba | 14 | 2 | 0 | FALSE | A |
| *16* | 8 | 0 | 0 | Laba+Ics | 9 | 1 | 3 | TRUE | A |
| *17* | 27 | 2 | 2 | Laba+Lama | 26 | 3 | 4 | FALSE | D |
| *18* | 6 | 1 | 2 | Laba+Ics | 7 | 1 | 4 | FALSE | C |
| *19* | 37 (32) | 3 (2) | 6(4) | Saba | 37 | 3 | 6 | FALSE | A (D) |
| *20* | 7 | 0 | 1 | Sama | 6 | 1 | 2 | TRUE | A |

Table 22. **Results from CDS service ‘copd-assess’ and input for CDS service ‘copd-careplan-review’, as collected from both the EHR of pulmonologist 5 and COPD-CDSS logs**. Keyword MISSING denotes the contextual data was not entered into the EHR.

| **use case ID** | **copd-assess result: GOLD group** | **user-selected GOLD group** | **copd-assess result: personalized COPD treatments** | **had annual influenza vaccine?** | **had pneumococcal vaccine?** | **age** | **is a smoker?** | **Has CKD?** | **Has CVD?** |
| --- | --- | --- | --- | --- | --- | --- | --- | --- | --- |
| 1 | D | D | [ [Laba+Lama], [Laba+Lama+Ics], [Laba+Ics],[Lama]] | FALSE | FALSE | 62 | TRUE | FALSE | TRUE |
| 2 | D | D | [ [Laba+Lama], [Laba+Lama+Ics], [Laba+Ics],[Lama]] | TRUE | TRUE | 75 | FALSE (TRUE) | FALSE (TRUE) | TRUE |
| 3 | D | D | [ [Laba+Lama], [Laba+Lama+Ics], [Laba+Ics],[Lama]] | FALSE | FALSE | 64 | FALSE | FALSE | FALSE |
| 4 | C | C | [[Lama],[Laba+Ics, Laba+Lama]] | FALSE | FALSE | 50 | TRUE | FALSE | FALSE |
| 5 | D | D | [ [Laba+Lama], [Laba+Lama+Ics], [Laba+Ics],[Lama]] | TRUE | TRUE | 62 | TRUE | FALSE | TRUE |
| 6 | B | B | [[Laba],[Lama],[Laba+Lama]] | FALSE | FALSE | 57 | TRUE | FALSE | FALSE |
| 7 | D | D | [ [Laba+Lama], [Laba+Lama+Ics], [Laba+Ics], [Lama]] | FALSE | FALSE | 68 | FALSE | FALSE (TRUE) | TRUE (FALSE) |
| 8 | D | D | [ [Laba+Lama], [Laba+Lama+Ics], [Laba+Ics], [Lama]] | TRUE | FALSE | 78 | FALSE | FALSE | TRUE |
| 9 | B | B | [ [Laba+Ics],[Laba+Lama], [Laba,Lama] ] | FALSE | FALSE | 69 | TRUE | FALSE | FALSE |
| 10 | B | B | [ [Laba], [Lama], [Laba+Lama] ] | FALSE | FALSE | 65 | FALSE | FALSE | TRUE |
| 11 | B | B | [ [Lama, Laba], [Laba+Lama]] | TRUE | FALSE | 71 | TRUE | FALSE (TRUE) | TRUE (FALSE) |
| 12 | A (D) | A (D) | [ [Saba+Sama, Saba], [Sama] ] | TRUE | FALSE | 76 | FALSE | FALSE | TRUE |
| 13 | A | A | [[Saba],[Saba+Sama,Sama]] | FALSE | FALSE | 42 | TRUE | FALSE | FALSE |
| 14 | D | D | [ [Laba+Lama], [Laba+Lama+Ics], [Laba+Ics], [Lama] ] | FALSE | FALSE | 59 | TRUE | FALSE | FALSE (TRUE) |
| 15 | B | B | [ [Laba], [Lama], [Laba+Lama] ] | TRUE | FALSE | 61 | FALSE | FALSE | FALSE |
| 16 | C | C | [ [Laba+Ics], [Laba+Lama], [Lama]] | FALSE | FALSE | 46 | FALSE | FALSE | FALSE |
| 17 | D | D | [ [Laba+Lama+Ics], [Laba+Ics], [Laba+Lama], [Lama] ] | TRUE | TRUE | 77 | TRUE | FALSE (TRUE) | FALSE |
| 18 | C | B | MISSING | FALSE | FALSE | 50 (62) | TRUE (FALSE) | FALSE | FALSE |
| 19 | D | D | [ [Laba+Lama], [Laba+Lama+Ics], [Laba+Ics],[Lama]] | TRUE | TRUE | 58 | FALSE | FALSE(TRUE) | TRUE |
| 20 | C | C | [ [Lama], [Laba+Lama, Laba+Ics]] | FALSE | FALSE | 55 | TRUE | FALSE | TRUE |

Table 23. **User-selected treatment(s) for CDS and corresponding results, with pulmonologist comments, for CDS service ‘copd-careplan-review’ and pulmonologist 5.**

| **use case ID** | **user-selected treatments** | **COPD care plan proposal 1** | **COPD care plan proposal 2** | **COPD care plan proposal 3** | **COPD care plan proposal 4** | **User-selected proposal/COPD treatment** | **Context-suitable recommendations** | **Pulmonologist comments** |
| --- | --- | --- | --- | --- | --- | --- | --- | --- |
| 1 | Laba+Lama | LabaLama,PulmonRehab,FluVac,Gluccorticds,BetaAgonists,SmokingTher | ·· | ·· | ·· | 1/Laba+Lama | ALL PROPOSED | ·· |
| 2 | ALL CDS_SUGGESTED | Lama,PulmonRehab,Gluccorticds,BetaAgonists | LabaLama,PulmonRehab,Gluccorticds,BetaAgonists | LabaIcs,PulmonRehab,BetaAgonists | LabaLamaIcs, BetaAgonists, PulmonRehab | 3/Laba+Ics | ALL PROPOSED | ·· |
| 3 | ALL CDS_SUGGESTED | LabaLamaIcs,PulmonRehab,FluVac | Lama,PulmonRehab,FluVac,Gluccorticds | LabaLama,PulmonRehab,FluVac,Gluccorticds | LabaIcs,FluVac,PulmonRehab | 4/Laba+Ics | ALL PROPOSED | ·· |
| 4 | ALL CDS_SUGGESTED | Lama,PulmonRehab,FluVac,SmokingTher,Gluccorticds | LabaLama,PulmonRehab,FluVac,SmokingTher,Gluccorticds | LabaIcs,PulmonRehab,FluVac,SmokingTher | ·· | 2/Laba+Lama | ALL PROPOSED | ·· |
| 5 | ALL CDS_SUGGESTED | LabaLamaIcs,PulmonRehab,SmokingTher,BetaAgonists | LabaIcs,PulmonRehab,SmokingTher,BetaAgonists | LabaLama,PulmonRehab,SmokingTher,Gluccorticds,BetaAgonists | Lama,PulmonRehab,Gluccorticds,SmokingTher, BetaAgonists | 2/Laba+Ics | ALL PROPOSED | ·· |
| 6 | ALL CDS_SUGGESTED | Lama,PulmonRehab,SmokingTher,Gluccorticds, FluVac | LabaLama,PulmonRehab,SmokingTher,Gluccorticds, FluVac | Laba,PulmonRehab,SmokingTher,Gluccorticds, FluVac | ·· | 2/Laba+Lama | ALL PROPOSED | ·· |
| 7 | ALL CDS_SUGGESTED | LabaLamaIcs,PulmonRehab, FluVac,BetaAgonists,PneumnVac | LabaIcs,PulmonRehab, FluVac,BetaAgonists,PneumnVac | LabaLama,PulmonRehab, FluVac,PneumnVac,BetaAgonists,Gluccorticds | Lama,PulmonRehab, FluVac,BetaAgonists,PneumnVac,Gluccorticds | 2/Laba+Ics | ALL PROPOSED | ·· |
| 8 | ALL CDS_SUGGESTED | LabaLama,PulmonRehab,BetaAgonists,PneumnVac,Gluccorticds | LabaIcs,PulmonRehab,BetaAgonists,PneumnVac | LabaLamaIcs,PulmonRehab,BetaAgonists,PneumnVac | Lama,PulmonRehab,BetaAgonists,PneumnVac,Gluccorticds | 1/Laba+Lama | ALL PROPOSED | ·· |
| 9 | ALL CDS_SUGGESTED | Lama,PulmonRehab,SmokingTher,FluVac,Gluccorticds,PneumnVac | LabaLama,PulmonRehab,SmokingTher,FluVac,Gluccorticds,PneumnVac | LabaIcs,PulmonRehab,SmokingTher,FluVac,PneumnVac | Laba,PulmonRehab,SmokingTher,FluVac,PneumnVac,Gluccorticds | 2/Laba+Lama | ALL PROPOSED | ·· |
| 10 | ALL CDS_SUGGESTED | LabaLama,PulmonRehab,Gluccorticds, FluVac, PneumnVac,BetaAgonists | Lama,PulmonRehab,Gluccorticds, FluVac, PneumnVac, BetaAgonists | Laba,PulmonRehab,Gluccorticds, FluVac, PneumnVac | ·· | 1/Laba+Lama | ALL PROPOSED | ·· |
| 11 | ALL CDS_SUGGESTED | Laba,PulmonRehab,SmokingTher,PneumnVac,Gluccorticds | Lama,PulmonRehab,SmokingTher,PneumnVac,Gluccorticds, BetaAgonists | PulmonRehab,SmokingTher,PneumnVac,Gluccorticds,LabaLama,BetaAgonists | ·· | 3/Laba+Lama | ALL PROPOSED | ·· |
| 12 | ALL CDS_SUGGESTED | Saba,PulmonRehab,PneumnVac | SabaSama,PulmonRehab,PneumnVac,BetaAgonists | Sama,PulmonRehab,PneumnVac,BetaAgonists | ·· | 2/Saba+Sama | ALL PROPOSED | ·· |
| 13 | ALL CDS_SUGGESTED | Saba,PulmonRehab,FluVac, SmokingTher | Sama,PulmonRehab,FluVac,SmokingTher | SabaSama,PulmonRehab,FluVac,SmokingTher | ·· | 1/Saba | ALL PROPOSED | ·· |
| 14 | ALL CDS_SUGGESTED | Lama,PulmonRehab,SmokingTher,FluVac,Gluccorticds | LabaLamaIcs,PulmonRehab,SmokingTher,FluVac | LabaLama,PulmonRehab,SmokingTher,FluVac,Gluccorticds | LabaIcs,PulmonRehab,SmokingTher,FluVac | 3/Laba+Ics | ALL PROPOSED | ·· |
| 15 | ALL CDS_SUGGESTED | Lama,PulmonRehab,Gluccorticds | LabaLama,PulmonRehab,Gluccorticds | Laba,PulmonRehab,Gluccorticds | ·· | 2/Laba+Lama | ALL PROPOSED | ·· |
| 16 | ALL CDS_SUGGESTED | Lama,PulmonRehab,FluVac,Gluccorticds | LabaLama,PulmonRehab,FluVac,Gluccorticds | LabaIcs,PulmonRehab,FluVac | ·· | 3/Laba+Ics | ALL PROPOSED | ·· |
| 17 | ALL CDS_SUGGESTED | LabaLama,PulmonRehab,Gluccorticds,SmokingTher | Lama,PulmonRehab,SmokingTher,Gluccorticds | LabaIcs,PulmonRehab,SmokingTher | LabaIcs,PulmonRehab,SmokingTher | 3/Laba+Ics | ALL PROPOSED | ·· |
| 18 | Lama, Laba+Lama, Laba+Ics, Laba | Lama,PulmonRehab,Gluccorticds,FluVac | LabaLama,PulmonRehab,Gluccorticds,FluVac | LabaIcs,PulmonRehab,FluVac | Laba,PulmonRehab,FluVac,Gluccorticds | 3/Laba+Ics | ALL PROPOSED | ·· |
| 19 | ALL CDS_SUGGESTED | Lama,BetaAgonists,PulmonRehab,Gluccorticds | LabaLama,BetaAgonists,PulmonRehab,Gluccorticds | LabaIcs,BetaAgonists,PulmonRehab | LabaLamaIcs,BetaAgonists,PulmonRehab | 4/Laba+Lama+Ics | ALL PROPOSED | ·· |
| 20 | ALL CDS_SUGGESTED | Lama,PulmonRehab,SmokingTher,FluVac,BetaAgonists,Gluccorticds | LabaIcs,PulmonRehab,SmokingTher,FluVac,BetaAgonists | LabaLama,PulmonRehab,SmokingTher,FluVac,BetaAgonists,Gluccorticds | ·· | 2/Laba+Ics | ALL PROPOSED | ·· |

##### B.3 Responses from evaluation questionnaire

Table 24. **COPD-CDSS evaluation questionnaire and responses from the five pulmonologists**.

|  | **Respondents** | | | | |
| --- | --- | --- | --- | --- | --- |
|  | Dr Miljan Ćulafić | Dr Vesna Dopuđa Pantić | Dr Draško Dubljanin | Dr Danica Ivković | Dr Vladimir Žugić |
| DSS interaction data | RR398 - DOCTOR 2: patients 1-2 contacts created on 15.09.2021, patients 3-20 contacts created on 21.09.2021 | RR326 - DOCTOR 1: patients 1-20 contacts were created on September 28, 2021 and again on October 1, 2021 for everyone. Contact is missing only for Patient 19 | RR1686 - DOCTOR 3: patients 1-15 created on 08.09.2021, patients 15???-20 created on 15.09.2021 | RR1296 - DOCTOR 4: created on 15.09.2021 | RR1684 - DOCTOR 5: patients 1-20 created on September 22, 2021 |
| How many COPD patients do you see on an average workday in the clinic? | 5 – 15 | 5 – 15 OR More than 15 | 5 – 15 | More than 15 | 5 – 15 |
| How often and in what context would you use the COPD Decision Support System (COPD DSS)? Please describe the situations in which the COPD DSS is most useful to you. | The COPD DSS is a good idea as a reminder to younger doctors who have just started outpatient work as a reminder from the guidelines. | For faster and easier orientation in the assessment of therapeutic options concerning the severity of the disease. | Relatively often, in the context of deciding on therapy. Useful when deciding on therapy in exacerbations of the disease. | In situations when the patient comes regularly for examinations, for better monitoring. | The system is a very good idea and a kind of guideline for doctors with little clinical experience at the very beginning of independent management of patients, and then their outpatient examinations. I think it is a very good reminder of all therapeutic modalities. |
| How else do you think you could use the COPD DSS (in the sense of ways, places and scenarios for use, interactions with other conditions etc.)? | Again, it can serve as an advising tool and as a reminder of other guidelines if they exist in other diseases (possibly to indicate drug interactions...) | Possibility of continuous monitoring of the patient from different institutions that are connected by the system. | Decision support systems on bronchial asthma and pneumonia can be considered (intake of inflammatory parameters, duration, swabs, comorbidities, inpatient and outpatient treatments). | To make something similar available to doctors in PHC Centres, for better monitoring of patients | It would not be bad to expand the field of action to other specialities (gastroenterology, endocrinology, cardiology...) and thus help in deciding the patient with comorbidities initially during outpatient consultations. |
| What are your goals and expectations when using the COPD DSS? Does the COPD DSS help you achieve your goals and if so, how? | The goal of modern medical treatment is a personalized approach to the patient, with a consequent specific medication. Therefore, this program can only serve as an advisory tool. | To improve the efficiency of work with a more precise assessment of the severity of the disease and the possibility of considering therapeutic options. | Goal: giving the most optimal therapy and eliminating subjectivity in decision making. It is achievable if all valid and relevant information is entered. | To be accessible and as easy to use as possible. | Reduce the time of re-evaluation of the patient's condition, provide a quick reminder of previous visits and accurate overview of previous medications and therapies as well as patients' clinical responses for better clinical decisions. |
| What do you think are the leading alternatives to the COPD DSS and why? In what aspects is the DSS better than alternatives and vice-versa? | I have not heard of an alternative to this system. | There are no alternatives to the COPD DSS in our health system. | I have not seen an alternative to this system. | For now, I would focus on the development of this system | I am unaware of the existence of alternative systems. |
| What positive experiences or outcomes have you had in using the COPD DSS? | Offered options for medication, based on the entered parameters on severity and discomfort of disease. Also, the idea of non-medical recommendations. | Easy to use and for decision making. | Clear and concise software (system). | More options for thinking and caring for patients. | The moment of offering the right therapeutic regimen for the patient based on previous visits is certainly purposeful and helpful. |
| What issues, concerns or problems have you had with the DSS? | Except for initial challenges in entering data, without problems and questions. | There is no therapeutic option for inhaled corticosteroids in deciding on a therapeutic option when entering data for patients who also have asthma; There are not enough options for entering the data on associated diseases of importance; Nephrotic syndrome is not the same as HBI 3rd or 4th degree; In several patients, the system changed the selected group of COPD and returned it to the level of the previously set group; For patient under No. 20, the system does not allow the completion of data entry…. | Minor technical problems that were easily resolved in consultations and with the support of the IT specialist. | Just technical, until I figured out how the system works | No major problems. |
| What features of the COPD DSS do you find the most useful? | The fact that types of requested data serve as a basis for further recommendations on medication (therapeutic options). | The choices for therapeutic options. | Recommendations for medication, drop-down menus, fast data entry. | Therapy (medication) suggestions and other options for the future actions. | The features that process the entered information after which the therapeutic modality is offered. |
| What features of the COPD DSS do you find the least useful? | None | None | None | None | I have not come across such features. |
| How can the COPD DSS be improved (e.g. user interface, navigation through the DSS, functionalities of the DSS, appearance, provided recommendation and the way they are presented, speed while working with it etc.)? | In outpatient work, e.g. integration with the existing hospital information system, so that all required data are entered is to be transferred in the doctor's report on the patient, but also to serve as a basis for further diagnosis and therapy (medication) recommendations. Avoiding the unnecessary entering of the same data is a waste of time. | The data must be more precise. E.g., drugs from the group of beta-agonists are divided into SABA and LABA, and it must be clear to which therapeutic option the data refers to and that the use of drugs from the group of beta-agonists is not advised. | The questionnaires should be on the first page, without the opening of new pages. | To get the data with as few clicks as possible. | I think that a little time was spent with this system (ed.: by me) for a bigger analysis and therefore some more serious proposals. The goal of every system is simplicity, speed and efficiency, with as few unnecessary pop-ups and additional questions as possible! A basic range of colours that would classify the most important actions is something that is certainly desirable. |
| What would make you more inclined to use the COPD DSS? What features or functions would you like to see developed further or added and why? | Possibly with reminders where diagnostic methods, which are indicated for patients, can be performed outside the institution where he/she is currently treated, or with a short reminder with the summary on a specific drug because there are many parallels in the drug market. | More efficient work, faster orientation in assessing the severity of the disease and choosing a therapy. | Functions of including other aspects (findings) such as spirometry, lab tests, X-rays. | As much patient data as possible should be entered into the system. | Earlier mentioned, permeation of information and offered therapies for patients with comorbidities /e.g. COPD + CMP + HBI / after which the real challenge would be to optimize the therapy with an overall perspective of such patients! |
| On a scale of 1 to 5, please rate your satisfaction with the COPD Decision Support System (COPD DSS). | 4 | 4 | 4 | 4 | 4 |
| Please provide a reason for (elaborate) your score in the previous question. | Nice try to create a reminder to make things easier for doctors. Certainly, upgrades and additions will have to be done. | Initial (added to clarify) data entry problems affected the overall satisfaction rating. The offered options that we do not agree with as well as other options that are offered after this, such as the therapy, are a bit confusing (translation to be confirmed?). | During the examination, the maximum time must be used, which is limited to 15 minutes per patient (in terms of the clinical workflow?). It seems to me that the time required to enter data is longer than the time available. Heliant (hospital patient record software) lacks the "one-click to desired goal" option. | ·· | I like the idea and the pilot project; the time ahead will show the extent to which it will come to life. |
| What do you think of the COPD DSS clinical performance? Please rate on a scale of 1 to 5. | 4 | 4 | 4 | 4 | 4 |
| What do you think of the COPD DSS technical performance (e.g. stability, speed)? Please rate on a scale of 1 to 5. | 5 | 4 | 3 | 4 | 5 |
| What do you think of the COPD DSS design (e.g. interface, navigation, functionality, appearance)? Please rate on a scale of 1 to 5. | 4 | 4 | 4 | 4 | 4 |
| Is there anything else you would like to share about the COPD DSS? | ·· | ·· | ·· | ·· | ·· |

#### Appendix C: Technical Documentation

#### General Information

##### System Overview

The COPD-CDS system is a cloud-based system that enables users to request support when managing the COPD of a patient via an electronic health record (EHR) system. Notifications for decision support comply with HL7 CDS Hooks specifications. Results from COPD-CDS are HL7-FHIR compliant.

The COPD-CDS system supports COPD management in two stages: first, the system assesses the COPD severity of the patient using the context data which is part of the client’s request call under hook with Id ‘copd-assess’. Next, the system reviews the patient’s COPD treatment and provides a collection of care plan proposals to update the patient’s COPD treatment pathway. This is done as part of hook ‘copd-careplan-review’.

The COPD-CDS system is composed of a collection of microservices to handle the context data included in an EHR system request for support, to enact computer-interpretable guidelines (CIGs), and to identify potential conflicting interactions among recommendations within a CIG (which it may be composed of one or more parts of distinct CIGs). Additionally, there is an argumentation-based mitigation service to convert a (possibly merged) CIG into a set of safe care plan proposals along with the rationale for including each recommendation as part of the care plan (personalised care planning).

##### Authorised Use Permission

This document and the corresponding COPD-CDS system are part of ROAD2H: Resource Optimisation, Argumentation, Decision Support and Knowledge Transfer to create Value via Learning Health Systems (<https://gow.epsrc.ukri.org/NGBOViewGrant.aspx?GrantRef=EP/P029558/1>). Access to the CDS system is limited and permission must be sought before making copies. Please contact the relevant department for more information.

#### Points of contact

##### Information

| Professor Brendan Delaney | (co-investigator) | |
| --- | --- | --- |
| Professor Francesca Toni | (co-Investigator) | |
| Professor Vasa Ćurčin | (co-investigator) | |
| Doctor Jesús Domínguez | (researcher co-investigator) | |
| Doctor Kristijonas Čyras | (researcher co-investigator) | |

##### Coordination

All queries regarding installation or use of the COPD-CDS system should be addressed to:

Doctor Jesús Domínguez

#### Acronyms and Abbreviations

| CDS | Clinical Decision Support |
| --- | --- |
| CDS-HsM | CDS Hooks Manager microservice |
| CDS-SsM | CDS Services Manager microservice |
| EHR | Electronic Health Records |
| COPD | Chronic Obstructive Pulmonary Disease |
| GOLD | Chronic Obstructive Pulmonary Disease |
| CIG | Computer-Interpretable Guideline |

##### System Concepts

This section describes the features of the COPD-CDS system, and the technologies used to build it.

###### Available CDS services

- **Hook `copd-assess`:** this hook is triggered when the pulmonologist has completed the airflow limitation severity assessment on the current patient with COPD. The hook context contains pairs of COPD measurements for CAT and mMRC dyspnoea scale scores, one pair taken from the patient record from a previous COPD assessment visit (if a previous visit exists) and another of measurements taken on the current encounter. Similarly for the number of COPD exacerbations. Additionally, the context includes information on whether asthma is present in the patient as well as the COPD diagnosis from the previous COPD visit (if such visit exists), that is, the identified COPD group and active COPD treatment. The result suggests a GOLD 2017 COPD group (that is, A, B, C, or D) and a list of preferentially ordered COPD treatments, both outcomes are personalised for the COPD patient at hand.
- **Hook `copd-careplan-review`:** this hook is triggered when the pulmonologist has assessed the COPD severity of the patient, by assigning/confirming a GOLD 2017 COPD group as well as selecting one or more COPD treatments for care planning decision support. The hook context contains the GOLD 20017 COPD group selected by the clinician, along with two collections of COPD drug types, or drug type combinations, one collection includes the treatments proposed by the pulmonologist alongside the patient; the other collection includes the treatment preferences as suggested by the COPD-CDS system as a response to hook `copd-assess` on the active patient. Additionally, there is some lifestyle and demographics information, and comorbidities and immunization status data. The response of the COPD-CDS system is composed of a collection of personalised care plan proposals, based on GOLD 2017, where each proposal contains non-conflictive clinical recommendations that have been selected taking into account the pulmonologist/patient preferences on COPD treatments, as well as other factors such as immunization, lifestyle -e.g., smoking status- and active comorbidities present in the patient's record, in particular cardiovascular and chronic kidney diseases, which could potentially interfere with some COPD treatments.

###### System Specification

The technologies used to construct the COPD-CDS system are

- Node.js v16.17
- Docker v4.13.0
- MongoDB v6.0
- SWI-Prolog v8.4.3
- Apache Jena-Fuseki v4.6.1
- SPARQL v1.1
- HL7 CDS Hooks v2.0
- HL7 FHIR v4.3.0
- SNOWSTORM by SNOMED International v7.11.0
- Python v3.4.0

#### Getting Started

##### System Requirements

Most of the system -excluding the mitigation service which it is currently live, running at a Heroku server address- has been dockerised (the process of packing, deploying, and running an application using Docker containers) so that there is no specific requirement for distinct operating systems. The software can run on machines with potentially any operating system installed, assuming the Docker desktop app is available for download for said operating system.

- Docker desktop can be downloaded here: <https://www.docker.com/products/docker-desktop/> .
- To simulate API request calls to the COPD-CDS system, download Postman v10 or higher here: <https://www.postman.com/>.
- To clone the source code from the Github repository, download git here: <https://git-scm.com/>.
- The COPD-CDS system repository is available here: <https://github.com/susoDominguez/COPD-CDS>
- The CDS-HsM repository is available here: <https://github.com/susoDominguez/cds-services-manager>
- The TMR-based CDS-SsM repository is available here: <https://github.com/susoDominguez/cds_hooks_manager>
- The TMR-based authoring tool, that is, the Interaction, Store and Reasoner microservices, is available here <https://github.com/susoDominguez/TMRWebX>
- The argumentation-based conflict resolution service is available here: <https://github.com/susoDominguez/ABAPlusG>
- The mapping from TMR terms (and `copd-assess` structured results, since there is no CIG involved) to FHIR instances is available here: <https://github.com/susoDominguez/TMR2FHIRconverter>
- The TMR ontologies and COPD CIG file are available here: <https://github.com/susoDominguez/TMR-CIG-COPD>
- The CDS hooks specifications, with examples, for `copd-assess` and `copd-careplan-review` are available here: <https://github.com/susoDominguez/ROAD2H-hooks>
- The MongoDB-based hook processing documents for `copd-assess` and `copd-careplan-review` are available here: <https://github.com/susoDominguez/mongodb-hook-processing-collections.git>
- The SNOMED CT server installation can be found here: <https://github.com/IHTSDO/snowstorm>. We access it using their FHIR API.

##### Installation

1. Download and Install Docker and Git.
2. Clone the COPD-CDS repository to your computer. This repository contains a docker-compose file to automate the build and running of the applications involved in the system.

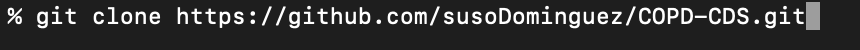

(Other cloning options are available from git)

3. change to the COPD-CDS directory
4. Inside the COPD-CDS folder do
   a. Clone the TMR-based implementation of the CGs authoring service suite

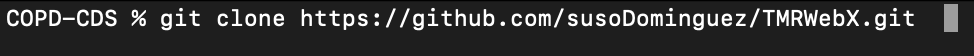

b. Clone the CDS-HsM repository

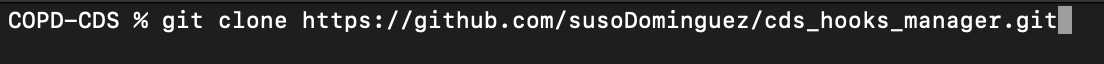

c. Clone the TMR-based implementation of the ABA+G formalism

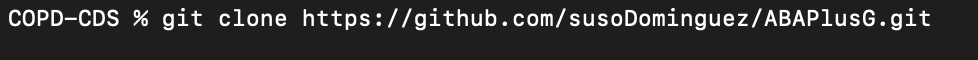

d. Clone the CDS-SsM repository

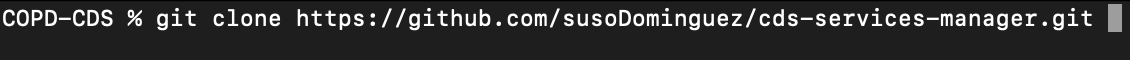

5. Let’s add the CDS Hooks processing documents to the CDS-HsM
   a. Go to the CDS-HsM directory
   b. Clone the processing documents repository into CDS-HsM

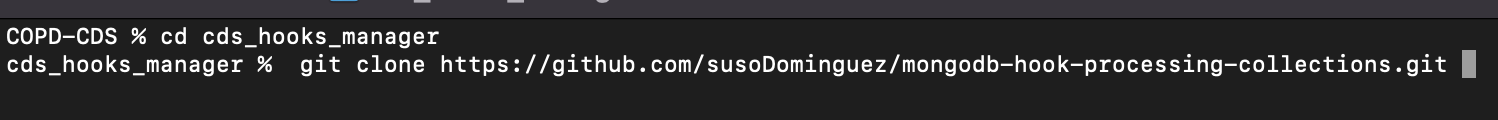

6. Let’s add the TMR schema and ontologies to the TMR-based authoring tool.
   a. Go back to the main folder of COPD-CDS
   b. Go to the reasoner microservice directory (/TMRWebX/backend/)
   c. Clone the TMR ontology

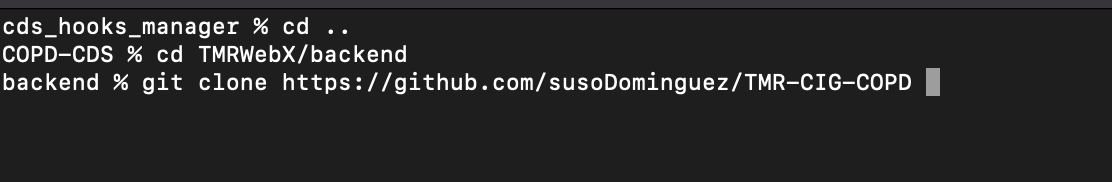

d. Go back to the COPD-CDS directory

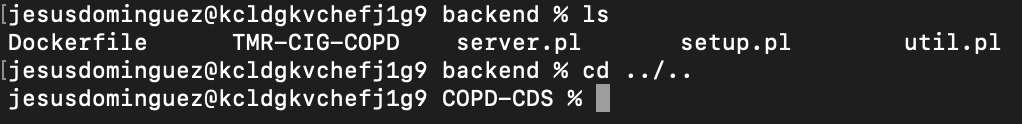

7. Next, let’s add the TMR to FHIR converter repository to the CDS-SsM
   a. Go to the FHIR_converter_module folder within the CDS-SsM.
   b. Clone the TMR2FHIRconverter repository.

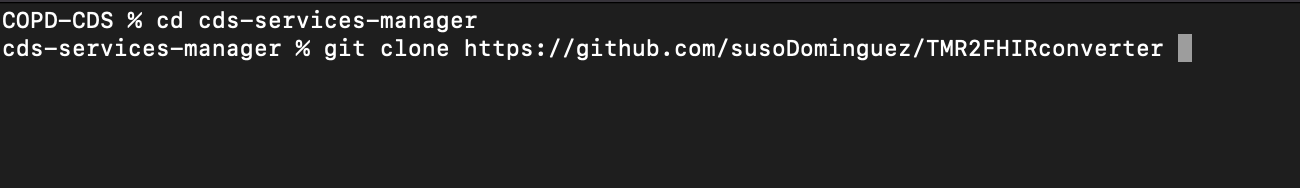

c. Return to the COPD-CDS directory to launch the application

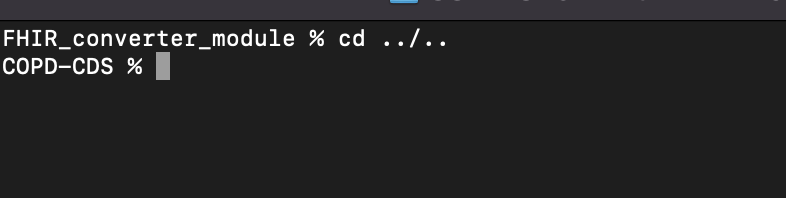

8. Now we are ready to build and launch the COPD-DSS.

#### Configuration

The collection of microservices is built automatically using an environment file within the COPD-CDS directory. Below, we provide the set of variables, and default values, that must be added to the environment file. Values can be changed by the user if required. A brief description of the semantics of each variable can be found after the assignment of values to variables.

**MONGODB_PORT=**27017

**CDS_HM_PORT=**3001

**CDS_SM_PORT=**3010

**SNOMEDCT_FHIR_SRVR_URL=**snowstorm-fhir.snomedtools.org

**CDS_SM_LOGS=**cds_sm_logs

**CDS_HM_LOGS=**cds_hm_logs

**CIG_INTERACTION_PORT=**8888

**CIG_STORE_PWD=**road2h

**CIG_STORE_PORT=**3030

**ARGUMENTATION_PORT**=5000

- **MONGODB_PORT:** port number of the MongoDB server
- **CDS_HM_PORT:** port number of the CDS-HsM
- **CDS_SM_PORT** port number of the CDS-SsM
- **SNOMEDCT_FHIR_SRVR_URL:** URL of the SNOMED CT FHIR API server. For testing purposes, we have provided the sandbox offered by the team at SNOWSTORM. If you would like to install your own SNOMED CT server and add its FHIR API URL instead, follow the instructions at <https://github.com/IHTSDO/snowstorm>
- **CDS_SM_LOGS:** name of the MongoDB collection to store CDS-SsM logs
- **CDS_HM_LOGS:** name of the MongoDB collection to store CDS-HsM logs
- **CIG_INTERACTION_PORT:** port number of the CIG Interaction microservice
- **CIG_STORE_PWD:** password to access the Jena/Fuseki server with ADMIN rights as part of the STORE microservice
- **ARGUMENTATION_PORT:** port number of the conflict resolution service

Now, create the environment file with name **.env** and use your favourite text editor to add the set of variables and corresponding values into the folder of COPD-CDS.

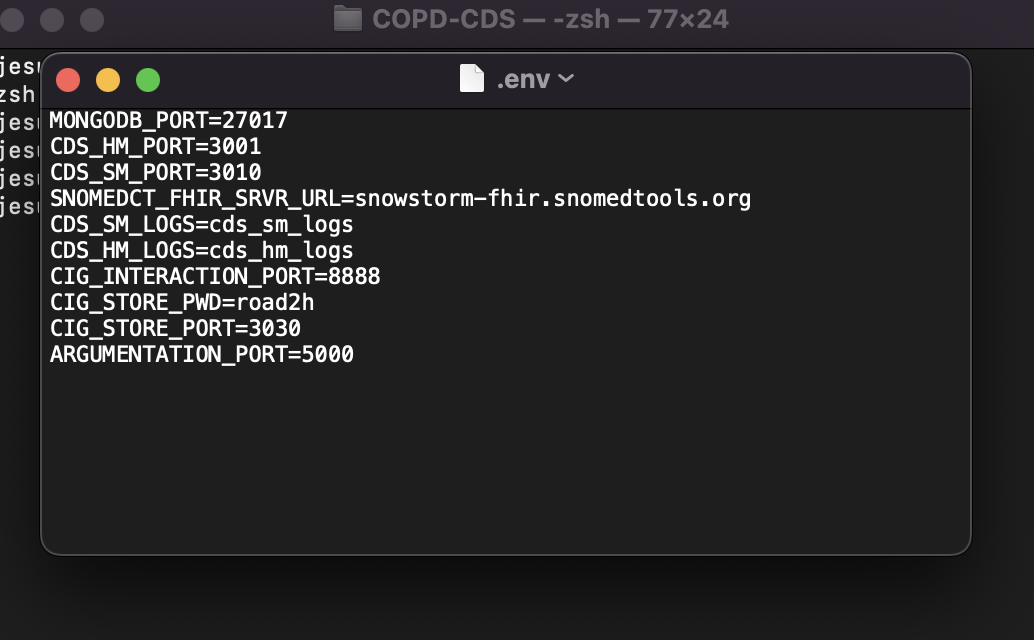

Finally, launch the COPD-CDS system:

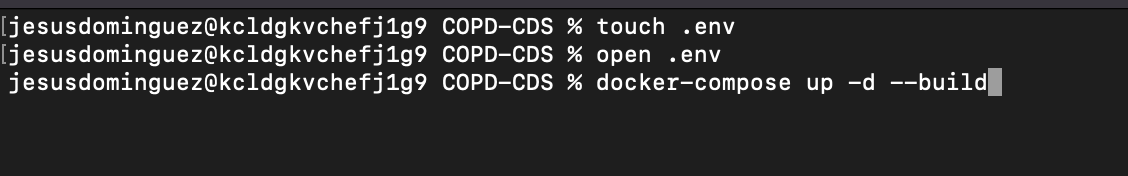

Then, all the services should be up and running.

Figure 1. Illustration depicting the collection of microservices, and conflict resolution service involved in the COPD-CDS system.

#### Loading TMR Knowledge into the Store microservice

The final step is to load the TMR ontologies into the Store microservice. To do that, in your browser go to <http://localhost:3030/>. When asked for credentials, the username is **admin** and the password is the one given to variable **CIG_STORE_PWD.** You will find the server with 4 empty datasets:

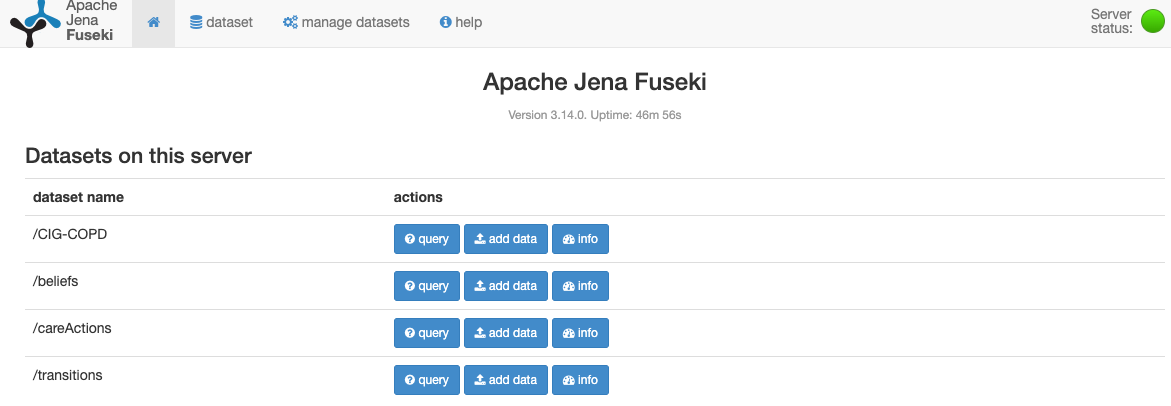

- In the row with dataset name **/CIG-COPD,** click on *add data* button then on *select files* button in the next page. The file is in *COPD-CDS/TMRWebX/backend/TMR-CIG-COPD/ontologies/guidelines/CIG-COPD.trig.* Open the file, then select *upload all* button.
- Similarly for dataset **/beliefs**, which has its corresponding file in *COPD-CDS/TMRWebX/backend/TMR-CIG-COPD/ontologies/beliefs_shorten.trig.*
- Similarly for dataset **/*careActions***, which has its corresponding file in *COPD-CDS/TMRWebX/backend/TMR-CIG-COPD/ontologies/careActions.trig.*
- Similarly for dataset **/transitions**, which has its corresponding file in *COPD-CDS/TMRWebX/backend/TMR-CIG-COPD/ontologies/transitions.trig.*

The system installation is now finished.

We have provided an additional microservice, mongo-express web server, that is linked to the MongoDB database and can be used to view/modify the collection of CDS hooks processing documents applied to both hooks. The web server is located by default in <http://localhost:8081/>.

### Using the COPD-CDS System

To request support from the COPD-CDS system, a notification must be sent using the CDS hooks specifications for hooks `copd-assess’ and `copd-careplan-review`.

CDS hooks specifications, with examples, for both hook contexts with Ids `copd-assess’ and `copd-careplan-review` alongside JSON-based input files for each of the use cases and hooks can be found here: <https://github.com/susoDominguez/ROAD2H-hooks>.

Next, we demonstrate how to launch each of the hooks with the JSON-based input files taken from the repository stated above.

#### Launch

##### Hook ‘copd-assess’

To invoke CDS for hook with Id ‘copd-assess’ and clinical workflow context from use case 1, we must make a request call as follows:

curl --location --request POST '127.0.0.1:3001/cds-services/copd-assess' \

--header 'Content-Type: application/json' \

--data-raw 'useCase_1.json'

where *useCase_1.json* is the file with the JSON document to be used as input for the COPD severity assessment of the first use case. The document is excessive in length to have it added on this document, but it is stored in the ROAD2H-hooks repository, in directory `ROAD2H use cases/copd-assess hook contexts`.

The response from the COPD-CDS system is a CDS suggestion card with the following (abbreviated) information:

{

"cards": [{

"summary": "Assessment of COPD",

"indicator": "info",

"source": {

"label": "GOLD 2017 COPD assessment"

},

"suggestions": [{

"label": "COPD assessment decision support",

"uuid": "00000001677163",

"actions": [{

"type": "update",

"description": "Update COPD assessment interface",

"resource": {

"resourceType": "Parameters",

"id": "copdAssessParameters",

"parameter": [{

"name": "patient",

"valueId": "1677163"

},

{

"name": "medicationBundle",

"resource": {

"resourceType": "Bundle",

"id": "medicationBundle",

"type": "collection",

"entry": [{

"resource": {

"resourceType": "Medication",

"id": "DrugCatLabaLama",

"code": {

"coding": [{

"code": "LabaLama",

"display": "medication containing a combination of LABA and LAMA"

}]

}

}

},{…},{…},{…},{…},{…},{…},

{

"resource": {

"resourceType": "Medication",

"id": "DrugCatLabaLamaIcs",

"code": {

"coding": [{

"code": "LabaLamaIcs",

"display": "medication containing a combination of LABA, LAMA and ICS"

}]

}

}

}

]

}

},

{

"name": "assessedCopdStage",

"valueCoding": {

"system": "http://snomed.info/sct",

"code": "1097901000000101"

}

},

{

"name": "copdGroupA",

"part": […]

},

{

"name": "copdGroupB",

"part": […]

},

{

"name": "copdGroupC",

"part": […]

},

{

"name": "copdGroupD",

"part": [{

"name": "code",

"valueCoding": {

"system": "http://snomed.info/sct",

"code": "1097901000000101",

"display": "Global Initiative for Chronic Obstructive Lung Disease 2017 group D"

}

},

{

"name": "medicationPreference_1",

"resource": {

"resourceType": "List",

"id": "list1",

"status": "current",

"mode": "changes",

"entry": [{

"item": {

"reference": "Medication/DrugCatLabaLama"

}

}]

}

},

{

"name": "medicationPreference_2",

"resource": {

"resourceType": "List",

"id": "list2",

"status": "current",

"mode": "changes",

"entry": [{

"item": {

"reference": "Medication/DrugCatLabaLamaIcs"

}

}]

}

},

{

"name": "medicationPreference_3",

"resource": {

"resourceType": "List",

"id": "list3",

"status": "current",

"mode": "changes",

"entry": [{

"item": {

"reference": "Medication/DrugCatLabaIcs"

}

}]

}

},

{

"name": "medicationPreference_4",

"resource": {

"resourceType": "List",

"id": "list4",

"status": "current",

"mode": "changes",

"entry": [{

"item": {

"reference": "Medication/DrugTLama"

}

}]

}

}

]

}

]

}

}]

}],

"selectionBehaviour": "at-most-one"

}]

}

Observe the card contains parameters to describe the patient identifier, the collection of drug types and drug type combinations for COPD, the personalised GOLD group (labelled as assessedCopdStage), and four parameters denoting each GOLD group (labelled as copdGroupA, copdGroupB, copdGroupC, and copdGroupD for GOLD group A – D, respectively). The personalised GOLD group is the result of the COPD severity symptom assessment using both current and previous measurements taken by the pulmonologist. Each parameter denoting a GOLD group contains the SNOMED CT term that represents said GOLD group and a list of ordered COPD drug types. Drug types have a priority order which follows the GOLD guideline algorithm for selecting suitable COPD treatment pathways for COPD patients. Although parameter with label assessedCopdStage identifies the current GOLD group of the patient, the COPD-CDS system provides a COPD treatment priority order for all four GOLD groups using the same clinical workflow context, to provide all available information to the pulmonologist while they verify/confirm the COPD-CDS system response.

The patient in this use case has been assessed by the COPD-CDS system as belonging to GOLD group D. Parameter copdGroupD contains four COPD treatments pathways, each labelled as medicationPreference_n where n is the COPD treatment preference level, ranging from 1 -the most preferred- to 4 -the least preferred-. Then, each medicationPreference_n contains an unordered list of references to FHIR medication instances in this CDS card, under resource labelled as medicationBundle. Accordingly, medicationPreference_1 has solely one reference, to the FHIR medication denoting a combination of LABA and LAMA bronchodilators; medicationPreference_2 has also a single reference, to the FHIR medication denoting a combination of LABA and LAMA and ICS bronchodilators; medicationPreference_3 has a single reference to the FHIR medication denoting a combination of LABA and ICS bronchodilators; and medicationPreference_4 has a single reference to the FHIR medication denoting a bronchodilator of type LAMA.

##### hook ‘copd-careplan-review’

To invoke CDS for hook with Id ‘copd-careplan-review’ and clinical workflow context from use case 1, we must make a request call as follows:

curl --location --request POST

'127.0.0.1:3001/cds-services/copd-careplan-review/cigModel/tmr' \

--header 'Content-Type: application/json' \

--data-raw 'useCase_1.json'

where *useCase_1.json* is the file with the JSON document to be used as input for the COPD treatment management of the first use case. The document is excessive in length to have it added on this document, but it is stored in the ROAD2H-hooks repository, in directory `ROAD2H use cases/copd-careplan-review hook contexts`.

The response from the COPD-CDS system is a CDS suggestion card with the following entries:

###### Medication resources

Medication resources in the context of the COPD-CDS system represent the application of care actions and include both the vaccines and the collection of COPD drug types or type combinations which are active in any of the care plan proposals suggested by the COPD-CDS system. For this iteration of the project, clinical terms returned to the CDS client were not identified by structured clinical vocabularies like SNOMED CT, but by the internal codes used by the TMR ontology.

{

"resource": {

"resourceType": "Medication",

"id": "DrugTFluVac",

"code": {

"coding": [

{

"system": "http://anonymous.org/data/DrugTFluVac",

"code": "FluVac",

"display": "administer influenza vaccine"

}

]

}

}

},

{

"resource": {

"resourceType": "Medication",

"id": "DrugTIcs",

"code": {

"coding": [

{

"system": "http://anonymous.org/data/DrugTIcs",

"code": "Ics",

"display": "administer inhaled corticosteroids"

}

]

}

}

},

{

"resource": {

"resourceType": "Medication",

"id": "DrugCatLabaIcs",

"code": {

"coding": [

{

"system": "http://anonymous.org/data/DrugCatLabaIcs",

"code": "LabaIcs",

"display": "administer a combination of LABA and ICS"

}

]

}

}

},

{

"resource": {

"resourceType": "Medication",

"id": "DrugTLama",

"code": {

"coding": [

{

"system": "http://anonymous.org/data/DrugTLama",

"code": "Lama",

"display": "administer LAMA"

}

]

}

}

},

{

"resource": {

"resourceType": "Medication",

"id": "DrugCatLabaLama",

"code": {

"coding": [

{

"system": "http://anonymous.org/data/DrugCatLabaLama",

"code": "LabaLama",

"display": "administer a combination of LABA and LAMA"

}

]

}

}

},

{

"resource": {

"resourceType": "Medication",

"id": "DrugCatBetaAgonist",

"code": {

"coding": [

{

"system": "http://anonymous.org/data/DrugCatBetaAgonist",

"code": "BetaAgonist",

"display": "administer Beta Agonists when cardiovascular disease is present"

}

]

}

}

},

{

"resource": {

"resourceType": "Medication",

"id": "DrugCatLabaLamaIcs",

"code": {

"coding": [

{

"system": "http://anonymous.org/data/DrugCatLabaLamaIcs",

"code": "LabaLamaIcs",

"display": "administer a combination of LABA, LAMA and ICS"

}

]

}

}

}

Medications are referenced by FHIR medicationRequest resources, which we illustrate after introduction other canonical resources that are also included in a medicationRequest.

Next, we introduce the collection of FHIR condition entries shown in the CDS card response.

###### condition resources

Condition resources that are not included directly within another resource (such as FHIR forecastEffect, a TMR-specialised FHIR type which it is introduced below) represent initial states, or situations, of a TMR transition, that is, the state in which the measured clinical property is found prior application of a care action. In this CDS card, we have the following entries:

{

"resource": {

"resourceType": "Condition",

"id": "SitHghRskFlu",

"code": {

"coding": [

{

"system": "http://anonymous.org/data/SitHghRskFlu",

"code": "SitHghRskFlu",

"display": "having a high risk of contracting influenza disease"

}

]

},

"subject": {

"reference": "Patient/1677163"

}

}

},

{

"resource": {

"resourceType": "Condition",

"id": "SitPrQol",

"code": {

"coding": [

{

"system": "http://anonymous.org/data/SitPrQol",

"code": "SitPrQol",

"display": "having a poor quality of life"

}

]

},

"subject": {

"reference": "Patient/1677163"

}

}

},

{

"resource": {

"resourceType": "Condition",

"id": "SitLwRskPneumn",

"code": {

"coding": [

{

"system": "http://anonymous.org/data/SitLwRskPneumn",

"code": "SitLwRskPneumn",

"display": "having a low risk of contracting pneumonia"

}

]

},

"subject": {

"reference": "Patient/1677163"

}

}

},

{

"resource": {

"resourceType": "Condition",

"id": "SitVerySevAls",

"code": {

"coding": [

{

"system": "http://anonymous.org/data/SitVerySevAls",

"code": "SitVerySevAls",

"display": "having a very severe airflow limitation severity"

}

]

},

"subject": {

"reference": "Patient/1677163"

}

}

},

{

"resource": {

"resourceType": "Condition",

"id": "SitPrLngHlth",

"code": {

"coding": [

{

"system": "http://anonymous.org/data/SitPrLngHlth",

"code": "SitPrLngHlth",

"display": "having a poor pulmonary health"

}

]

},

"subject": {

"reference": "Patient/1677163"

}

}

},

{

"resource": {

"resourceType": "Condition",

"id": "SitLwRskCrd",

"code": {

"coding": [

{

"system": "http://anonymous.org/data/SitLwRskCrd",

"code": "SitLwRskCrd",

"display": "low risk of having cardiac rhythm disturbances"

}

]

},

"subject": {

"reference": "Patient/1677163"

}

}

}

Condition resources are referenced by forecastEffect resources, which we introduce next.

###### Forecasteffect resources

FHIR forecastEffect was designed to represent a TMR causation belief term, that is, the expected main, and possibly secondary, effects of administering a care action to a patient to change the state of a clinical property. The definition of FHIR forecastEffect can be found in <https://github.com/susoDominguez/ROAD2H-hooks>. A descriptive example is given next, after introducing the collection of forecastEffect entries which are part of the CDS card response.

{

"resource": {

"resourceType": "ForecastEffect",

"id": "SitHghRskFlu2SitLwRskFluMp",

"typeOfEffect": "main-effect",

"typeOfEvent": "therapeutic-effect",

"subject": {

"reference": "Patient/1677163"

},

"appliesTo": {

"careActionInstance": [

{

"reference": "MedicationRequest/RecCOPD-FluVacDecPropRskFluShould"

}

],

"conditionAddressed": {

"reference": "Condition/SitHghRskFlu"

}

},

"expectedOutcomeCode": {

"coding": [

{

"system": "http://anonymous.org/data/SitLwRskFlu",

"code": "SitLwRskFlu",

"display": "having a low risk of contracting influenza disease"

}

]

},

"targetMeasurement": {

"measuredProperty": {

"coding": [

{

"system": "http://anonymous.org/data/PropRskFlu",

"code": "PropRskFlu",

"display": "risk of contracting influenza disease"

}

]

},

"degreeOfChange": "decrease"

},

"probability": "always",

"evidence": "high"

}

},

{

"resource": {

"resourceType": "ForecastEffect",

"id": "SitPrQol2SitNrmQolMp",

"typeOfEffect": "main-effect",

"typeOfEvent": "therapeutic-effect",

"subject": {

"reference": "Patient/1677163"

},

"appliesTo": {

"careActionInstance": [

{

"reference": "ServiceRequest/RecCOPD-SmokeThrpyIncPropQolShould"

}

],

"conditionAddressed": {

"reference": "Condition/SitPrQol"

}

},

"expectedOutcomeCode": {

"coding": [

{

"system": "http://anonymous.org/data/SitNrmQol",

"code": "SitNrmQol",

"display": "having a standard quality of life"

}

]

},

"targetMeasurement": {

"measuredProperty": {

"coding": [

{

"system": "http://anonymous.org/data/PropQol",

"code": "PropQol",

"display": "quality of life"

}

]

},

"degreeOfChange": "increase"

},

"probability": "always",

"evidence": "high"

}

},

{

"resource": {

"resourceType": "ForecastEffect",

"id": "SitLwRskPneumn2SitHghRskPneumnMn",

"typeOfEffect": "main-effect",

"typeOfEvent": "adverse-effect",

"subject": {

"reference": "Patient/1677163"

},

"appliesTo": {

"careActionInstance": [

{

"reference": "MedicationRequest/RecCOPD-IcsIncPropRskPneumnShouldnot"

}

],

"conditionAddressed": {

"reference": "Condition/SitLwRskPneumn"

}

},

"expectedOutcomeCode": {

"coding": [

{

"system": "http://anonymous.org/data/SitHghRskPneumn",

"code": "SitHghRskPneumn",

"display": "having a high risk of contracting pneumonia"

}

]

},

"targetMeasurement": {

"measuredProperty": {

"coding": [

{

"system": "http://anonymous.org/data/PropRskPneumn",

"code": "PropRskPneumn",

"display": "risk of pneumonia"

}

]

},

"degreeOfChange": "increase"

},

"probability": "always",

"evidence": "high"

}

},

{

"resource": {

"resourceType": "ForecastEffect",

"id": "SitVerySevAls2SitSevAlsMp",

"typeOfEffect": "main-effect",

"typeOfEvent": "therapeutic-effect",

"subject": {

"reference": "Patient/1677163"

},

"appliesTo": {

"careActionInstance": [

{

"reference": "MedicationRequest/RecCOPD-LabaIcsDecVerySevPropAlsShould"

},

{

"reference": "MedicationRequest/RecCOPD-LamaDecVerySevPropAlsShould"

},

{

"reference": "MedicationRequest/RecCOPD-LabaLamaDecVerySevPropAlsShould"

},

{

"reference": "MedicationRequest/RecCOPD-LabaLamaIcsDecVerySevPropAlsShould"

}

],

"conditionAddressed": {

"reference": "Condition/SitVerySevAls"

}

},

"expectedOutcomeCode": {

"coding": [

{

"system": "http://anonymous.org/data/SitSevAls",

"code": "SitSevAls",

"display": "having a severe airflow limitation severity"

}

]

},

"targetMeasurement": {

"measuredProperty": {

"coding": [

{

"system": "http://anonymous.org/data/PropAls",

"code": "PropAls",

"display": "airflow limitation severity"

}

]

},

"degreeOfChange": "decrease"

},

"probability": "always",

"evidence": "high"

}

},

{

"resource": {

"resourceType": "ForecastEffect",

"id": "SitPrLngHlth2SitNrmLngHlthMp",

"typeOfEffect": "main-effect",

"typeOfEvent": "therapeutic-effect",

"subject": {

"reference": "Patient/1677163"

},

"appliesTo": {

"careActionInstance": [

{

"reference": "ServiceRequest/RecCOPD-LngRehabIncPropLngHlthShould"

}

],

"conditionAddressed": {

"reference": "Condition/SitPrLngHlth"

}

},

"expectedOutcomeCode": {

"coding": [

{

"system": "http://anonymous.org/data/SitNrmLngHlth",

"code": "SitNrmLngHlth",

"display": "having a standard pulmonary health"

}

]

},

"targetMeasurement": {

"measuredProperty": {

"coding": [

{

"system": "http://anonymous.org/data/PropLngHlth",

"code": "PropLngHlth",

"display": "pulmonary health"

}

]

},

"degreeOfChange": "increase"

},

"probability": "always",

"evidence": "high"

}

},

{

"resource": {

"resourceType": "ForecastEffect",

"id": "SitLwRskCrd2SitHghRskCrdMn",

"typeOfEffect": "main-effect",

"typeOfEvent": "adverse-effect",

"subject": {

"reference": "Patient/1677163"

},

"appliesTo": {

"careActionInstance": [

{

"reference": "MedicationRequest/RecCOPD-BetaAgonistIncPropRskCrdShouldnot"

}

],

"conditionAddressed": {

"reference": "Condition/SitLwRskCrd"

}

},

"expectedOutcomeCode": {

"coding": [

{

"system": "http://anonymous.org/data/SitHghRskCrd",

"code": "SitHghRskCrd",

"display": "high risk of having cardiac rhythm disturbances"

}

]

},

"targetMeasurement": {

"measuredProperty": {

"coding": [

{

"system": "http://anonymous.org/data/PropRskCrd",

"code": "PropRskCrd",

"display": "risk of cardiac rhythm disturbances"

}

]

},

"degreeOfChange": "increase"

},

"probability": "always",

"evidence": "high"

}

}

Let’s go over the forecastEffect instance with id = SitVerySevAls2SitSevAlsMp. In a nutshell, this resource represents the expected effect of administering COPD treatments to a patient with very severe COPD symptoms, that is, a patient assessed to follow treatment pathways from GOLD group D. The type of effect is denoted as main-effect (for this proof-of-concept we did not include side-effects) and the type of event as therapeutical, that is, the effect of the care action, or actions, has a positive result in the overall health of the patient, particularly in *decreasing* (see parameter degreeOfChange) the condition addressed -represented as a reference to one of the condition resources we introduced earlier-: *having a very severe airflow limitation severity*. The expected outcome of this forecastEffect is also represented in the resource: *having a severe airflow limitation severity.* Both the initial state and expected outcome share the same measured property, that is, the property that the user wants to change by administering a care action. In this case, the measured property is the *airflow limitation severity*. Observe that this causation belief, or forecastEffect resource, is shared by more than one care action, in fact, it is share by four care actions, each representing a medicationRequest resource for each of the personalised COPD treatments from GOLD group D suitable for the current patient. MedicationRequest resources are introduced next. Finally, the forecastEffect resource identifies the probability (always) of this effect happening as described, and the level of evidence (high) found in the literature to make such claim. Parameters probability and evidence have fixed values for all forecastEffect resources in this iteration of the project.

###### medicationrequest and servicerequest resources

MedicationRequest and ServiceRequest resources describe the reasons and outcomes of care actions in a patient. The former is applicable to (drug-based) treatments and the latter to therapies.

{

"resource": {

"resourceType": "ServiceRequest",

"id": "RecCOPD-SmokeThrpyIncPropQolShould",

"status": "active",

"intent": "plan",

"instantiatesUri": "https://goldcopd.org/wp-content/uploads/2016/12/wms-GOLD-2017-Pocket-Guide.pdf",

"doNotPerform": false,

"reasonReference": [

{

"reference": "Condition/SitPrQol"

}

],

"forecast-effects": [

{

"reference": "ForecastEffect/SitPrQol2SitNrmQolMp"

}

],

"code": {

"coding": [

{

"system": "http://anonymous.org/data/NonDrugTSmokeThrpy",

"code": "SmokeThrpy",

"display": "administer smoking cessation therapy"

}

]

},

"subject": {

"reference": "Patient/1677163"

}

}

},

{

"resource": {

"resourceType": "ServiceRequest",

"id": "RecCOPD-LngRehabIncPropLngHlthShould",

"status": "active",

"intent": "plan",

"instantiatesUri": "https://goldcopd.org/wp-content/uploads/2016/12/wms-GOLD-2017-Pocket-Guide.pdf",

"doNotPerform": false,

"reasonReference": [

{

"reference": "Condition/SitPrLngHlth"

}

],

"forecast-effects": [

{

"reference": "ForecastEffect/SitPrLngHlth2SitNrmLngHlthMp"

}

],

"code": {

"coding": [

{

"system": "http://anonymous.org/data/NonDrugTLngRehab",

"code": "LngRehab",

"display": "administer pulmonary rehabilitation"

}

]

},

"subject": {

"reference": "Patient/1677163"

}

}

},

{

"resource": {

"resourceType": "MedicationRequest",

"id": "RecCOPD-FluVacDecPropRskFluShould",

"status": "active",

"intent": "plan",

"instantiatesUri": "https://goldcopd.org/wp-content/uploads/2016/12/wms-GOLD-2017-Pocket-Guide.pdf",

"doNotPerform": false,

"reasonReference": [

{

"reference": "Condition/SitHghRskFlu"

}

],

"forecast-effects": [

{

"reference": "ForecastEffect/SitHghRskFlu2SitLwRskFluMp"

}

],

"medicationReference": {

"reference": "Medication/DrugTFluVac"

},

"subject": {

"reference": "Patient/1677163"

},

"detectedIssue": []

}

},

{

"resource": {

"resourceType": "MedicationRequest",

"id": "RecCOPD-IcsIncPropRskPneumnShouldnot",

"status": "active",

"intent": "plan",

"instantiatesUri": "https://goldcopd.org/wp-content/uploads/2016/12/wms-GOLD-2017-Pocket-Guide.pdf",

"doNotPerform": true,

"reasonReference": [

{

"reference": "Condition/SitLwRskPneumn"

}

],

"forecast-effects": [

{

"reference": "ForecastEffect/SitLwRskPneumn2SitHghRskPneumnMn"

}

],

"medicationReference": {

"reference": "Medication/DrugTIcs"

},

"subject": {

"reference": "Patient/1677163"

},

"detectedIssue": [

{

"reference": "DetectedIssue/contradiction2"

},

{

"reference": "DetectedIssue/contradiction3"

}

]

}

},

{

"resource": {

"resourceType": "MedicationRequest",

"id": "RecCOPD-LabaIcsDecVerySevPropAlsShould",

"status": "active",

"intent": "plan",

"instantiatesUri": "https://goldcopd.org/wp-content/uploads/2016/12/wms-GOLD-2017-Pocket-Guide.pdf",

"doNotPerform": false,

"reasonReference": [

{

"reference": "Condition/SitVerySevAls"

}

],

"forecast-effects": [

{

"reference": "ForecastEffect/SitVerySevAls2SitSevAlsMp"

}

],

"medicationReference": {

"reference": "Medication/DrugCatLabaIcs"

},

"subject": {

"reference": "Patient/1677163"

},

"detectedIssue": [

{

"reference": "DetectedIssue/alternative1"

},

{

"reference": "DetectedIssue/contradiction2"

}

]

}

},

{

"resource": {

"resourceType": "MedicationRequest",

"id": "RecCOPD-LamaDecVerySevPropAlsShould",

"status": "active",

"intent": "plan",

"instantiatesUri": "https://goldcopd.org/wp-content/uploads/2016/12/wms-GOLD-2017-Pocket-Guide.pdf",

"doNotPerform": false,

"reasonReference": [

{

"reference": "Condition/SitVerySevAls"

}

],

"forecast-effects": [

{

"reference": "ForecastEffect/SitVerySevAls2SitSevAlsMp"

}

],

"medicationReference": {

"reference": "Medication/DrugTLama"

},

"subject": {

"reference": "Patient/1677163"

},

"detectedIssue": [

{

"reference": "DetectedIssue/repetition0"

},

{

"reference": "DetectedIssue/alternative1"

}

]

}

},

{

"resource": {

"resourceType": "MedicationRequest",

"id": "RecCOPD-LabaLamaDecVerySevPropAlsShould",

"status": "active",

"intent": "plan",

"instantiatesUri": "https://goldcopd.org/wp-content/uploads/2016/12/wms-GOLD-2017-Pocket-Guide.pdf",

"doNotPerform": false,

"reasonReference": [

{

"reference": "Condition/SitVerySevAls"

}

],

"forecast-effects": [

{

"reference": "ForecastEffect/SitVerySevAls2SitSevAlsMp"

}

],

"medicationReference": {

"reference": "Medication/DrugCatLabaLama"

},

"subject": {

"reference": "Patient/1677163"

},

"detectedIssue": [

{

"reference": "DetectedIssue/repetition0"

},

{

"reference": "DetectedIssue/alternative1"

}

]

}

},

{

"resource": {

"resourceType": "MedicationRequest",

"id": "RecCOPD-BetaAgonistIncPropRskCrdShouldnot",

"status": "active",

"intent": "plan",

"instantiatesUri": "https://goldcopd.org/wp-content/uploads/2016/12/wms-GOLD-2017-Pocket-Guide.pdf",

"doNotPerform": true,

"reasonReference": [

{

"reference": "Condition/SitLwRskCrd"

}

],

"forecast-effects": [

{

"reference": "ForecastEffect/SitLwRskCrd2SitHghRskCrdMn"

}

],

"medicationReference": {

"reference": "Medication/DrugCatBetaAgonist"

},

"subject": {

"reference": "Patient/1677163"

},

"detectedIssue": []

}

},

{

"resource": {

"resourceType": "MedicationRequest",

"id": "RecCOPD-LabaLamaIcsDecVerySevPropAlsShould",

"status": "active",

"intent": "plan",

"instantiatesUri": "https://goldcopd.org/wp-content/uploads/2016/12/wms-GOLD-2017-Pocket-Guide.pdf",

"doNotPerform": false,

"reasonReference": [

{

"reference": "Condition/SitVerySevAls"

}

],

"forecast-effects": [

{

"reference": "ForecastEffect/SitVerySevAls2SitSevAlsMp"

}

],

"medicationReference": {

"reference": "Medication/DrugCatLabaLamaIcs"

},

"subject": {

"reference": "Patient/1677163"

},

"detectedIssue": [

{

"reference": "DetectedIssue/repetition0"

},

{

"reference": "DetectedIssue/alternative1"

},

{

"reference": "DetectedIssue/contradiction3"

}

]

}

}

MedicationRequest and ServiceRequest resources are intended as part of a care plan ("intent": "plan") and the evidence is taken from the GOLD guideline ("instantiatesUri": "https://goldcopd.org/wp-content/uploads/2016/12/wms-GOLD-2017-Pocket-Guide.pdf"). Each resource references one or more forecastEffect instances (a main-effect and zero or more side-effect instances). For instance, as we demonstrated above, each of the MedicationRequest instances representing a COPD treatment from GOLD group D have the same forecastEffect reference, and, equally, the same condition referencing the initial state (very severe) of the airflow limitation severity. A crucial part of either resource is the parameter "doNotPerform", which can be either true, then the recommendation is to not administer the care action, or false, which implies the recommendation has positive effects and thus it should be applied. Among other parameters, we also have "detectedIssue", which references resources of the same name, and identifies potential interactions that have been previously detected by the logic rules of the TMR framework.

Following the codes and references from each MedicationRequest and ServiceRequest we can deduce the following information (starting at the top of the shown resources):

- A request to administer smoking cessation therapy towards having a standard quality of life.
- A request to administer pulmonary rehabilitation towards having a standard pulmonary health.
- A request to administer the influenza vaccina towards having a low risk of contracting influenza disease.
- A request to NOT administer ICS to avoid having a high risk of contracting pneumonia.
  - A contradiction interaction has been detected regarding this request.
- A request to administer a combination of LABA + ICS towards decreasing airflow limitation severity from very severe to severe.
  - An alternative interaction has been detected regarding this request.
  - A contradiction interaction has been detected regarding this request.
- A request to administer LAMA towards decreasing airflow limitation severity from very severe to severe.
  - A repetition interaction has been detected regarding this request.
  - An alternative interaction has been detected regarding this request.
- A request to administer a combination of LABA + LAMA towards decreasing airflow limitation severity from very severe to severe.
  - A repetition interaction has been detected regarding this request.
  - An alternative interaction has been detected regarding this request.
- A request to NOT administer beta agonists when cardiovascular disease is present to avoid risk of cardiac rhythm disturbances from having a high risk to a low risk.
  - A contradiction interaction has been detected regarding this request.
- A request to administer a combination of LABA + LAMA + ICS towards decreasing airflow limitation severity from very severe to severe.
  - A repetition interaction has been detected regarding this request.
  - An alternative interaction has been detected regarding this request.
  - A contradiction interaction has been detected regarding this request.

We go over FHIR DetectedIssue types next.

###### detectedissue resources

As stated in the FHIR resource types page (<http://www.hl7.org/fhir/detectedissue.html>), DetectedIssue resources indicate an actual or potential clinical issue with or between one or more active or proposed clinical actions for a patient; e.g. Drug-drug interaction, Ineffective treatment frequency, Procedure-condition conflict, etc.

Let’s have a look at the instances shown in the CDS card:

{

"resource": {

"resourceType": "DetectedIssue",

"id": "repetition0",

"status": "preliminary",

"code": {

"coding": [

{

"system": "http://terminology.hl7.org/CodeSystem/v3-ActCode",

"code": "DUPTHPY",

"display": "Duplicate Therapy Alert"

}

]

},

"implicated": [

{

"reference": "MedicationRequest/RecCOPD-LabaLamaDecVerySevPropAlsShould"

},

{

"reference": "MedicationRequest/RecCOPD-LabaLamaIcsDecVerySevPropAlsShould"

},

{

"reference": "MedicationRequest/RecCOPD-LamaDecVerySevPropAlsShould"

}

],

"mitigation": [

{

"action": {

"coding": [

{

"system": "http://terminology.hl7.org/CodeSystem/v3-ActCode",

"code": "13",

"display": "Stopped Concurrent Therapy"

}

]

}

}

]

}

},

{

"resource": {

"resourceType": "DetectedIssue",

"id": "alternative1",

"status": "preliminary",

"code": {

"coding": [

{

"system": "http://anonymous.org/CodeSystem/interactions",

"code": "ALTHRPY",

"display": "Alternative Therapies With Same Intended Effect"

}

]

},

"implicated": [

{

"reference": "MedicationRequest/RecCOPD-LabaIcsDecVerySevPropAlsShould"

},

{

"reference": "MedicationRequest/RecCOPD-LabaLamaDecVerySevPropAlsShould"

},

{

"reference": "MedicationRequest/RecCOPD-LabaLamaIcsDecVerySevPropAlsShould"

},

{

"reference": "MedicationRequest/RecCOPD-LamaDecVerySevPropAlsShould"

}

],

"mitigation": [

{

"action": {

"coding": [

{

"system": "http://anonymous.org/CodeSystem/interactions",

"code": "NOTREQ",

"display": "Mitigation Not Required"

}

]

}

}

]

}

},

{

"resource": {

"resourceType": "DetectedIssue",

"id": "contradiction2",

"status": "preliminary",

"code": {

"coding": [

{

"system": "http://terminology.hl7.org/CodeSystem/v3-ActCode",

"code": "DACT",

"display": "Drug Action Detected Issue"

}

]

},

"implicated": [

{

"reference": "MedicationRequest/RecCOPD-IcsIncPropRskPneumnShouldnot"

},

{

"reference": "MedicationRequest/RecCOPD-LabaIcsDecVerySevPropAlsShould"

}

],

"mitigation": [

{

"action": {

"coding": [

{

"system": "http://terminology.hl7.org/CodeSystem/v3-ActCode",

"code": "13",

"display": "Stopped Concurrent Therapy"

}

]

}

}

]

}

},

{

"resource": {

"resourceType": "DetectedIssue",

"id": "contradiction3",

"status": "preliminary",

"code": {

"coding": [

{

"system": "http://terminology.hl7.org/CodeSystem/v3-ActCode",

"code": "DACT",

"display": "Drug Action Detected Issue"

}

]

},

"implicated": [

{

"reference": "MedicationRequest/RecCOPD-IcsIncPropRskPneumnShouldnot"

},

{

"reference": "MedicationRequest/RecCOPD-LabaLamaIcsDecVerySevPropAlsShould"

}

],

"mitigation": [

{

"action": {

"coding": [

{

"system": "http://terminology.hl7.org/CodeSystem/v3-ActCode",

"code": "13",

"display": "Stopped Concurrent Therapy"

}

]

}

}

]

}

}

The DetectedIssue resource identifies the interaction, the implicated FHIR MedicationRequest instances and the mitigation applied by the conflict mitigation service. The instances are self-explanatory hence we move on to the last part, the FHIR carePlan which, with support from the conflict mitigation service, distributes MedicationRequest and ServiceRequest resources into personalised conflict-safe care plan proposals.

###### careplan resources

There are four proposals designed by the COPD-CDS system as COPD management care plans. One for each COPD treatment included.

{

"resource": {

"resourceType": "CarePlan",

"id": "CarePlan0",

"status": "active",

"intent": "plan",

"title": "suggested treatments: LABA + LAMA ",

"subject": {

"reference": "Patient/1677163"

},

"activity": [

{

"reference": "MedicationRequest/RecCOPD-LabaLamaDecVerySevPropAlsShould"

},

{

"reference": "MedicationRequest/RecCOPD-IcsIncPropRskPneumnShouldnot"

},

{

"reference": "ServiceRequest/RecCOPD-SmokeThrpyIncPropQolShould"

},

{

"reference": "ServiceRequest/RecCOPD-LngRehabIncPropLngHlthShould"

},

{

"reference": "MedicationRequest/RecCOPD-BetaAgonistIncPropRskCrdShouldnot"

},

{

"reference": "MedicationRequest/RecCOPD-FluVacDecPropRskFluShould"

}

]

}

},

{

"resource": {

"resourceType": "CarePlan",

"id": "CarePlan1",

"status": "active",

"intent": "plan",

"title": "suggested treatments: LAMA ",

"subject": {

"reference": "Patient/1677163"

},

"activity": [

{

"reference": "MedicationRequest/RecCOPD-LamaDecVerySevPropAlsShould"

},

{

"reference": "MedicationRequest/RecCOPD-IcsIncPropRskPneumnShouldnot"

},

{

"reference": "ServiceRequest/RecCOPD-SmokeThrpyIncPropQolShould"

},

{

"reference": "ServiceRequest/RecCOPD-LngRehabIncPropLngHlthShould"

},

{

"reference": "MedicationRequest/RecCOPD-BetaAgonistIncPropRskCrdShouldnot"

},

{

"reference": "MedicationRequest/RecCOPD-FluVacDecPropRskFluShould"

}

]

}

},

{

"resource": {

"resourceType": "CarePlan",

"id": "CarePlan2",

"status": "active",

"intent": "plan",

"title": "suggested treatments: LABA + ICS ",

"subject": {

"reference": "Patient/1677163"

},

"activity": [

{

"reference": "ServiceRequest/RecCOPD-SmokeThrpyIncPropQolShould"

},

{

"reference": "ServiceRequest/RecCOPD-LngRehabIncPropLngHlthShould"

},

{

"reference": "MedicationRequest/RecCOPD-BetaAgonistIncPropRskCrdShouldnot"

},

{

"reference": "MedicationRequest/RecCOPD-FluVacDecPropRskFluShould"

},

{

"reference": "MedicationRequest/RecCOPD-LabaIcsDecVerySevPropAlsShould"

}

]

}

},

{

"resource": {

"resourceType": "CarePlan",

"id": "CarePlan3",

"status": "active",

"intent": "plan",

"title": "suggested treatments: LABA + LAMA + ICS ",

"subject": {

"reference": "Patient/1677163"

},

"activity": [

{

"reference": "ServiceRequest/RecCOPD-SmokeThrpyIncPropQolShould"

},

{

"reference": "ServiceRequest/RecCOPD-LngRehabIncPropLngHlthShould"

},

{

"reference": "MedicationRequest/RecCOPD-BetaAgonistIncPropRskCrdShouldnot"

},

{

"reference": "MedicationRequest/RecCOPD-FluVacDecPropRskFluShould"

},

{

"reference": "MedicationRequest/RecCOPD-LabaLamaIcsDecVerySevPropAlsShould"

}

]

}

}

For each of the carePlan instances, the main COPD drug type or drug type combination has been identified and added as part of the title, for a quick reference for the user to view. Observe that the carePlan instance is simply a collection of references to FHIR MedicationRequest and ServiceRequest instances that have been introduced above. The carePlan resource is the entry point for the graphical user interface to display information relevant to the current patient and the management of their COPD symptoms.
